## Supplement for "The Impact of Vaccination to Control COVID-19 Burden in the United States: A Simulation Modeling Approach"

**Supplementary Material**

### Section A. Details of COVAM

#### Overview of COVAM

COVAM is an agent-based simulation model in which agents represent the individuals in a community. Full details of COVAM are published elsewhere^1^. Key parameters used in COVAM are given in Supplement Table 1.

Supplement Table 1. COVAM parameters specific to regions. (adapted from Alagoz, et al 2020 ^1^)

| Parameter | Value, by region | | | Source |
| --- | --- | --- | --- | --- |
|  | Dane County | Milwaukee Metro | New York City |  |
| Population size | 542,364 | 1,576,236 | 8,398,748 | US Census data^2^ |
| Date of simulation start | March 4th, 2020 | March 4th, 2020 | March 4th, 2020 | -- |
| Number of daily imported exposed agents and dates of importation | 3 daily from Mar 5- 25, 2020; 1 daily onward | 6 daily from Mar 5-25, 2020; 3 daily onward | 160 daily from Mar 5-25, 2020; 32 daily onward | Calibration |
| Number of daily close contacts without social distancing | 10 | 10 | 20 | Calibration and literature^3,4^ |
| Adherence to social distancing, expressed as percentage drop in number of contacts | Mar 4 - 11, 2020: 0% Mar 12 - 25, 2020: linear increase from 0 - 91% Mar 26 - Apr 30, 2020: 80% May 1 - 13, 2020: 70%  May 14 - June 30, 2020: 60%  July 1 - Aug 31, 2020: 70%  Sep 1 - Nov 8, 2020: 65% Nov 9, 2020 – end of simulation: 70% | Mar 4 – 11, 2020: 0% Mar 12 - 22, 2020: linear increase from 0 - 60% Mar 23 - Apr 7, 2020: 60% Apr 8 - 12, 2020: 80% Apr 13 - May 13, 2020: 70% May 14 - 25, 2020: 60% May 26 - July 15, 2020: 70% July 16 - Aug 14, 2020: 65%  Aug 15 - Sep 29, 2020: 70%  Sep 30 - Oct 29, 2020: 60%  Oct 30 - Nov 8, 2020: 65%  Nov 9, 2020– end of simulation: 70% | Mar 4 - 11, 2020: 0% Mar 12 - 25, 2020: linear increase from 0 - 78% Mar 26 - Apr 17, 2020: 85%  Apr 18 - June 7, 2020: 90%  June 8 - July 1, 2020: 85%  July 2 - Aug 15, 2020: 90%  Aug 16 - Sep 14, 2020: 80%  Sep 15 - Nov 14, 2020: 85%  Nov 15 - Dec 5, 2020: 70%  Dec 6 - Dec 31, 2020: 85%  Jan 1, 2021 – end of simulation: 80% | Cellphone data and calibration^5-7^ |
| Probability that agent is tested | Mar 4 - Apr 14, 2020: 75% Apr 15 - May 9, 2020: 80% May 10, 2020 - end: 90% | Mar 4 - Apr 14, 2020: 75% Apr 15 - May 9, 2020: 80% May 10, 2020 - end: 90% | Mar 4 - Apr 14, 2020: 75% Apr 15 - May 9, 2020: 80% May 10, 2020 - end: 90% | Literature and calibration^8,9^ |
| Age distribution (percentage) | 0-19y: 20.8% 20-44y: 46.5% 45-54y: 10.3% 55-64y: 10.2% 65-74y: 7.4% 75-84y: 3.3% 85y+: 1.5% | 0-19y: 28.6% 20-44y: 36.2% 45-54y: 13.7% 55-64y: 8.5% 65-74y: 6.6% 75-84y: 4.7% 85y+: 1.0% | 0-19y: 23.2% 20-44y: 38.6% 45-54y: 13.0% 55-64y: 11.7% 65-74y: 7.1% 75-84y: 3.4% 85y+: 1.9% | US Census data^2^ |

### Section B. Model Validation Results

In this section, we present the updated results of the validation of COVAM. Model structure and all model inputs for Dane County, Milwaukee metro area, and NYC except vaccination and adherence to NPIs were fixed before July 31, 2020. The structure of the vaccination and vaccination-related inputs were fixed by December 15, 2020. Adherence to social distancing inputs were fixed by January 5, 2021 for Dane County and Milwaukee and by January 14, 2021 for NYC and the predictions of COVAM were compared against the reported number of confirmed cases over time after this date.

In these runs, we used the following settings for the model inputs: vaccination effectiveness is 90%, vaccination coverage is 50%, daily vaccination capacity is 0.1%, drop in adherence after vaccination is 20%, number of imported cases is 1, 3, and 16 for Dane County, Milwaukee, and NYC, respectively, and adherence to NPIs after February 1, 2021 is 70%, 70%, and 85%, for Dane County, Milwaukee, and NYC, respectively. The results in this section use the average values of 100 replications, providing steady estimates around the mean parameter values with small standard errors. Data on case numbers were obtained from the Wisconsin Department of Health Services and the NYC Department of Health.^10,11^

Supplement Figure 1. Model validation results. In each of the following figures, red dots represent the actual observed cumulative number of confirmed cases, the black solid line represents the model’s predictions, error bars around the black solid line represent 95% confidence intervals for the model’s predictions based on 100 replications, the green dotted line represents the date after which no model input parameter except adherence to NPIs was modified, and the blue dashed line represents the date after which no model input parameter was modified.

(a) Dane County

Total number of confirmed cases

|  | Dane County- actual confirmed cases | Alagoz model results |
| --- | --- | --- |
| 2020-03-15 | 10 | 30 |
| 2020-03-16 | 19 | 39 |
| 2020-03-17 | 23 | 51 |
| 2020-03-18 | 27 | 64 |
| 2020-03-19 | 32 | 79 |
| 2020-03-20 | 49 | 96 |
| 2020-03-21 | 61 | 116 |
| 2020-03-22 | 70 | 135 |
| 2020-03-23 | 72 | 155 |
| 2020-03-24 | 88 | 174 |
| 2020-03-25 | 114 | 193 |
| 2020-03-26 | 133 | 210 |
| 2020-03-27 | 158 | 225 |
| 2020-03-28 | 172 | 237 |
| 2020-03-29 | 187 | 244 |
| 2020-03-30 | 209 | 258 |
| 2020-03-31 | 232 | 272 |
| 2020-04-01 | 238 | 284 |
| 2020-04-02 | 252 | 295 |
| 2020-04-03 | 269 | 305 |
| 2020-04-04 | 280 | 314 |
| 2020-04-05 | 292 | 323 |
| 2020-04-06 | 303 | 332 |
| 2020-04-07 | 306 | 340 |
| 2020-04-08 | 313 | 348 |
| 2020-04-09 | 324 | 356 |
| 2020-04-10 | 332 | 363 |
| 2020-04-11 | 339 | 370 |
| 2020-04-12 | 342 | 377 |
| 2020-04-13 | 348 | 383 |
| 2020-04-14 | 350 | 390 |
| 2020-04-15 | 352 | 396 |
| 2020-04-16 | 357 | 402 |
| 2020-04-17 | 361 | 408 |
| 2020-04-18 | 365 | 414 |
| 2020-04-19 | 382 | 420 |
| 2020-04-20 | 387 | 425 |
| 2020-04-21 | 387 | 430 |
| 2020-04-22 | 390 | 435 |
| 2020-04-23 | 399 | 440 |
| 2020-04-24 | 401 | 445 |
| 2020-04-25 | 405 | 449 |
| 2020-04-26 | 414 | 454 |
| 2020-04-27 | 417 | 459 |
| 2020-04-28 | 424 | 463 |
| 2020-04-29 | 428 | 467 |
| 2020-04-30 | 430 | 472 |
| 2020-05-01 | 433 | 476 |
| 2020-05-02 | 438 | 479 |
| 2020-05-03 | 444 | 483 |
| 2020-05-04 | 444 | 487 |
| 2020-05-05 | 445 | 492 |
| 2020-05-06 | 447 | 497 |
| 2020-05-07 | 455 | 502 |
| 2020-05-08 | 465 | 507 |
| 2020-05-09 | 472 | 512 |
| 2020-05-10 | 474 | 519 |
| 2020-05-11 | 477 | 525 |
| 2020-05-12 | 490 | 531 |
| 2020-05-13 | 494 | 537 |
| 2020-05-14 | 507 | 544 |
| 2020-05-15 | 519 | 551 |
| 2020-05-16 | 537 | 558 |
| 2020-05-17 | 538 | 565 |
| 2020-05-18 | 541 | 574 |
| 2020-05-19 | 563 | 583 |
| 2020-05-20 | 576 | 593 |
| 2020-05-21 | 587 | 603 |
| 2020-05-22 | 594 | 614 |
| 2020-05-23 | 596 | 625 |
| 2020-05-24 | 604 | 636 |
| 2020-05-25 | 630 | 649 |
| 2020-05-26 | 653 | 662 |
| 2020-05-27 | 679 | 676 |
| 2020-05-28 | 705 | 691 |
| 2020-05-29 | 723 | 707 |
| 2020-05-30 | 730 | 723 |
| 2020-05-31 | 735 | 740 |
| 2020-06-01 | 759 | 759 |
| 2020-06-02 | 773 | 778 |
| 2020-06-03 | 789 | 797 |
| 2020-06-04 | 800 | 818 |
| 2020-06-05 | 814 | 840 |
| 2020-06-06 | 843 | 863 |
| 2020-06-07 | 856 | 887 |
| 2020-06-08 | 871 | 913 |
| 2020-06-09 | 884 | 940 |
| 2020-06-10 | 896 | 968 |
| 2020-06-11 | 930 | 997 |
| 2020-06-12 | 933 | 1028 |
| 2020-06-13 | 956 | 1061 |
| 2020-06-14 | 968 | 1096 |
| 2020-06-15 | 980 | 1132 |
| 2020-06-16 | 1000 | 1169 |
| 2020-06-17 | 1023 | 1207 |
| 2020-06-18 | 1046 | 1249 |
| 2020-06-19 | 1071 | 1292 |
| 2020-06-20 | 1111 | 1338 |
| 2020-06-21 | 1141 | 1385 |
| 2020-06-22 | 1172 | 1434 |
| 2020-06-23 | 1245 | 1486 |
| 2020-06-24 | 1350 | 1541 |
| 2020-06-25 | 1446 | 1597 |
| 2020-06-26 | 1525 | 1657 |
| 2020-06-27 | 1650 | 1721 |
| 2020-06-28 | 1751 | 1787 |
| 2020-06-29 | 1876 | 1855 |
| 2020-06-30 | 1952 | 1927 |
| 2020-07-01 | 2075 | 2003 |
| 2020-07-02 | 2192 | 2082 |
| 2020-07-03 | 2302 | 2165 |
| 2020-07-04 | 2381 | 2251 |
| 2020-07-05 | 2475 | 2320 |
| 2020-07-06 | 2510 | 2392 |
| 2020-07-07 | 2597 | 2467 |
| 2020-07-08 | 2652 | 2541 |
| 2020-07-09 | 2771 | 2616 |
| 2020-07-10 | 2889 | 2692 |
| 2020-07-11 | 2995 | 2768 |
| 2020-07-12 | 3071 | 2846 |
| 2020-07-13 | 3151 | 2922 |
| 2020-07-14 | 3250 | 2999 |
| 2020-07-15 | 3298 | 3073 |
| 2020-07-16 | 3376 | 3147 |
| 2020-07-17 | 3438 | 3222 |
| 2020-07-18 | 3495 | 3299 |
| 2020-07-19 | 3529 | 3376 |
| 2020-07-20 | 3562 | 3453 |
| 2020-07-21 | 3623 | 3528 |
| 2020-07-22 | 3666 | 3605 |
| 2020-07-23 | 3729 | 3682 |
| 2020-07-24 | 3776 | 3757 |
| 2020-07-25 | 3825 | 3832 |
| 2020-07-26 | 3870 | 3908 |
| 2020-07-27 | 3904 | 3983 |
| 2020-07-28 | 3970 | 4060 |
| 2020-07-29 | 4007 | 4137 |
| 2020-07-30 | 4086 | 4213 |
| 2020-07-31 | 4112 | 4290 |
| 2020-08-01 | 4198 | 4364 |
| 2020-08-02 | 4230 | 4439 |
| 2020-08-03 | 4263 | 4516 |
| 2020-08-04 | 4340 | 4592 |
| 2020-08-05 | 4376 | 4669 |
| 2020-08-06 | 4436 | 4747 |
| 2020-08-07 | 4492 | 4823 |
| 2020-08-08 | 4536 | 4900 |
| 2020-08-09 | 4554 | 4977 |
| 2020-08-10 | 4578 | 5055 |
| 2020-08-11 | 4630 | 5132 |
| 2020-08-12 | 4666 | 5210 |
| 2020-08-13 | 4749 | 5288 |
| 2020-08-14 | 4783 | 5366 |
| 2020-08-15 | 4849 | 5443 |
| 2020-08-16 | 4874 | 5520 |
| 2020-08-17 | 4890 | 5597 |
| 2020-08-18 | 4924 | 5675 |
| 2020-08-19 | 4955 | 5751 |
| 2020-08-20 | 4998 | 5829 |
| 2020-08-21 | 5053 | 5906 |
| 2020-08-22 | 5088 | 5983 |
| 2020-08-23 | 5117 | 6061 |
| 2020-08-24 | 5161 | 6137 |
| 2020-08-25 | 5209 | 6213 |
| 2020-08-26 | 5294 | 6290 |
| 2020-08-27 | 5330 | 6368 |
| 2020-08-28 | 5367 | 6445 |
| 2020-08-29 | 5426 | 6523 |
| 2020-08-30 | 5429 | 6601 |
| 2020-08-31 | 5520 | 6676 |
| 2020-09-01 | 5564 | 6754 |
| 2020-09-02 | 5639 | 6831 |
| 2020-09-03 | 5799 | 6908 |
| 2020-09-04 | 5961 | 6985 |
| 2020-09-05 | 6118 | 7075 |
| 2020-09-06 | 6208 | 7164 |
| 2020-09-07 | 6427 | 7254 |
| 2020-09-08 | 6489 | 7346 |
| 2020-09-09 | 6971 | 7439 |
| 2020-09-10 | 7254 | 7533 |
| 2020-09-11 | 7389 | 7631 |
| 2020-09-12 |  | 7731 |
| 2020-09-13 | 7846 | 7833 |
| 2020-09-14 | 7896 | 7940 |
| 2020-09-15 | 7984 | 8048 |
| 2020-09-16 | 8242 | 8157 |
| 2020-09-17 | 8455 | 8270 |
| 2020-09-18 | 8635 | 8386 |
| 2020-09-19 | 8815 | 8504 |
| 2020-09-20 | 8895 | 8623 |
| 2020-09-21 | 8947 | 8748 |
| 2020-09-22 | 9040 | 8874 |
| 2020-09-23 | 9203 | 9002 |
| 2020-09-24 | 9345 | 9133 |
| 2020-09-25 | 9488 | 9269 |
| 2020-09-26 | 9607 | 9406 |
| 2020-09-27 | 9763 | 9545 |
| 2020-09-28 | 9857 | 9689 |
| 2020-09-29 | 10129 | 9834 |
| 2020-09-30 | 10201 | 9984 |

|  | Validation begin date |
| --- | --- |
| 2021-01-04 | 100000 |

|  | Set Parameters |
| --- | --- |
| 2020-12-15 | 1000000 |

|  | Dane County- actual confirmed cases |
| --- | --- |
| 2020-03-15 | 10 |
| 2020-03-16 | 19 |
| 2020-03-17 | 23 |
| 2020-03-18 | 27 |
| 2020-03-19 | 32 |
| 2020-03-20 | 49 |
| 2020-03-21 | 61 |
| 2020-03-22 | 70 |
| 2020-03-23 | 72 |
| 2020-03-24 | 88 |
| 2020-03-25 | 114 |
| 2020-03-26 | 133 |
| 2020-03-27 | 158 |
| 2020-03-28 | 172 |
| 2020-03-29 | 187 |
| 2020-03-30 | 209 |
| 2020-03-31 | 232 |
| 2020-04-01 | 238 |
| 2020-04-02 | 252 |
| 2020-04-03 | 269 |
| 2020-04-04 | 280 |
| 2020-04-05 | 292 |
| 2020-04-06 | 303 |
| 2020-04-07 | 306 |
| 2020-04-08 | 313 |
| 2020-04-09 | 324 |
| 2020-04-10 | 332 |
| 2020-04-11 | 339 |
| 2020-04-12 | 342 |
| 2020-04-13 | 348 |
| 2020-04-14 | 350 |
| 2020-04-15 | 352 |
| 2020-04-16 | 357 |
| 2020-04-17 | 361 |
| 2020-04-18 | 365 |
| 2020-04-19 | 382 |
| 2020-04-20 | 387 |
| 2020-04-21 | 387 |
| 2020-04-22 | 390 |
| 2020-04-23 | 399 |
| 2020-04-24 | 401 |
| 2020-04-25 | 405 |
| 2020-04-26 | 414 |
| 2020-04-27 | 417 |
| 2020-04-28 | 424 |
| 2020-04-29 | 428 |
| 2020-04-30 | 430 |
| 2020-05-01 | 433 |
| 2020-05-02 | 438 |
| 2020-05-03 | 444 |
| 2020-05-04 | 444 |
| 2020-05-05 | 445 |
| 2020-05-06 | 447 |
| 2020-05-07 | 455 |
| 2020-05-08 | 465 |
| 2020-05-09 | 472 |
| 2020-05-10 | 474 |
| 2020-05-11 | 477 |
| 2020-05-12 | 490 |
| 2020-05-13 | 494 |
| 2020-05-14 | 507 |
| 2020-05-15 | 519 |
| 2020-05-16 | 537 |
| 2020-05-17 | 538 |
| 2020-05-18 | 541 |
| 2020-05-19 | 563 |
| 2020-05-20 | 576 |
| 2020-05-21 | 587 |
| 2020-05-22 | 594 |
| 2020-05-23 | 596 |
| 2020-05-24 | 604 |
| 2020-05-25 | 630 |
| 2020-05-26 | 653 |
| 2020-05-27 | 679 |
| 2020-05-28 | 705 |
| 2020-05-29 | 723 |
| 2020-05-30 | 730 |
| 2020-05-31 | 735 |
| 2020-06-01 | 759 |
| 2020-06-02 | 773 |
| 2020-06-03 | 789 |
| 2020-06-04 | 800 |
| 2020-06-05 | 814 |
| 2020-06-06 | 843 |
| 2020-06-07 | 856 |
| 2020-06-08 | 871 |
| 2020-06-09 | 884 |
| 2020-06-10 | 896 |
| 2020-06-11 | 930 |
| 2020-06-12 | 933 |
| 2020-06-13 | 956 |
| 2020-06-14 | 968 |
| 2020-06-15 | 980 |
| 2020-06-16 | 1000 |
| 2020-06-17 | 1023 |
| 2020-06-18 | 1046 |
| 2020-06-19 | 1071 |
| 2020-06-20 | 1111 |
| 2020-06-21 | 1141 |
| 2020-06-22 | 1172 |
| 2020-06-23 | 1245 |
| 2020-06-24 | 1350 |
| 2020-06-25 | 1446 |
| 2020-06-26 | 1525 |
| 2020-06-27 | 1650 |
| 2020-06-28 | 1751 |
| 2020-06-29 | 1876 |
| 2020-06-30 | 1952 |
| 2020-07-01 | 2075 |
| 2020-07-02 | 2192 |
| 2020-07-03 | 2302 |
| 2020-07-04 | 2381 |
| 2020-07-05 | 2475 |
| 2020-07-06 | 2510 |
| 2020-07-07 | 2597 |
| 2020-07-08 | 2652 |
| 2020-07-09 | 2771 |
| 2020-07-10 | 2889 |
| 2020-07-11 | 2995 |
| 2020-07-12 | 3071 |
| 2020-07-13 | 3151 |
| 2020-07-14 | 3250 |
| 2020-07-15 | 3298 |
| 2020-07-16 | 3376 |
| 2020-07-17 | 3438 |
| 2020-07-18 | 3495 |
| 2020-07-19 | 3529 |
| 2020-07-20 | 3562 |
| 2020-07-21 | 3623 |
| 2020-07-22 | 3666 |
| 2020-07-23 | 3729 |
| 2020-07-24 | 3776 |
| 2020-07-25 | 3825 |
| 2020-07-26 | 3870 |
| 2020-07-27 | 3904 |
| 2020-07-28 | 3970 |
| 2020-07-29 | 4007 |
| 2020-07-30 | 4086 |
| 2020-07-31 | 4112 |
| 2020-08-01 | 4198 |
| 2020-08-02 | 4230 |
| 2020-08-03 | 4263 |
| 2020-08-04 | 4340 |
| 2020-08-05 | 4376 |
| 2020-08-06 | 4436 |
| 2020-08-07 | 4492 |
| 2020-08-08 | 4536 |
| 2020-08-09 | 4554 |
| 2020-08-10 | 4578 |
| 2020-08-11 | 4630 |
| 2020-08-12 | 4666 |
| 2020-08-13 | 4749 |
| 2020-08-14 | 4783 |
| 2020-08-15 | 4849 |
| 2020-08-16 | 4874 |
| 2020-08-17 | 4890 |
| 2020-08-18 | 4924 |
| 2020-08-19 | 4955 |
| 2020-08-20 | 4998 |
| 2020-08-21 | 5053 |
| 2020-08-22 | 5088 |
| 2020-08-23 | 5117 |
| 2020-08-24 | 5161 |
| 2020-08-25 | 5209 |
| 2020-08-26 | 5294 |
| 2020-08-27 | 5330 |
| 2020-08-28 | 5367 |
| 2020-08-29 | 5426 |
| 2020-08-30 | 5429 |
| 2020-08-31 | 5520 |
| 2020-09-01 | 5564 |
| 2020-09-02 | 5639 |
| 2020-09-03 | 5799 |
| 2020-09-04 | 5961 |
| 2020-09-05 | 6118 |
| 2020-09-06 | 6208 |
| 2020-09-07 | 6427 |
| 2020-09-08 | 6489 |
| 2020-09-09 | 6971 |
| 2020-09-10 | 7254 |
| 2020-09-11 | 7389 |
| 2020-09-13 | 7846 |
| 2020-09-14 | 7896 |
| 2020-09-15 | 7984 |
| 2020-09-16 | 8242 |
| 2020-09-17 | 8455 |
| 2020-09-18 | 8635 |
| 2020-09-19 | 8815 |
| 2020-09-20 | 8895 |
| 2020-09-21 | 8947 |
| 2020-09-22 | 9040 |
| 2020-09-23 | 9203 |
| 2020-09-24 | 9345 |
| 2020-09-25 | 9488 |
| 2020-09-26 | 9607 |
| 2020-09-27 | 9763 |
| 2020-09-28 | 9857 |
| 2020-09-29 | 10129 |
| 2020-09-30 | 10201 |

(b) Milwaukee

Cumulative number of confiemed cases

|  | Milwaukee actual confirmed cases | Alagoz model results |
| --- | --- | --- |
| 2020-03-13 | 8 | 38 |
| 2020-03-14 | 10 | 51 |
| 2020-03-15 | 16 | 67 |
| 2020-03-16 | 28 | 88 |
| 2020-03-17 | 54 | 110 |
| 2020-03-18 | 79 | 140 |
| 2020-03-19 | 110 | 173 |
| 2020-03-20 | 160 | 212 |
| 2020-03-21 | 239 | 254 |
| 2020-03-22 | 283 | 302 |
| 2020-03-23 | 300 | 352 |
| 2020-03-24 | 373 | 404 |
| 2020-03-25 | 450 | 457 |
| 2020-03-26 | 529 | 509 |
| 2020-03-27 | 625 | 568 |
| 2020-03-28 | 713 | 631 |
| 2020-03-29 | 780 | 697 |
| 2020-03-30 | 858 | 769 |
| 2020-03-31 | 977 | 847 |
| 2020-04-01 | 1082 | 925 |
| 2020-04-02 | 1186 | 1007 |
| 2020-04-03 | 1324 | 1096 |
| 2020-04-04 | 1415 | 1185 |
| 2020-04-05 | 1535 | 1282 |
| 2020-04-06 | 1614 | 1382 |
| 2020-04-07 | 1723 | 1489 |
| 2020-04-08 | 1802 | 1597 |
| 2020-04-09 | 1906 | 1712 |
| 2020-04-10 | 1997 | 1836 |
| 2020-04-11 | 2062 | 1966 |
| 2020-04-12 | 2106 | 2037 |
| 2020-04-13 | 2165 | 2114 |
| 2020-04-14 | 2263 | 2191 |
| 2020-04-15 | 2348 | 2270 |
| 2020-04-16 | 2427 | 2350 |
| 2020-04-17 | 2485 | 2461 |
| 2020-04-18 | 2578 | 2567 |
| 2020-04-19 | 2626 | 2668 |
| 2020-04-20 | 2673 | 2766 |
| 2020-04-21 | 2750 | 2861 |
| 2020-04-22 | 2832 | 2950 |
| 2020-04-23 | 2891 | 3045 |
| 2020-04-24 | 2998 | 3140 |
| 2020-04-25 | 3113 | 3236 |
| 2020-04-26 | 3167 | 3333 |
| 2020-04-27 | 3225 | 3433 |
| 2020-04-28 | 3327 | 3531 |
| 2020-04-29 | 3462 | 3628 |
| 2020-04-30 | 3581 | 3726 |
| 2020-05-01 | 3692 | 3827 |
| 2020-05-02 | 3797 | 3924 |
| 2020-05-03 | 3861 | 4023 |
| 2020-05-04 | 3922 | 4123 |
| 2020-05-05 | 4045 | 4226 |
| 2020-05-06 | 4165 | 4325 |
| 2020-05-07 | 4324 | 4426 |
| 2020-05-08 | 4470 | 4524 |
| 2020-05-09 | 4574 | 4629 |
| 2020-05-10 | 4656 | 4745 |
| 2020-05-11 | 4709 | 4857 |
| 2020-05-12 | 4868 | 4974 |
| 2020-05-13 | 5069 | 5090 |
| 2020-05-14 | 5237 | 5208 |
| 2020-05-15 | 5486 | 5323 |
| 2020-05-16 | 5691 | 5438 |
| 2020-05-17 | 5758 | 5551 |
| 2020-05-18 | 5848 | 5701 |
| 2020-05-19 | 6180 | 5852 |
| 2020-05-20 | 6385 | 6002 |
| 2020-05-21 | 6583 | 6160 |
| 2020-05-22 | 6855 | 6324 |
| 2020-05-23 | 7076 | 6493 |
| 2020-05-24 | 7263 | 6675 |
| 2020-05-25 | 7442 | 6863 |
| 2020-05-26 | 7697 | 7062 |
| 2020-05-27 | 7991 | 7270 |
| 2020-05-28 | 8491 | 7492 |
| 2020-05-29 | 8754 | 7722 |
| 2020-05-30 | 8860 | 7906 |
| 2020-05-31 | 8930 | 8093 |
| 2020-06-01 | 9151 | 8291 |
| 2020-06-02 | 9283 | 8491 |
| 2020-06-03 | 9593 | 8694 |
| 2020-06-04 | 9774 | 8899 |
| 2020-06-05 | 9940 | 9104 |
| 2020-06-06 | 10081 | 9312 |
| 2020-06-07 | 10186 | 9518 |
| 2020-06-08 | 10267 | 9720 |
| 2020-06-09 | 10406 | 9918 |
| 2020-06-10 | 10557 | 10123 |
| 2020-06-11 | 10671 | 10330 |
| 2020-06-12 | 10792 | 10535 |
| 2020-06-13 | 10898 | 10738 |
| 2020-06-14 | 10990 | 10943 |
| 2020-06-15 | 11081 | 11151 |
| 2020-06-16 | 11190 | 11354 |
| 2020-06-17 | 11405 | 11557 |
| 2020-06-18 | 11529 | 11758 |
| 2020-06-19 | 11675 | 11964 |
| 2020-06-20 | 11798 | 12171 |
| 2020-06-21 | 11901 | 12375 |
| 2020-06-22 | 11960 | 12578 |
| 2020-06-23 | 12140 | 12786 |
| 2020-06-24 | 12290 | 12997 |
| 2020-06-25 | 12448 | 13204 |
| 2020-06-26 | 12664 | 13412 |
| 2020-06-27 | 12819 | 13615 |
| 2020-06-28 | 12948 | 13815 |
| 2020-06-29 | 13130 | 14024 |
| 2020-06-30 | 13338 | 14231 |
| 2020-07-01 | 13535 | 14440 |
| 2020-07-02 | 13748 | 14652 |
| 2020-07-03 | 14075 | 14860 |
| 2020-07-04 | 14302 | 15064 |
| 2020-07-05 | 14507 | 15271 |
| 2020-07-06 | 14684 | 15477 |
| 2020-07-07 | 14937 | 15685 |
| 2020-07-08 | 15272 | 15891 |
| 2020-07-09 | 15641 | 16098 |
| 2020-07-10 | 16076 | 16308 |
| 2020-07-11 | 16413 | 16518 |
| 2020-07-12 | 16627 | 16727 |
| 2020-07-13 | 17033 | 16936 |
| 2020-07-14 | 17407 | 17146 |
| 2020-07-15 | 17871 | 17355 |
| 2020-07-16 | 18280 | 17563 |
| 2020-07-17 | 18700 | 17774 |
| 2020-07-18 | 19123 | 17979 |
| 2020-07-19 | 19451 | 18187 |
| 2020-07-20 | 20019 | 18431 |
| 2020-07-21 | 20347 | 18677 |
| 2020-07-22 | 20900 | 18917 |
| 2020-07-23 | 21422 | 19160 |
| 2020-07-24 | 21858 | 19411 |
| 2020-07-25 | 22309 | 19667 |
| 2020-07-26 | 22565 | 19933 |
| 2020-07-27 | 22908 | 20201 |
| 2020-07-28 | 23294 | 20475 |
| 2020-07-29 | 23754 | 20757 |
| 2020-07-30 | 24069 | 21046 |
| 2020-07-31 | 24515 | 21343 |
| 2020-08-01 | 24838 | 21639 |
| 2020-08-02 | 24996 | 21954 |
| 2020-08-03 | 25273 | 22270 |
| 2020-08-04 | 25566 | 22596 |
| 2020-08-05 | 25910 | 22926 |
| 2020-08-06 | 26305 | 23265 |
| 2020-08-07 | 26823 | 23622 |
| 2020-08-08 | 27054 | 23977 |
| 2020-08-09 | 27235 | 24345 |
| 2020-08-10 | 27509 | 24720 |
| 2020-08-11 | 27636 | 25106 |
| 2020-08-12 | 28065 | 25498 |
| 2020-08-13 | 28481 | 25899 |
| 2020-08-14 | 28777 | 26306 |
| 2020-08-15 | 29064 | 26718 |
| 2020-08-16 | 29231 | 27146 |
| 2020-08-17 | 29438 | 27590 |
| 2020-08-18 | 29689 | 28034 |
| 2020-08-19 | 30022 | 28423 |
| 2020-08-20 | 30331 | 28826 |
| 2020-08-21 | 30638 | 29237 |
| 2020-08-22 | 30733 | 29639 |
| 2020-08-23 | 30861 | 30053 |
| 2020-08-24 | 31030 | 30465 |
| 2020-08-25 | 31258 | 30875 |
| 2020-08-26 | 31520 | 31282 |
| 2020-08-27 | 31774 | 31692 |
| 2020-08-28 | 31994 | 32099 |
| 2020-08-29 | 32136 | 32496 |
| 2020-08-30 | 32178 | 32896 |
| 2020-08-31 | 32378 | 33298 |
| 2020-09-01 | 32517 | 33703 |
| 2020-09-02 | 32689 | 34102 |
| 2020-09-03 | 33066 | 34501 |
| 2020-09-04 | 33244 | 34901 |
| 2020-09-05 | 33394 | 35299 |
| 2020-09-06 | 33472 | 35686 |
| 2020-09-07 | 33652 | 36076 |
| 2020-09-08 | 33756 | 36478 |
| 2020-09-09 | 33983 | 36868 |
| 2020-09-10 | 34222 | 37253 |
| 2020-09-11 | 34481 | 37643 |
| 2020-09-12 | 34735 | 38027 |
| 2020-09-13 | 34824 | 38416 |
| 2020-09-14 | 35019 | 38801 |
| 2020-09-15 | 35299 | 39186 |
| 2020-09-16 | 35674 | 39572 |
| 2020-09-17 | 36183 | 39952 |
| 2020-09-18 | 36595 | 40329 |
| 2020-09-19 | 36950 | 40706 |
| 2020-09-20 | 37122 | 41081 |
| 2020-09-21 | 37538 | 41457 |
| 2020-09-22 | 37833 | 41836 |
| 2020-09-23 | 38343 | 42209 |
| 2020-09-24 | 38813 | 42579 |
| 2020-09-25 | 39377 | 42950 |
| 2020-09-26 | 39854 | 43329 |
| 2020-09-27 | 40049 | 43696 |
| 2020-09-28 | 40510 | 44061 |

|  | Validation begin date |
| --- | --- |
| 2021-01-04 | 100000 |

|  | Set Parameters |
| --- | --- |
| 2020-12-13 | 1000000 |

(c) NYC

Cumulative number of confiemed cases

|  | NYC- actual confirmed cases | Alagoz model results |
| --- | --- | --- |
| 2020-03-13 | 137 | 970 |
| 2020-03-14 | 185 | 1416 |
| 2020-03-15 | 269 | 2088 |
| 2020-03-16 | 464 | 3019 |
| 2020-03-17 | 923 | 4209 |
| 2020-03-18 | 2009 | 5744 |
| 2020-03-19 | 3954 | 7623 |
| 2020-03-20 | 5683 | 9794 |
| 2020-03-21 | 8115 | 12226 |
| 2020-03-22 | 10764 | 14803 |
| 2020-03-23 | 13119 | 17237 |
| 2020-03-24 | 15597 | 19264 |
| 2020-03-25 | 17856 | 21702 |
| 2020-03-26 | 23112 | 24464 |
| 2020-03-27 | 26697 | 27356 |
| 2020-03-28 | 30765 | 30399 |
| 2020-03-29 | 33474 | 33612 |
| 2020-03-30 | 38087 | 36926 |
| 2020-03-31 | 41771 | 40383 |
| 2020-04-01 | 47439 | 43962 |
| 2020-04-02 | 49707 | 47704 |
| 2020-04-03 | 57160 | 51558 |
| 2020-04-04 | 60850 | 55724 |
| 2020-04-05 | 64955 | 60041 |
| 2020-04-06 | 68776 | 64551 |
| 2020-04-07 | 74601 | 69293 |
| 2020-04-08 | 80204 | 74174 |
| 2020-04-09 | 87725 | 79294 |
| 2020-04-10 | 94409 | 84677 |
| 2020-04-11 | 98715 | 90188 |
| 2020-04-12 | 104410 | 96006 |
| 2020-04-13 | 107263 | 102121 |
| 2020-04-14 | 111424 | 108476 |
| 2020-04-15 | 117565 | 115583 |
| 2020-04-16 | 122148 | 122960 |
| 2020-04-17 | 126368 | 126975 |
| 2020-04-18 | 129788 | 131186 |
| 2020-04-19 | 132467 | 135585 |
| 2020-04-20 | 134874 | 139848 |
| 2020-04-21 | 138435 | 143941 |
| 2020-04-22 | 141754 | 147885 |
| 2020-04-23 | 146139 | 151528 |
| 2020-04-24 | 150576 | 154909 |
| 2020-04-25 | 153204 | 158114 |
| 2020-04-26 | 156100 | 161007 |
| 2020-04-27 | 157713 | 163601 |
| 2020-04-28 | 159865 | 166148 |
| 2020-04-29 | 162212 | 168612 |
| 2020-04-30 | 164505 | 170889 |
| 2020-05-01 | 166883 | 173053 |
| 2020-05-02 | 168845 | 175078 |
| 2020-05-03 | 170534 | 176994 |
| 2020-05-04 | 171723 | 178782 |
| 2020-05-05 | 173288 | 180446 |
| 2020-05-06 | 174709 | 182009 |
| 2020-05-07 | 176086 | 183465 |
| 2020-05-08 | 177481 | 184859 |
| 2020-05-09 | 178766 | 186173 |
| 2020-05-10 | 183662 | 187546 |
| 2020-05-11 | 184319 | 188824 |
| 2020-05-12 | 185206 | 190036 |
| 2020-05-13 | 186293 | 191206 |
| 2020-05-14 | 187848 | 192290 |
| 2020-05-15 | 189031 | 193291 |
| 2020-05-16 | 190408 | 194217 |
| 2020-05-17 | 191073 | 195088 |
| 2020-05-18 | 191650 | 195887 |
| 2020-05-19 | 192374 | 196657 |
| 2020-05-20 | 192840 | 197411 |
| 2020-05-21 | 193951 | 198095 |
| 2020-05-22 | 194667 | 198759 |
| 2020-05-23 | 195452 | 199378 |
| 2020-05-24 | 196098 | 199977 |
| 2020-05-25 | 196623 | 200537 |
| 2020-05-26 | 197351 | 201048 |
| 2020-05-27 | 198255 | 201534 |
| 2020-05-28 | 199038 | 201998 |
| 2020-05-29 | 199893 | 202433 |
| 2020-05-30 | 200547 | 202856 |
| 2020-05-31 | 200830 | 203261 |
| 2020-06-01 | 201123 | 203643 |
| 2020-06-02 | 201806 | 204004 |
| 2020-06-03 | 202319 | 204348 |
| 2020-06-04 | 202829 | 204669 |
| 2020-06-05 | 203430 | 204976 |
| 2020-06-06 | 203819 | 205263 |
| 2020-06-07 | 204253 | 205540 |
| 2020-06-08 | 204576 | 205812 |
| 2020-06-09 | 205011 | 206067 |
| 2020-06-10 | 205405 | 206310 |
| 2020-06-11 | 205846 | 206544 |
| 2020-06-12 | 206322 | 206858 |
| 2020-06-13 | 206606 | 207155 |
| 2020-06-14 | 207049 | 207452 |
| 2020-06-15 | 207373 | 207741 |
| 2020-06-16 | 207821 | 208025 |
| 2020-06-17 | 208097 | 208306 |
| 2020-06-18 | 208455 | 208599 |
| 2020-06-19 | 208763 | 208884 |
| 2020-06-20 | 209313 | 209187 |
| 2020-06-21 | 209562 | 209502 |
| 2020-06-22 | 209893 | 209809 |
| 2020-06-23 | 210185 | 210126 |
| 2020-06-24 | 210523 | 210438 |
| 2020-06-25 | 210908 | 210748 |
| 2020-06-26 | 211229 | 211054 |
| 2020-06-27 | 211569 | 211376 |
| 2020-06-28 | 211893 | 211704 |
| 2020-06-29 | 212072 | 212042 |
| 2020-06-30 | 212412 | 212380 |
| 2020-07-01 | 212774 | 212716 |
| 2020-07-02 | 213212 | 213052 |
| 2020-07-03 | 213561 | 213393 |
| 2020-07-04 | 213852 | 213733 |
| 2020-07-05 | 214061 | 214082 |
| 2020-07-06 | 214371 | 214314 |
| 2020-07-07 | 214570 | 214561 |
| 2020-07-08 | 214952 | 214811 |
| 2020-07-09 | 215265 | 215052 |
| 2020-07-10 | 215598 | 215296 |
| 2020-07-11 | 215924 | 215526 |
| 2020-07-12 | 216199 | 215742 |
| 2020-07-13 | 216468 | 215945 |
| 2020-07-14 | 216834 | 216138 |
| 2020-07-15 | 217230 | 216333 |
| 2020-07-16 | 217562 | 216513 |
| 2020-07-17 | 217895 | 216692 |
| 2020-07-18 | 218159 | 216872 |
| 2020-07-19 | 218478 | 217036 |
| 2020-07-20 | 219030 | 217196 |
| 2020-07-21 | 219128 | 217359 |
| 2020-07-22 | 219489 | 217513 |
| 2020-07-23 | 219641 | 217660 |
| 2020-07-24 | 220172 | 217808 |
| 2020-07-25 | 220266 | 217947 |
| 2020-07-26 | 220483 | 218081 |
| 2020-07-27 | 220907 | 218210 |
| 2020-07-28 | 221220 | 218337 |
| 2020-07-29 | 221534 | 218458 |
| 2020-07-30 | 221822 | 218586 |
| 2020-07-31 | 222131 | 218706 |
| 2020-08-01 | 222330 | 218822 |
| 2020-08-02 | 222522 | 218940 |
| 2020-08-03 | 222840 | 219048 |
| 2020-08-04 | 223186 | 219156 |
| 2020-08-05 | 223473 | 219265 |
| 2020-08-06 | 223876 | 219368 |
| 2020-08-07 | 224117 | 219475 |
| 2020-08-08 | 224438 | 219578 |
| 2020-08-09 | 224684 | 219685 |
| 2020-08-10 | 224920 | 219786 |
| 2020-08-11 | 225284 | 219882 |
| 2020-08-12 | 225713 | 219976 |
| 2020-08-13 | 226043 | 220079 |
| 2020-08-14 | 226367 | 220175 |
| 2020-08-15 | 226756 | 220274 |
| 2020-08-16 | 226977 | 220371 |
| 2020-08-17 | 227158 | 220472 |
| 2020-08-18 | 227419 | 220571 |
| 2020-08-19 | 227724 | 220662 |
| 2020-08-20 | 227927 | 220827 |
| 2020-08-21 | 228144 | 220984 |
| 2020-08-22 | 228428 | 221135 |
| 2020-08-23 | 228617 | 221304 |
| 2020-08-24 | 228788 | 221486 |
| 2020-08-25 | 228971 | 221678 |
| 2020-08-26 | 229207 | 221879 |
| 2020-08-27 | 229518 | 222095 |
| 2020-08-28 | 229751 | 222327 |
| 2020-08-29 | 229980 | 222576 |
| 2020-08-30 | 230205 | 222839 |
| 2020-08-31 | 230496 | 223110 |
| 2020-09-01 | 230744 | 223385 |
| 2020-09-02 | 231068 | 223697 |
| 2020-09-03 | 231290 | 224019 |
| 2020-09-04 | 231594 | 224346 |
| 2020-09-05 |  | 224689 |
| 2020-09-06 | 231824 | 225068 |
| 2020-09-07 | 231594 | 225459 |
| 2020-09-08 | 232232 | 225875 |
| 2020-09-09 | 232710 | 226316 |
| 2020-09-10 | 232926 | 226777 |
| 2020-09-11 | 233216 | 227245 |
| 2020-09-12 |  | 227758 |
| 2020-09-13 | 233636 | 228292 |
| 2020-09-14 | 233972 | 228831 |
| 2020-09-15 | 234225 | 229421 |
| 2020-09-16 | 234500 | 230029 |
| 2020-09-17 | 234827 | 230662 |
| 2020-09-18 | 235461 | 231338 |
| 2020-09-19 | 235649 | 231876 |
| 2020-09-20 | 235969 | 232440 |
| 2020-09-21 | 236253 | 233016 |
| 2020-09-22 | 236462 | 233610 |
| 2020-09-23 | 236841 | 234211 |
| 2020-09-24 | 237227 | 234809 |
| 2020-09-25 | 237628 | 235405 |
| 2020-09-26 | 237971 | 236017 |
| 2020-09-27 | 238338 | 236615 |
| 2020-09-28 | 238733 | 237225 |

|  | Validation begin date |
| --- | --- |
| 2021-01-14 | 1000000 |

|  | Set Parameters |
| --- | --- |
| 2020-12-15 | 1000000 |

### Section C. Sensitivity Analyses

We conducted a sensitivity analysis on several parameters. In this appendix, we describe these experiments.

#### Section C.1 Sensitivity analysis on the drop in adherence to nonpharmaceutical interventions after vaccination

Our base-case assumes that there is a 20% drop in adherence to NPIs among vaccinated individuals regardless of whether they gain immunity or not after vaccination. In this sensitivity analysis, we did all experiments by assuming that there is no drop in adherence to NPIs.

Supplement Table 2. Controllable spread date and number of cases on December 31, 2021 for different vaccination capacity scenarios when there is no drop in adherence to nonpharmaceutical interventions after vaccination (vaccine effectiveness 90%, vaccine coverage 50%). Numbers in parentheses represent percent reduction relative to no vaccine.

| Dane County | | | | | | | | | | | | | |
| --- | --- | --- | --- | --- | --- | --- | --- | --- | --- | --- | --- | --- | --- |
|  | 75% Adherence | | 70% Adherence | | | | 65% Adherence | | | 60% Adherence | | Dynamic Adherence | |
| Vaccination capacity | Date | Number of cases | Date | | | Number of cases | Date | Number of cases | | Date | Number of cases | Date | Number of cases |
| No Vaccine | 27-Mar-2021 | 43,932 | 18-Jul-2021 | | | 51,784 | 17-Mar-2022 | 111,014 | | 29-Oct-2021 | 200,642 | After June 2022 | 79,285 |
| 0.05% | 22-Mar-2021 | 43,222  (2%) | 25-May-2021 | | | 48,016  (7%) | 20-Oct-2021 | 73,747  (34%) | | 14-Oct-2021 | 150,022  (25%) | 18-Mar-2022 | 72,088  (9%) |
| 0.1% | 18-Mar-2021 | 42,697  (3%) | 2-May-2021 | | | 46,110  (11%) | 4-Aug-2021 | 59,452  (46%) | | 17-Sep-2021 | 109,029  (46%) | 28-Oct-2021 | 64,654  (18%) |
| 0.25% | 12-Mar-2021 | 42,164  (4%) | 1-Apr-2021 | | | 43,374  (16%) | 4-May-2021 | 46,128  (58%) | | 16-Jun-2021 | 53,602  (73%) | 30-May-2021 | 46,579  (41%) |
| 0.5% | 27-Feb-2021 | 40,720  (7%) | 10-Mar-2021 | | | 41,514  (20%) | 26-Mar-2021 | 42,903  (61%) | | 13-Apr-2021 | 45,483  (77%) | 30-Mar-2021 | 42,547  (46%) |
| Milwaukee | | | | | | | | | | | | | |
|  | 75% Adherence | | | | 70% Adherence | | 65% Adherence | | | 60% Adherence | | Dynamic Adherence | |
| Vaccination capacity | Date | Number of cases | | Date | | Number of cases | Date | | Number of cases | Date | Number of cases | Date | Number of cases |
| No Vaccine | 23-Mar-2021 | 167,626 | | 12-Jun-2021 | | 185,343 | 26-Mar-2022 | | 309,809 | 18-Nov-2021 | 567,311 | After June 2022 | 271,692 |
| 0.05% | 18-Mar-2021 | 165,699  (1%) | | 7-May-2021 | | 177,194  (4%) | 19-Sep-2021 | | 230,229  (26%) | 20-Oct-2021 | 418,533  (26%) | 2-Feb-2022 | 252,269  (7%) |
| 0.1% | 14-Mar-2021 | 164,239  (2%) | | 21-Apr-2021 | | 172,714  (7%) | 11-Jul-2021 | | 201,894  (35%) | 10-Sep-2021 | 312,232  (45%) | 2-Oct-2021 | 213,120  (22%) |
| 0.25% | 6-Mar-2021 | 161,330  (4%) | | 27-Mar-2021 | | 165,781  (11%) | 28-Apr-2021 | | 175,693  (43%) | 7-Jun-2021 | 201,573  (64%) | 15-May-2021 | 173,455  (36%) |
| 0.5% | 26-Feb-2021 | 158,587  (5%) | | 8-Mar-2021 | | 160,720  (13%) | 21-Mar-2021 | | 164,264  (47%) | 7-Apr-2021 | 170,621  (70%) | 26-Mar-2021 | 163,338  (40%) |
| NYC | | | | | | | | | | | | | |
|  | 90% Adherence | | | | 85% Adherence | | 80% Adherence | | | 75% Adherence | | Dynamic Adherence | |
| Vaccination capacity | Date | Number of cases | | Date | | Number of cases | Date | Number of cases | | Date | Number of cases | Date | Number of cases |
| No Vaccine | 22-Mar-2021 | 699,352 | | 27-Aug-2021 | | 1,110,070 | 4-Sep-2021 | 3,307,090 | | 21-Jun-2021 | 4,999,100 | After June 2022 | 1,308,990 |
| 0.05% | 19-Mar-2021 | 686,290  (2%) | | 28-Jun-2021 | | 987,825  (11%) | 20-Aug-2021 | 2,765,350  (16%) | | 20-Jun-2021 | 4,562,030  (9%) | After June 2022 | 1,274,190  (3%) |
| 0.1% | 17-Mar-2021 | 674,662  (4%) | | 31-May-2021 | | 911,821  (18%) | 3-Aug-2021 | 2,290,640  (31%) | | 19-Jun-2021 | 4,120,280  (18%) | 16-Feb-2022 | 1,193,630  (9%) |
| 0.25% | 11-Mar-2021 | 647,683  (7%) | | 23-Apr-2021 | | 786,153  (29%) | 16-Jun-2021 | 1,394,020  (58%) | | 11-Jun-2021 | 2,843,430  (43%) | 31-Jul-2021 | 865,510  (34%) |
| 0.5% | 4-Mar-2021 | 616,204  (12%) | | 27-Mar-2021 | | 687,376  (38%) | 30-Apr-2021 | 888,980  (73%) | | 6-Jun-2021 | 1,463,240  (71%) | 19-May-2021 | 696,766  (47%) |

Supplement Table 3. Controllable spread date and number of cases on December 31, 2021 for different vaccination effectiveness scenarios when there is no drop in adherence to nonpharmaceutical interventions after vaccination (vaccine coverage 50%, vaccination capacity 0.01%). Numbers in parentheses represent percent reduction in the number of cases relative to no vaccine.

| Dane County | | | | | | | | | | |
| --- | --- | --- | --- | --- | --- | --- | --- | --- | --- | --- |
|  | 75% Adherence | | 70% Adherence | | 65% Adherence | | 60% Adherence | | Dynamic Adherence | |
| Vaccine effectiveness | Date | Number of cases | Date | Number of cases | Date | Number of cases | Date | Number of cases | Date | Number of cases |
| No Vaccine | 27-Mar-2021 | 43,932 | 18-Jul-2021 | 51,784 | 17-Mar-2022 | 111,014 | 29-Oct-2021 | 200,642 | After June 2022 | 79,285 |
| 50% | 21-Mar-2021 | 43,178  (2%) | 22-May-2021 | 47,778  (8%) | 16-Oct-2021 | 71,736  (35%) | 20-Oct-2021 | 145,366  (28%) | After June 2022 | 73,250  (8%) |
| 75% | 18-Mar-2021 | 42,884  (2%) | 6-May-2021 | 46,710  (10%) | 25-Aug-2021 | 63,188  (43%) | 3-Oct-2021 | 121,636  (39%) | 15-Feb-2022 | 70,676  (11%) |
| 90% | 18-Mar-2021 | 42,697  (3%) | 2-May-2021 | 46,110  (11%) | 4-Aug-2021 | 59,452  (46%) | 17-Sep-2021 | 109,029  (46%) | 28-Oct-2021 | 64,654  (18%) |
| Milwaukee | | | | | | | | | | |
|  | 75% Adherence | | 70% Adherence | | 65% Adherence | | 60% Adherence | | Dynamic Adherence | |
| Vaccine effectiveness | Date | Number of cases | Date | Number of cases | Date | Number of cases | Date | Number of cases | Date | Number of cases |
| No Vaccine | 23-Mar-2021 | 167,626 | 12-Jun-2021 | 185,343 | 26-Mar-2022 | 309,809 | 18-Nov-2021 | 567,311 | After June 2022 | 271,692 |
| 50% | 18-Mar-2021 | 165,603  (1%) | 5-May-2021 | 176,771  (5%) | 7-Sep-2021 | 226,100  (27%) | 29-Oct-2021 | 405,917  (28%) | After June 2022 | 260,551  (4%) |
| 75% | 16-Mar-2021 | 164,805  (2%) | 25-Apr-2021 | 174,158  (6%) | 29-Jul-2021 | 209,451  (32%) | 1-Oct-2021 | 343,125  (40%) | 24-Dec-2021 | 232,820  (14%) |
| 90% | 14-Mar-2021 | 164,239  (2%) | 21-Apr-2021 | 172,714  (7%) | 11-Jul-2021 | 201,894  (35%) | 10-Sep-2021 | 312,232  (45%) | 2-Oct-2021 | 213,120  (22%) |
| NYC | | | | | | | | | | |
|  | 90% Adherence | | 85% Adherence | | 80% Adherence | | 75% Adherence | | Dynamic Adherence | |
| Vaccine effectiveness | Date | Number of cases | Date | Number of cases | Date | Number of cases | Date | Number of cases | Date | Number of cases |
| No Vaccine | 22-Mar-2021 | 699,352 | 27-Aug-2021 | 1,110,070 | 4-Sep-2021 | 3,307,090 | 21-Jun-2021 | 4,999,100 | After June 2022 | 1,308,990 |
| 50% | 19-Mar-2021 | 685,428  (2%) | 25-Jun-2021 | 979,607  (12%) | 19-Aug-2021 | 2,711,210  (18%) | 20-Jun-2021 | 4,514,110  (10%) | After June 2022 | 1,272,390  (3%) |
| 75% | 17-Mar-2021 | 678,957  (3%) | 6-Jun-2021 | 936,081  (16%) | 11-Aug-2021 | 2,444,070  (26%) | 19-Jun-2021 | 4,269,560  (15%) | After June 2022 | 1,256,520  (4%) |
| 90% | 17-Mar-2021 | 674,662  (4%) | 31-May-2021 | 911,821  (18%) | 3-Aug-2021 | 2,290,640  (31%) | 19-Jun-2021 | 4,120,280  (18%) | 16-Feb-2022 | 1,193,630  (9%) |

Supplement Figure 2. Impact of vaccine coverage on the number of confirmed cases in different regions for two different scenarios of adherence to nonpharmaceutical interventions when there is no drop in adherence after vaccination. In this figure, we assumed that vaccine effectiveness is equal to 90%, vaccine capacity is 0.1%/day.

a. Dane, adherence to NPIs is fixed at a level of 70%

Total number of confirmed cases

|  | No Vaccine | 25% | 50% | 60% | 75% | 100% Vaccinated |
| --- | --- | --- | --- | --- | --- | --- |
| 2020-03-15 | 0 | 0 | 0 | 0 | 0 | 0 |
| 2020-03-16 | 0 | 0 | 0 | 0 | 0 | 0 |
| 2020-03-17 | 0 | 0 | 0 | 0 | 0 | 0 |
| 2020-03-18 | 0 | 0 | 0 | 0 | 0 | 0 |
| 2020-03-19 | 1 | 1 | 1 | 1 | 1 | 1 |
| 2020-03-20 | 3 | 3 | 3 | 3 | 3 | 3 |
| 2020-03-21 | 5 | 5 | 5 | 5 | 5 | 5 |
| 2020-03-22 | 9 | 9 | 9 | 9 | 9 | 9 |
| 2020-03-23 | 12 | 12 | 12 | 12 | 12 | 12 |
| 2020-03-24 | 17 | 17 | 17 | 17 | 17 | 17 |
| 2020-03-25 | 23 | 23 | 23 | 23 | 23 | 23 |
| 2020-03-26 | 30 | 30 | 30 | 30 | 30 | 30 |
| 2020-03-27 | 39 | 39 | 39 | 39 | 39 | 39 |
| 2020-03-28 | 51 | 51 | 51 | 51 | 51 | 51 |
| 2020-03-29 | 64 | 64 | 64 | 64 | 64 | 64 |
| 2020-03-30 | 79 | 79 | 79 | 79 | 79 | 79 |
| 2020-03-31 | 96 | 96 | 96 | 96 | 96 | 96 |
| 2020-04-01 | 116 | 116 | 116 | 116 | 116 | 116 |
| 2020-04-02 | 135 | 135 | 135 | 135 | 135 | 135 |
| 2020-04-03 | 155 | 155 | 155 | 155 | 155 | 155 |
| 2020-04-04 | 174 | 174 | 174 | 174 | 174 | 174 |
| 2020-04-05 | 193 | 193 | 193 | 193 | 193 | 193 |
| 2020-04-06 | 210 | 210 | 210 | 210 | 210 | 210 |
| 2020-04-07 | 225 | 225 | 225 | 225 | 225 | 225 |
| 2020-04-08 | 237 | 237 | 237 | 237 | 237 | 237 |
| 2020-04-09 | 244 | 244 | 244 | 244 | 244 | 244 |
| 2020-04-10 | 258 | 258 | 258 | 258 | 258 | 258 |
| 2020-04-11 | 272 | 272 | 272 | 272 | 272 | 272 |
| 2020-04-12 | 284 | 284 | 284 | 284 | 284 | 284 |
| 2020-04-13 | 295 | 295 | 295 | 295 | 295 | 295 |
| 2020-04-14 | 305 | 305 | 305 | 305 | 305 | 305 |
| 2020-04-15 | 314 | 314 | 314 | 314 | 314 | 314 |
| 2020-04-16 | 323 | 323 | 323 | 323 | 323 | 323 |
| 2020-04-17 | 332 | 332 | 332 | 332 | 332 | 332 |
| 2020-04-18 | 340 | 340 | 340 | 340 | 340 | 340 |
| 2020-04-19 | 348 | 348 | 348 | 348 | 348 | 348 |
| 2020-04-20 | 356 | 356 | 356 | 356 | 356 | 356 |
| 2020-04-21 | 363 | 363 | 363 | 363 | 363 | 363 |
| 2020-04-22 | 370 | 370 | 370 | 370 | 370 | 370 |
| 2020-04-23 | 377 | 377 | 377 | 377 | 377 | 377 |
| 2020-04-24 | 383 | 383 | 383 | 383 | 383 | 383 |
| 2020-04-25 | 390 | 390 | 390 | 390 | 390 | 390 |
| 2020-04-26 | 396 | 396 | 396 | 396 | 396 | 396 |
| 2020-04-27 | 402 | 402 | 402 | 402 | 402 | 402 |
| 2020-04-28 | 408 | 408 | 408 | 408 | 408 | 408 |
| 2020-04-29 | 414 | 414 | 414 | 414 | 414 | 414 |
| 2020-04-30 | 420 | 420 | 420 | 420 | 420 | 420 |
| 2020-05-01 | 425 | 425 | 425 | 425 | 425 | 425 |
| 2020-05-02 | 430 | 430 | 430 | 430 | 430 | 430 |
| 2020-05-03 | 435 | 435 | 435 | 435 | 435 | 435 |
| 2020-05-04 | 440 | 440 | 440 | 440 | 440 | 440 |
| 2020-05-05 | 445 | 445 | 445 | 445 | 445 | 445 |
| 2020-05-06 | 449 | 449 | 449 | 449 | 449 | 449 |
| 2020-05-07 | 454 | 454 | 454 | 454 | 454 | 454 |
| 2020-05-08 | 459 | 459 | 459 | 459 | 459 | 459 |
| 2020-05-09 | 463 | 463 | 463 | 463 | 463 | 463 |
| 2020-05-10 | 467 | 467 | 467 | 467 | 467 | 467 |
| 2020-05-11 | 472 | 472 | 472 | 472 | 472 | 472 |
| 2020-05-12 | 476 | 476 | 476 | 476 | 476 | 476 |
| 2020-05-13 | 479 | 479 | 479 | 479 | 479 | 479 |
| 2020-05-14 | 483 | 483 | 483 | 483 | 483 | 483 |
| 2020-05-15 | 487 | 487 | 487 | 487 | 487 | 487 |
| 2020-05-16 | 492 | 492 | 492 | 492 | 492 | 492 |
| 2020-05-17 | 497 | 497 | 497 | 497 | 497 | 497 |
| 2020-05-18 | 502 | 502 | 502 | 502 | 502 | 502 |
| 2020-05-19 | 507 | 507 | 507 | 507 | 507 | 507 |
| 2020-05-20 | 512 | 512 | 512 | 512 | 512 | 512 |
| 2020-05-21 | 519 | 519 | 519 | 519 | 519 | 519 |
| 2020-05-22 | 525 | 525 | 525 | 525 | 525 | 525 |
| 2020-05-23 | 531 | 531 | 531 | 531 | 531 | 531 |
| 2020-05-24 | 537 | 537 | 537 | 537 | 537 | 537 |
| 2020-05-25 | 544 | 544 | 544 | 544 | 544 | 544 |
| 2020-05-26 | 551 | 551 | 551 | 551 | 551 | 551 |
| 2020-05-27 | 558 | 558 | 558 | 558 | 558 | 558 |
| 2020-05-28 | 565 | 565 | 565 | 565 | 565 | 565 |
| 2020-05-29 | 574 | 574 | 574 | 574 | 574 | 574 |
| 2020-05-30 | 583 | 583 | 583 | 583 | 583 | 583 |
| 2020-05-31 | 593 | 593 | 593 | 593 | 593 | 593 |
| 2020-06-01 | 603 | 603 | 603 | 603 | 603 | 603 |
| 2020-06-02 | 614 | 614 | 614 | 614 | 614 | 614 |
| 2020-06-03 | 625 | 625 | 625 | 625 | 625 | 625 |
| 2020-06-04 | 636 | 636 | 636 | 636 | 636 | 636 |
| 2020-06-05 | 649 | 649 | 649 | 649 | 649 | 649 |
| 2020-06-06 | 662 | 662 | 662 | 662 | 662 | 662 |
| 2020-06-07 | 676 | 676 | 676 | 676 | 676 | 676 |
| 2020-06-08 | 691 | 691 | 691 | 691 | 691 | 691 |
| 2020-06-09 | 707 | 707 | 707 | 707 | 707 | 707 |
| 2020-06-10 | 723 | 723 | 723 | 723 | 723 | 723 |
| 2020-06-11 | 740 | 740 | 740 | 740 | 740 | 740 |
| 2020-06-12 | 759 | 759 | 759 | 759 | 759 | 759 |
| 2020-06-13 | 778 | 778 | 778 | 778 | 778 | 778 |
| 2020-06-14 | 797 | 797 | 797 | 797 | 797 | 797 |
| 2020-06-15 | 818 | 818 | 818 | 818 | 818 | 818 |
| 2020-06-16 | 840 | 840 | 840 | 840 | 840 | 840 |
| 2020-06-17 | 863 | 863 | 863 | 863 | 863 | 863 |
| 2020-06-18 | 887 | 887 | 887 | 887 | 887 | 887 |
| 2020-06-19 | 913 | 913 | 913 | 913 | 913 | 913 |
| 2020-06-20 | 940 | 940 | 940 | 940 | 940 | 940 |
| 2020-06-21 | 968 | 968 | 968 | 968 | 968 | 968 |
| 2020-06-22 | 997 | 997 | 997 | 997 | 997 | 997 |
| 2020-06-23 | 1028 | 1028 | 1028 | 1028 | 1028 | 1028 |
| 2020-06-24 | 1061 | 1061 | 1061 | 1061 | 1061 | 1061 |
| 2020-06-25 | 1096 | 1096 | 1096 | 1096 | 1096 | 1096 |
| 2020-06-26 | 1132 | 1132 | 1132 | 1132 | 1132 | 1132 |
| 2020-06-27 | 1169 | 1169 | 1169 | 1169 | 1169 | 1169 |
| 2020-06-28 | 1207 | 1207 | 1207 | 1207 | 1207 | 1207 |
| 2020-06-29 | 1249 | 1249 | 1249 | 1249 | 1249 | 1249 |
| 2020-06-30 | 1292 | 1292 | 1292 | 1292 | 1292 | 1292 |
| 2020-07-01 | 1338 | 1338 | 1338 | 1338 | 1338 | 1338 |
| 2020-07-02 | 1385 | 1385 | 1385 | 1385 | 1385 | 1385 |
| 2020-07-03 | 1434 | 1434 | 1434 | 1434 | 1434 | 1434 |
| 2020-07-04 | 1486 | 1486 | 1486 | 1486 | 1486 | 1486 |
| 2020-07-05 | 1541 | 1541 | 1541 | 1541 | 1541 | 1541 |
| 2020-07-06 | 1597 | 1597 | 1597 | 1597 | 1597 | 1597 |
| 2020-07-07 | 1657 | 1657 | 1657 | 1657 | 1657 | 1657 |
| 2020-07-08 | 1721 | 1721 | 1721 | 1721 | 1721 | 1721 |
| 2020-07-09 | 1787 | 1787 | 1787 | 1787 | 1787 | 1787 |
| 2020-07-10 | 1855 | 1855 | 1855 | 1855 | 1855 | 1855 |
| 2020-07-11 | 1927 | 1927 | 1927 | 1927 | 1927 | 1927 |
| 2020-07-12 | 2003 | 2003 | 2003 | 2003 | 2003 | 2003 |
| 2020-07-13 | 2082 | 2082 | 2082 | 2082 | 2082 | 2082 |
| 2020-07-14 | 2165 | 2165 | 2165 | 2165 | 2165 | 2165 |
| 2020-07-15 | 2251 | 2251 | 2251 | 2251 | 2251 | 2251 |
| 2020-07-16 | 2320 | 2320 | 2320 | 2320 | 2320 | 2320 |
| 2020-07-17 | 2392 | 2392 | 2392 | 2392 | 2392 | 2392 |
| 2020-07-18 | 2467 | 2467 | 2467 | 2467 | 2467 | 2467 |
| 2020-07-19 | 2541 | 2541 | 2541 | 2541 | 2541 | 2541 |
| 2020-07-20 | 2616 | 2616 | 2616 | 2616 | 2616 | 2616 |
| 2020-07-21 | 2692 | 2692 | 2692 | 2692 | 2692 | 2692 |
| 2020-07-22 | 2768 | 2768 | 2768 | 2768 | 2768 | 2768 |
| 2020-07-23 | 2846 | 2846 | 2846 | 2846 | 2846 | 2846 |
| 2020-07-24 | 2922 | 2922 | 2922 | 2922 | 2922 | 2922 |
| 2020-07-25 | 2999 | 2999 | 2999 | 2999 | 2999 | 2999 |
| 2020-07-26 | 3073 | 3073 | 3073 | 3073 | 3073 | 3073 |
| 2020-07-27 | 3147 | 3147 | 3147 | 3147 | 3147 | 3147 |
| 2020-07-28 | 3222 | 3222 | 3222 | 3222 | 3222 | 3222 |
| 2020-07-29 | 3299 | 3299 | 3299 | 3299 | 3299 | 3299 |
| 2020-07-30 | 3376 | 3376 | 3376 | 3376 | 3376 | 3376 |
| 2020-07-31 | 3453 | 3453 | 3453 | 3453 | 3453 | 3453 |
| 2020-08-01 | 3528 | 3528 | 3528 | 3528 | 3528 | 3528 |
| 2020-08-02 | 3605 | 3605 | 3605 | 3605 | 3605 | 3605 |
| 2020-08-03 | 3682 | 3682 | 3682 | 3682 | 3682 | 3682 |
| 2020-08-04 | 3757 | 3757 | 3757 | 3757 | 3757 | 3757 |
| 2020-08-05 | 3832 | 3832 | 3832 | 3832 | 3832 | 3832 |
| 2020-08-06 | 3908 | 3908 | 3908 | 3908 | 3908 | 3908 |
| 2020-08-07 | 3983 | 3983 | 3983 | 3983 | 3983 | 3983 |
| 2020-08-08 | 4060 | 4060 | 4060 | 4060 | 4060 | 4060 |
| 2020-08-09 | 4137 | 4137 | 4137 | 4137 | 4137 | 4137 |
| 2020-08-10 | 4213 | 4213 | 4213 | 4213 | 4213 | 4213 |
| 2020-08-11 | 4290 | 4290 | 4290 | 4290 | 4290 | 4290 |
| 2020-08-12 | 4364 | 4364 | 4364 | 4364 | 4364 | 4364 |
| 2020-08-13 | 4439 | 4439 | 4439 | 4439 | 4439 | 4439 |
| 2020-08-14 | 4516 | 4516 | 4516 | 4516 | 4516 | 4516 |
| 2020-08-15 | 4592 | 4592 | 4592 | 4592 | 4592 | 4592 |
| 2020-08-16 | 4669 | 4669 | 4669 | 4669 | 4669 | 4669 |
| 2020-08-17 | 4747 | 4747 | 4747 | 4747 | 4747 | 4747 |
| 2020-08-18 | 4823 | 4823 | 4823 | 4823 | 4823 | 4823 |
| 2020-08-19 | 4900 | 4900 | 4900 | 4900 | 4900 | 4900 |
| 2020-08-20 | 4977 | 4977 | 4977 | 4977 | 4977 | 4977 |
| 2020-08-21 | 5055 | 5055 | 5055 | 5055 | 5055 | 5055 |
| 2020-08-22 | 5132 | 5132 | 5132 | 5132 | 5132 | 5132 |
| 2020-08-23 | 5210 | 5210 | 5210 | 5210 | 5210 | 5210 |
| 2020-08-24 | 5288 | 5288 | 5288 | 5288 | 5288 | 5288 |
| 2020-08-25 | 5366 | 5366 | 5366 | 5366 | 5366 | 5366 |
| 2020-08-26 | 5443 | 5443 | 5443 | 5443 | 5443 | 5443 |
| 2020-08-27 | 5520 | 5520 | 5520 | 5520 | 5520 | 5520 |
| 2020-08-28 | 5597 | 5597 | 5597 | 5597 | 5597 | 5597 |
| 2020-08-29 | 5675 | 5675 | 5675 | 5675 | 5675 | 5675 |
| 2020-08-30 | 5751 | 5751 | 5751 | 5751 | 5751 | 5751 |
| 2020-08-31 | 5829 | 5829 | 5829 | 5829 | 5829 | 5829 |
| 2020-09-01 | 5906 | 5906 | 5906 | 5906 | 5906 | 5906 |
| 2020-09-02 | 5983 | 5983 | 5983 | 5983 | 5983 | 5983 |
| 2020-09-03 | 6061 | 6061 | 6061 | 6061 | 6061 | 6061 |
| 2020-09-04 | 6137 | 6137 | 6137 | 6137 | 6137 | 6137 |
| 2020-09-05 | 6213 | 6213 | 6213 | 6213 | 6213 | 6213 |
| 2020-09-06 | 6290 | 6290 | 6290 | 6290 | 6290 | 6290 |
| 2020-09-07 | 6368 | 6368 | 6368 | 6368 | 6368 | 6368 |
| 2020-09-08 | 6445 | 6445 | 6445 | 6445 | 6445 | 6445 |
| 2020-09-09 | 6523 | 6523 | 6523 | 6523 | 6523 | 6523 |
| 2020-09-10 | 6601 | 6601 | 6601 | 6601 | 6601 | 6601 |
| 2020-09-11 | 6676 | 6676 | 6676 | 6676 | 6676 | 6676 |
| 2020-09-12 | 6754 | 6754 | 6754 | 6754 | 6754 | 6754 |
| 2020-09-13 | 6831 | 6831 | 6831 | 6831 | 6831 | 6831 |
| 2020-09-14 | 6908 | 6908 | 6908 | 6908 | 6908 | 6908 |
| 2020-09-15 | 6985 | 6985 | 6985 | 6985 | 6985 | 6985 |
| 2020-09-16 | 7075 | 7075 | 7075 | 7075 | 7075 | 7075 |
| 2020-09-17 | 7164 | 7164 | 7164 | 7164 | 7164 | 7164 |
| 2020-09-18 | 7254 | 7254 | 7254 | 7254 | 7254 | 7254 |
| 2020-09-19 | 7346 | 7346 | 7346 | 7346 | 7346 | 7346 |
| 2020-09-20 | 7439 | 7439 | 7439 | 7439 | 7439 | 7439 |
| 2020-09-21 | 7533 | 7533 | 7533 | 7533 | 7533 | 7533 |
| 2020-09-22 | 7631 | 7631 | 7631 | 7631 | 7631 | 7631 |
| 2020-09-23 | 7731 | 7731 | 7731 | 7731 | 7731 | 7731 |
| 2020-09-24 | 7833 | 7833 | 7833 | 7833 | 7833 | 7833 |
| 2020-09-25 | 7940 | 7940 | 7940 | 7940 | 7940 | 7940 |
| 2020-09-26 | 8048 | 8048 | 8048 | 8048 | 8048 | 8048 |
| 2020-09-27 | 8157 | 8157 | 8157 | 8157 | 8157 | 8157 |
| 2020-09-28 | 8270 | 8270 | 8270 | 8270 | 8270 | 8270 |
| 2020-09-29 | 8386 | 8386 | 8386 | 8386 | 8386 | 8386 |
| 2020-09-30 | 8504 | 8504 | 8504 | 8504 | 8504 | 8504 |

b. Dane, adherence to NPIs is dynamic

Total number of confirmed cases

|  | No Vaccine | 25% | 50% | 60% | 75% | 100% Vaccinated |
| --- | --- | --- | --- | --- | --- | --- |
| 2020-03-15 | 0 | 0 | 0 | 0 | 0 | 0 |
| 2020-03-16 | 0 | 0 | 0 | 0 | 0 | 0 |
| 2020-03-17 | 0 | 0 | 0 | 0 | 0 | 0 |
| 2020-03-18 | 0 | 0 | 0 | 0 | 0 | 0 |
| 2020-03-19 | 1 | 1 | 1 | 1 | 1 | 1 |
| 2020-03-20 | 3 | 3 | 3 | 3 | 3 | 3 |
| 2020-03-21 | 5 | 5 | 5 | 5 | 5 | 5 |
| 2020-03-22 | 9 | 9 | 9 | 9 | 9 | 9 |
| 2020-03-23 | 12 | 12 | 12 | 12 | 12 | 12 |
| 2020-03-24 | 17 | 17 | 17 | 17 | 17 | 17 |
| 2020-03-25 | 23 | 23 | 23 | 23 | 23 | 23 |
| 2020-03-26 | 30 | 30 | 30 | 30 | 30 | 30 |
| 2020-03-27 | 39 | 39 | 39 | 39 | 39 | 39 |
| 2020-03-28 | 51 | 51 | 51 | 51 | 51 | 51 |
| 2020-03-29 | 64 | 64 | 64 | 64 | 64 | 64 |
| 2020-03-30 | 79 | 79 | 79 | 79 | 79 | 79 |
| 2020-03-31 | 96 | 96 | 96 | 96 | 96 | 96 |
| 2020-04-01 | 116 | 116 | 116 | 116 | 116 | 116 |
| 2020-04-02 | 135 | 135 | 135 | 135 | 135 | 135 |
| 2020-04-03 | 155 | 155 | 155 | 155 | 155 | 155 |
| 2020-04-04 | 174 | 174 | 174 | 174 | 174 | 174 |
| 2020-04-05 | 193 | 193 | 193 | 193 | 193 | 193 |
| 2020-04-06 | 210 | 210 | 210 | 210 | 210 | 210 |
| 2020-04-07 | 225 | 225 | 225 | 225 | 225 | 225 |
| 2020-04-08 | 237 | 237 | 237 | 237 | 237 | 237 |
| 2020-04-09 | 244 | 244 | 244 | 244 | 244 | 244 |
| 2020-04-10 | 258 | 258 | 258 | 258 | 258 | 258 |
| 2020-04-11 | 272 | 272 | 272 | 272 | 272 | 272 |
| 2020-04-12 | 284 | 284 | 284 | 284 | 284 | 284 |
| 2020-04-13 | 295 | 295 | 295 | 295 | 295 | 295 |
| 2020-04-14 | 305 | 305 | 305 | 305 | 305 | 305 |
| 2020-04-15 | 314 | 314 | 314 | 314 | 314 | 314 |
| 2020-04-16 | 323 | 323 | 323 | 323 | 323 | 323 |
| 2020-04-17 | 332 | 332 | 332 | 332 | 332 | 332 |
| 2020-04-18 | 340 | 340 | 340 | 340 | 340 | 340 |
| 2020-04-19 | 348 | 348 | 348 | 348 | 348 | 348 |
| 2020-04-20 | 356 | 356 | 356 | 356 | 356 | 356 |
| 2020-04-21 | 363 | 363 | 363 | 363 | 363 | 363 |
| 2020-04-22 | 370 | 370 | 370 | 370 | 370 | 370 |
| 2020-04-23 | 377 | 377 | 377 | 377 | 377 | 377 |
| 2020-04-24 | 383 | 383 | 383 | 383 | 383 | 383 |
| 2020-04-25 | 390 | 390 | 390 | 390 | 390 | 390 |
| 2020-04-26 | 396 | 396 | 396 | 396 | 396 | 396 |
| 2020-04-27 | 402 | 402 | 402 | 402 | 402 | 402 |
| 2020-04-28 | 408 | 408 | 408 | 408 | 408 | 408 |
| 2020-04-29 | 414 | 414 | 414 | 414 | 414 | 414 |
| 2020-04-30 | 420 | 420 | 420 | 420 | 420 | 420 |
| 2020-05-01 | 425 | 425 | 425 | 425 | 425 | 425 |
| 2020-05-02 | 430 | 430 | 430 | 430 | 430 | 430 |
| 2020-05-03 | 435 | 435 | 435 | 435 | 435 | 435 |
| 2020-05-04 | 440 | 440 | 440 | 440 | 440 | 440 |
| 2020-05-05 | 445 | 445 | 445 | 445 | 445 | 445 |
| 2020-05-06 | 449 | 449 | 449 | 449 | 449 | 449 |
| 2020-05-07 | 454 | 454 | 454 | 454 | 454 | 454 |
| 2020-05-08 | 459 | 459 | 459 | 459 | 459 | 459 |
| 2020-05-09 | 463 | 463 | 463 | 463 | 463 | 463 |
| 2020-05-10 | 467 | 467 | 467 | 467 | 467 | 467 |
| 2020-05-11 | 472 | 472 | 472 | 472 | 472 | 472 |
| 2020-05-12 | 476 | 476 | 476 | 476 | 476 | 476 |
| 2020-05-13 | 479 | 479 | 479 | 479 | 479 | 479 |
| 2020-05-14 | 483 | 483 | 483 | 483 | 483 | 483 |
| 2020-05-15 | 487 | 487 | 487 | 487 | 487 | 487 |
| 2020-05-16 | 492 | 492 | 492 | 492 | 492 | 492 |
| 2020-05-17 | 497 | 497 | 497 | 497 | 497 | 497 |
| 2020-05-18 | 502 | 502 | 502 | 502 | 502 | 502 |
| 2020-05-19 | 507 | 507 | 507 | 507 | 507 | 507 |
| 2020-05-20 | 512 | 512 | 512 | 512 | 512 | 512 |
| 2020-05-21 | 519 | 519 | 519 | 519 | 519 | 519 |
| 2020-05-22 | 525 | 525 | 525 | 525 | 525 | 525 |
| 2020-05-23 | 531 | 531 | 531 | 531 | 531 | 531 |
| 2020-05-24 | 537 | 537 | 537 | 537 | 537 | 537 |
| 2020-05-25 | 544 | 544 | 544 | 544 | 544 | 544 |
| 2020-05-26 | 551 | 551 | 551 | 551 | 551 | 551 |
| 2020-05-27 | 558 | 558 | 558 | 558 | 558 | 558 |
| 2020-05-28 | 565 | 565 | 565 | 565 | 565 | 565 |
| 2020-05-29 | 574 | 574 | 574 | 574 | 574 | 574 |
| 2020-05-30 | 583 | 583 | 583 | 583 | 583 | 583 |
| 2020-05-31 | 593 | 593 | 593 | 593 | 593 | 593 |
| 2020-06-01 | 603 | 603 | 603 | 603 | 603 | 603 |
| 2020-06-02 | 614 | 614 | 614 | 614 | 614 | 614 |
| 2020-06-03 | 625 | 625 | 625 | 625 | 625 | 625 |
| 2020-06-04 | 636 | 636 | 636 | 636 | 636 | 636 |
| 2020-06-05 | 649 | 649 | 649 | 649 | 649 | 649 |
| 2020-06-06 | 662 | 662 | 662 | 662 | 662 | 662 |
| 2020-06-07 | 676 | 676 | 676 | 676 | 676 | 676 |
| 2020-06-08 | 691 | 691 | 691 | 691 | 691 | 691 |
| 2020-06-09 | 707 | 707 | 707 | 707 | 707 | 707 |
| 2020-06-10 | 723 | 723 | 723 | 723 | 723 | 723 |
| 2020-06-11 | 740 | 740 | 740 | 740 | 740 | 740 |
| 2020-06-12 | 759 | 759 | 759 | 759 | 759 | 759 |
| 2020-06-13 | 778 | 778 | 778 | 778 | 778 | 778 |
| 2020-06-14 | 797 | 797 | 797 | 797 | 797 | 797 |
| 2020-06-15 | 818 | 818 | 818 | 818 | 818 | 818 |
| 2020-06-16 | 840 | 840 | 840 | 840 | 840 | 840 |
| 2020-06-17 | 863 | 863 | 863 | 863 | 863 | 863 |
| 2020-06-18 | 887 | 887 | 887 | 887 | 887 | 887 |
| 2020-06-19 | 913 | 913 | 913 | 913 | 913 | 913 |
| 2020-06-20 | 940 | 940 | 940 | 940 | 940 | 940 |
| 2020-06-21 | 968 | 968 | 968 | 968 | 968 | 968 |
| 2020-06-22 | 997 | 997 | 997 | 997 | 997 | 997 |
| 2020-06-23 | 1028 | 1028 | 1028 | 1028 | 1028 | 1028 |
| 2020-06-24 | 1061 | 1061 | 1061 | 1061 | 1061 | 1061 |
| 2020-06-25 | 1096 | 1096 | 1096 | 1096 | 1096 | 1096 |
| 2020-06-26 | 1132 | 1132 | 1132 | 1132 | 1132 | 1132 |
| 2020-06-27 | 1169 | 1169 | 1169 | 1169 | 1169 | 1169 |
| 2020-06-28 | 1207 | 1207 | 1207 | 1207 | 1207 | 1207 |
| 2020-06-29 | 1249 | 1249 | 1249 | 1249 | 1249 | 1249 |
| 2020-06-30 | 1292 | 1292 | 1292 | 1292 | 1292 | 1292 |
| 2020-07-01 | 1338 | 1338 | 1338 | 1338 | 1338 | 1338 |
| 2020-07-02 | 1385 | 1385 | 1385 | 1385 | 1385 | 1385 |
| 2020-07-03 | 1434 | 1434 | 1434 | 1434 | 1434 | 1434 |
| 2020-07-04 | 1486 | 1486 | 1486 | 1486 | 1486 | 1486 |
| 2020-07-05 | 1541 | 1541 | 1541 | 1541 | 1541 | 1541 |
| 2020-07-06 | 1597 | 1597 | 1597 | 1597 | 1597 | 1597 |
| 2020-07-07 | 1657 | 1657 | 1657 | 1657 | 1657 | 1657 |
| 2020-07-08 | 1721 | 1721 | 1721 | 1721 | 1721 | 1721 |
| 2020-07-09 | 1787 | 1787 | 1787 | 1787 | 1787 | 1787 |
| 2020-07-10 | 1855 | 1855 | 1855 | 1855 | 1855 | 1855 |
| 2020-07-11 | 1927 | 1927 | 1927 | 1927 | 1927 | 1927 |
| 2020-07-12 | 2003 | 2003 | 2003 | 2003 | 2003 | 2003 |
| 2020-07-13 | 2082 | 2082 | 2082 | 2082 | 2082 | 2082 |
| 2020-07-14 | 2165 | 2165 | 2165 | 2165 | 2165 | 2165 |
| 2020-07-15 | 2251 | 2251 | 2251 | 2251 | 2251 | 2251 |
| 2020-07-16 | 2320 | 2320 | 2320 | 2320 | 2320 | 2320 |
| 2020-07-17 | 2392 | 2392 | 2392 | 2392 | 2392 | 2392 |
| 2020-07-18 | 2467 | 2467 | 2467 | 2467 | 2467 | 2467 |
| 2020-07-19 | 2541 | 2541 | 2541 | 2541 | 2541 | 2541 |
| 2020-07-20 | 2616 | 2616 | 2616 | 2616 | 2616 | 2616 |
| 2020-07-21 | 2692 | 2692 | 2692 | 2692 | 2692 | 2692 |
| 2020-07-22 | 2768 | 2768 | 2768 | 2768 | 2768 | 2768 |
| 2020-07-23 | 2846 | 2846 | 2846 | 2846 | 2846 | 2846 |
| 2020-07-24 | 2922 | 2922 | 2922 | 2922 | 2922 | 2922 |
| 2020-07-25 | 2999 | 2999 | 2999 | 2999 | 2999 | 2999 |
| 2020-07-26 | 3073 | 3073 | 3073 | 3073 | 3073 | 3073 |
| 2020-07-27 | 3147 | 3147 | 3147 | 3147 | 3147 | 3147 |
| 2020-07-28 | 3222 | 3222 | 3222 | 3222 | 3222 | 3222 |
| 2020-07-29 | 3299 | 3299 | 3299 | 3299 | 3299 | 3299 |
| 2020-07-30 | 3376 | 3376 | 3376 | 3376 | 3376 | 3376 |
| 2020-07-31 | 3453 | 3453 | 3453 | 3453 | 3453 | 3453 |
| 2020-08-01 | 3528 | 3528 | 3528 | 3528 | 3528 | 3528 |
| 2020-08-02 | 3605 | 3605 | 3605 | 3605 | 3605 | 3605 |
| 2020-08-03 | 3682 | 3682 | 3682 | 3682 | 3682 | 3682 |
| 2020-08-04 | 3757 | 3757 | 3757 | 3757 | 3757 | 3757 |
| 2020-08-05 | 3832 | 3832 | 3832 | 3832 | 3832 | 3832 |
| 2020-08-06 | 3908 | 3908 | 3908 | 3908 | 3908 | 3908 |
| 2020-08-07 | 3983 | 3983 | 3983 | 3983 | 3983 | 3983 |
| 2020-08-08 | 4060 | 4060 | 4060 | 4060 | 4060 | 4060 |
| 2020-08-09 | 4137 | 4137 | 4137 | 4137 | 4137 | 4137 |
| 2020-08-10 | 4213 | 4213 | 4213 | 4213 | 4213 | 4213 |
| 2020-08-11 | 4290 | 4290 | 4290 | 4290 | 4290 | 4290 |
| 2020-08-12 | 4364 | 4364 | 4364 | 4364 | 4364 | 4364 |
| 2020-08-13 | 4439 | 4439 | 4439 | 4439 | 4439 | 4439 |
| 2020-08-14 | 4516 | 4516 | 4516 | 4516 | 4516 | 4516 |
| 2020-08-15 | 4592 | 4592 | 4592 | 4592 | 4592 | 4592 |
| 2020-08-16 | 4669 | 4669 | 4669 | 4669 | 4669 | 4669 |
| 2020-08-17 | 4747 | 4747 | 4747 | 4747 | 4747 | 4747 |
| 2020-08-18 | 4823 | 4823 | 4823 | 4823 | 4823 | 4823 |
| 2020-08-19 | 4900 | 4900 | 4900 | 4900 | 4900 | 4900 |
| 2020-08-20 | 4977 | 4977 | 4977 | 4977 | 4977 | 4977 |
| 2020-08-21 | 5055 | 5055 | 5055 | 5055 | 5055 | 5055 |
| 2020-08-22 | 5132 | 5132 | 5132 | 5132 | 5132 | 5132 |
| 2020-08-23 | 5210 | 5210 | 5210 | 5210 | 5210 | 5210 |
| 2020-08-24 | 5288 | 5288 | 5288 | 5288 | 5288 | 5288 |
| 2020-08-25 | 5366 | 5366 | 5366 | 5366 | 5366 | 5366 |
| 2020-08-26 | 5443 | 5443 | 5443 | 5443 | 5443 | 5443 |
| 2020-08-27 | 5520 | 5520 | 5520 | 5520 | 5520 | 5520 |
| 2020-08-28 | 5597 | 5597 | 5597 | 5597 | 5597 | 5597 |
| 2020-08-29 | 5675 | 5675 | 5675 | 5675 | 5675 | 5675 |
| 2020-08-30 | 5751 | 5751 | 5751 | 5751 | 5751 | 5751 |
| 2020-08-31 | 5829 | 5829 | 5829 | 5829 | 5829 | 5829 |
| 2020-09-01 | 5906 | 5906 | 5906 | 5906 | 5906 | 5906 |
| 2020-09-02 | 5983 | 5983 | 5983 | 5983 | 5983 | 5983 |
| 2020-09-03 | 6061 | 6061 | 6061 | 6061 | 6061 | 6061 |
| 2020-09-04 | 6137 | 6137 | 6137 | 6137 | 6137 | 6137 |
| 2020-09-05 | 6213 | 6213 | 6213 | 6213 | 6213 | 6213 |
| 2020-09-06 | 6290 | 6290 | 6290 | 6290 | 6290 | 6290 |
| 2020-09-07 | 6368 | 6368 | 6368 | 6368 | 6368 | 6368 |
| 2020-09-08 | 6445 | 6445 | 6445 | 6445 | 6445 | 6445 |
| 2020-09-09 | 6523 | 6523 | 6523 | 6523 | 6523 | 6523 |
| 2020-09-10 | 6601 | 6601 | 6601 | 6601 | 6601 | 6601 |
| 2020-09-11 | 6676 | 6676 | 6676 | 6676 | 6676 | 6676 |
| 2020-09-12 | 6754 | 6754 | 6754 | 6754 | 6754 | 6754 |
| 2020-09-13 | 6831 | 6831 | 6831 | 6831 | 6831 | 6831 |
| 2020-09-14 | 6908 | 6908 | 6908 | 6908 | 6908 | 6908 |
| 2020-09-15 | 6985 | 6985 | 6985 | 6985 | 6985 | 6985 |
| 2020-09-16 | 7075 | 7075 | 7075 | 7075 | 7075 | 7075 |
| 2020-09-17 | 7164 | 7164 | 7164 | 7164 | 7164 | 7164 |
| 2020-09-18 | 7254 | 7254 | 7254 | 7254 | 7254 | 7254 |
| 2020-09-19 | 7346 | 7346 | 7346 | 7346 | 7346 | 7346 |
| 2020-09-20 | 7439 | 7439 | 7439 | 7439 | 7439 | 7439 |
| 2020-09-21 | 7533 | 7533 | 7533 | 7533 | 7533 | 7533 |
| 2020-09-22 | 7631 | 7631 | 7631 | 7631 | 7631 | 7631 |
| 2020-09-23 | 7731 | 7731 | 7731 | 7731 | 7731 | 7731 |
| 2020-09-24 | 7833 | 7833 | 7833 | 7833 | 7833 | 7833 |
| 2020-09-25 | 7940 | 7940 | 7940 | 7940 | 7940 | 7940 |
| 2020-09-26 | 8048 | 8048 | 8048 | 8048 | 8048 | 8048 |
| 2020-09-27 | 8157 | 8157 | 8157 | 8157 | 8157 | 8157 |
| 2020-09-28 | 8270 | 8270 | 8270 | 8270 | 8270 | 8270 |
| 2020-09-29 | 8386 | 8386 | 8386 | 8386 | 8386 | 8386 |
| 2020-09-30 | 8504 | 8504 | 8504 | 8504 | 8504 | 8504 |

c. Milwaukee, adherence to NPIs is fixed at a level of 70%

Total number of confirmed cases

|  | No Vaccine | 25% | 50% | 60% | 75% | 100% Vaccinated |
| --- | --- | --- | --- | --- | --- | --- |
| 2020-03-15 | 0 | 0 | 0 | 0 | 0 | 0 |
| 2020-03-16 | 0 | 0 | 0 | 0 | 0 | 0 |
| 2020-03-17 | 0 | 0 | 0 | 0 | 0 | 0 |
| 2020-03-18 | 0 | 0 | 0 | 0 | 0 | 0 |
| 2020-03-19 | 3 | 3 | 3 | 3 | 3 | 3 |
| 2020-03-20 | 7 | 7 | 7 | 7 | 7 | 7 |
| 2020-03-21 | 12 | 12 | 12 | 12 | 12 | 12 |
| 2020-03-22 | 20 | 20 | 20 | 20 | 20 | 20 |
| 2020-03-23 | 28 | 28 | 28 | 28 | 28 | 28 |
| 2020-03-24 | 38 | 38 | 38 | 38 | 38 | 38 |
| 2020-03-25 | 51 | 51 | 51 | 51 | 51 | 51 |
| 2020-03-26 | 67 | 67 | 67 | 67 | 67 | 67 |
| 2020-03-27 | 88 | 88 | 88 | 88 | 88 | 88 |
| 2020-03-28 | 110 | 110 | 110 | 110 | 110 | 110 |
| 2020-03-29 | 140 | 140 | 140 | 140 | 140 | 140 |
| 2020-03-30 | 173 | 173 | 173 | 173 | 173 | 173 |
| 2020-03-31 | 212 | 212 | 212 | 212 | 212 | 212 |
| 2020-04-01 | 254 | 254 | 254 | 254 | 254 | 254 |
| 2020-04-02 | 302 | 302 | 302 | 302 | 302 | 302 |
| 2020-04-03 | 352 | 352 | 352 | 352 | 352 | 352 |
| 2020-04-04 | 404 | 404 | 404 | 404 | 404 | 404 |
| 2020-04-05 | 457 | 457 | 457 | 457 | 457 | 457 |
| 2020-04-06 | 509 | 509 | 509 | 509 | 509 | 509 |
| 2020-04-07 | 568 | 568 | 568 | 568 | 568 | 568 |
| 2020-04-08 | 631 | 631 | 631 | 631 | 631 | 631 |
| 2020-04-09 | 697 | 697 | 697 | 697 | 697 | 697 |
| 2020-04-10 | 769 | 769 | 769 | 769 | 769 | 769 |
| 2020-04-11 | 847 | 847 | 847 | 847 | 847 | 847 |
| 2020-04-12 | 925 | 925 | 925 | 925 | 925 | 925 |
| 2020-04-13 | 1007 | 1007 | 1007 | 1007 | 1007 | 1007 |
| 2020-04-14 | 1096 | 1096 | 1096 | 1096 | 1096 | 1096 |
| 2020-04-15 | 1185 | 1185 | 1185 | 1185 | 1185 | 1185 |
| 2020-04-16 | 1282 | 1282 | 1282 | 1282 | 1282 | 1282 |
| 2020-04-17 | 1382 | 1382 | 1382 | 1382 | 1382 | 1382 |
| 2020-04-18 | 1489 | 1489 | 1489 | 1489 | 1489 | 1489 |
| 2020-04-19 | 1597 | 1597 | 1597 | 1597 | 1597 | 1597 |
| 2020-04-20 | 1712 | 1712 | 1712 | 1712 | 1712 | 1712 |
| 2020-04-21 | 1836 | 1836 | 1836 | 1836 | 1836 | 1836 |
| 2020-04-22 | 1966 | 1966 | 1966 | 1966 | 1966 | 1966 |
| 2020-04-23 | 2037 | 2037 | 2037 | 2037 | 2037 | 2037 |
| 2020-04-24 | 2114 | 2114 | 2114 | 2114 | 2114 | 2114 |
| 2020-04-25 | 2191 | 2191 | 2191 | 2191 | 2191 | 2191 |
| 2020-04-26 | 2270 | 2270 | 2270 | 2270 | 2270 | 2270 |
| 2020-04-27 | 2350 | 2350 | 2350 | 2350 | 2350 | 2350 |
| 2020-04-28 | 2461 | 2461 | 2461 | 2461 | 2461 | 2461 |
| 2020-04-29 | 2567 | 2567 | 2567 | 2567 | 2567 | 2567 |
| 2020-04-30 | 2668 | 2668 | 2668 | 2668 | 2668 | 2668 |
| 2020-05-01 | 2766 | 2766 | 2766 | 2766 | 2766 | 2766 |
| 2020-05-02 | 2861 | 2861 | 2861 | 2861 | 2861 | 2861 |
| 2020-05-03 | 2950 | 2950 | 2950 | 2950 | 2950 | 2950 |
| 2020-05-04 | 3045 | 3045 | 3045 | 3045 | 3045 | 3045 |
| 2020-05-05 | 3140 | 3140 | 3140 | 3140 | 3140 | 3140 |
| 2020-05-06 | 3236 | 3236 | 3236 | 3236 | 3236 | 3236 |
| 2020-05-07 | 3333 | 3333 | 3333 | 3333 | 3333 | 3333 |
| 2020-05-08 | 3433 | 3433 | 3433 | 3433 | 3433 | 3433 |
| 2020-05-09 | 3531 | 3531 | 3531 | 3531 | 3531 | 3531 |
| 2020-05-10 | 3628 | 3628 | 3628 | 3628 | 3628 | 3628 |
| 2020-05-11 | 3726 | 3726 | 3726 | 3726 | 3726 | 3726 |
| 2020-05-12 | 3827 | 3827 | 3827 | 3827 | 3827 | 3827 |
| 2020-05-13 | 3924 | 3924 | 3924 | 3924 | 3924 | 3924 |
| 2020-05-14 | 4023 | 4023 | 4023 | 4023 | 4023 | 4023 |
| 2020-05-15 | 4123 | 4123 | 4123 | 4123 | 4123 | 4123 |
| 2020-05-16 | 4226 | 4226 | 4226 | 4226 | 4226 | 4226 |
| 2020-05-17 | 4325 | 4325 | 4325 | 4325 | 4325 | 4325 |
| 2020-05-18 | 4426 | 4426 | 4426 | 4426 | 4426 | 4426 |
| 2020-05-19 | 4524 | 4524 | 4524 | 4524 | 4524 | 4524 |
| 2020-05-20 | 4629 | 4629 | 4629 | 4629 | 4629 | 4629 |
| 2020-05-21 | 4745 | 4745 | 4745 | 4745 | 4745 | 4745 |
| 2020-05-22 | 4857 | 4857 | 4857 | 4857 | 4857 | 4857 |
| 2020-05-23 | 4974 | 4974 | 4974 | 4974 | 4974 | 4974 |
| 2020-05-24 | 5090 | 5090 | 5090 | 5090 | 5090 | 5090 |
| 2020-05-25 | 5208 | 5208 | 5208 | 5208 | 5208 | 5208 |
| 2020-05-26 | 5323 | 5323 | 5323 | 5323 | 5323 | 5323 |
| 2020-05-27 | 5438 | 5438 | 5438 | 5438 | 5438 | 5438 |
| 2020-05-28 | 5551 | 5551 | 5551 | 5551 | 5551 | 5551 |
| 2020-05-29 | 5701 | 5701 | 5701 | 5701 | 5701 | 5701 |
| 2020-05-30 | 5852 | 5852 | 5852 | 5852 | 5852 | 5852 |
| 2020-05-31 | 6002 | 6002 | 6002 | 6002 | 6002 | 6002 |
| 2020-06-01 | 6160 | 6160 | 6160 | 6160 | 6160 | 6160 |
| 2020-06-02 | 6324 | 6324 | 6324 | 6324 | 6324 | 6324 |
| 2020-06-03 | 6493 | 6493 | 6493 | 6493 | 6493 | 6493 |
| 2020-06-04 | 6675 | 6675 | 6675 | 6675 | 6675 | 6675 |
| 2020-06-05 | 6863 | 6863 | 6863 | 6863 | 6863 | 6863 |
| 2020-06-06 | 7062 | 7062 | 7062 | 7062 | 7062 | 7062 |
| 2020-06-07 | 7270 | 7270 | 7270 | 7270 | 7270 | 7270 |
| 2020-06-08 | 7492 | 7492 | 7492 | 7492 | 7492 | 7492 |
| 2020-06-09 | 7722 | 7722 | 7722 | 7722 | 7722 | 7722 |
| 2020-06-10 | 7906 | 7906 | 7906 | 7906 | 7906 | 7906 |
| 2020-06-11 | 8093 | 8093 | 8093 | 8093 | 8093 | 8093 |
| 2020-06-12 | 8291 | 8291 | 8291 | 8291 | 8291 | 8291 |
| 2020-06-13 | 8491 | 8491 | 8491 | 8491 | 8491 | 8491 |
| 2020-06-14 | 8694 | 8694 | 8694 | 8694 | 8694 | 8694 |
| 2020-06-15 | 8899 | 8899 | 8899 | 8899 | 8899 | 8899 |
| 2020-06-16 | 9104 | 9104 | 9104 | 9104 | 9104 | 9104 |
| 2020-06-17 | 9312 | 9312 | 9312 | 9312 | 9312 | 9312 |
| 2020-06-18 | 9518 | 9518 | 9518 | 9518 | 9518 | 9518 |
| 2020-06-19 | 9720 | 9720 | 9720 | 9720 | 9720 | 9720 |
| 2020-06-20 | 9918 | 9918 | 9918 | 9918 | 9918 | 9918 |
| 2020-06-21 | 10123 | 10123 | 10123 | 10123 | 10123 | 10123 |
| 2020-06-22 | 10330 | 10330 | 10330 | 10330 | 10330 | 10330 |
| 2020-06-23 | 10535 | 10535 | 10535 | 10535 | 10535 | 10535 |
| 2020-06-24 | 10738 | 10738 | 10738 | 10738 | 10738 | 10738 |
| 2020-06-25 | 10943 | 10943 | 10943 | 10943 | 10943 | 10943 |
| 2020-06-26 | 11151 | 11151 | 11151 | 11151 | 11151 | 11151 |
| 2020-06-27 | 11354 | 11354 | 11354 | 11354 | 11354 | 11354 |
| 2020-06-28 | 11557 | 11557 | 11557 | 11557 | 11557 | 11557 |
| 2020-06-29 | 11758 | 11758 | 11758 | 11758 | 11758 | 11758 |
| 2020-06-30 | 11964 | 11964 | 11964 | 11964 | 11964 | 11964 |
| 2020-07-01 | 12171 | 12171 | 12171 | 12171 | 12171 | 12171 |
| 2020-07-02 | 12375 | 12375 | 12375 | 12375 | 12375 | 12375 |
| 2020-07-03 | 12578 | 12578 | 12578 | 12578 | 12578 | 12578 |
| 2020-07-04 | 12786 | 12786 | 12786 | 12786 | 12786 | 12786 |
| 2020-07-05 | 12997 | 12997 | 12997 | 12997 | 12997 | 12997 |
| 2020-07-06 | 13204 | 13204 | 13204 | 13204 | 13204 | 13204 |
| 2020-07-07 | 13412 | 13412 | 13412 | 13412 | 13412 | 13412 |
| 2020-07-08 | 13615 | 13615 | 13615 | 13615 | 13615 | 13615 |
| 2020-07-09 | 13815 | 13815 | 13815 | 13815 | 13815 | 13815 |
| 2020-07-10 | 14024 | 14024 | 14024 | 14024 | 14024 | 14024 |
| 2020-07-11 | 14231 | 14231 | 14231 | 14231 | 14231 | 14231 |
| 2020-07-12 | 14440 | 14440 | 14440 | 14440 | 14440 | 14440 |
| 2020-07-13 | 14652 | 14652 | 14652 | 14652 | 14652 | 14652 |
| 2020-07-14 | 14860 | 14860 | 14860 | 14860 | 14860 | 14860 |
| 2020-07-15 | 15064 | 15064 | 15064 | 15064 | 15064 | 15064 |
| 2020-07-16 | 15271 | 15271 | 15271 | 15271 | 15271 | 15271 |
| 2020-07-17 | 15477 | 15477 | 15477 | 15477 | 15477 | 15477 |
| 2020-07-18 | 15685 | 15685 | 15685 | 15685 | 15685 | 15685 |
| 2020-07-19 | 15891 | 15891 | 15891 | 15891 | 15891 | 15891 |
| 2020-07-20 | 16098 | 16098 | 16098 | 16098 | 16098 | 16098 |
| 2020-07-21 | 16308 | 16308 | 16308 | 16308 | 16308 | 16308 |
| 2020-07-22 | 16518 | 16518 | 16518 | 16518 | 16518 | 16518 |
| 2020-07-23 | 16727 | 16727 | 16727 | 16727 | 16727 | 16727 |
| 2020-07-24 | 16936 | 16936 | 16936 | 16936 | 16936 | 16936 |
| 2020-07-25 | 17146 | 17146 | 17146 | 17146 | 17146 | 17146 |
| 2020-07-26 | 17355 | 17355 | 17355 | 17355 | 17355 | 17355 |
| 2020-07-27 | 17563 | 17563 | 17563 | 17563 | 17563 | 17563 |
| 2020-07-28 | 17774 | 17774 | 17774 | 17774 | 17774 | 17774 |
| 2020-07-29 | 17979 | 17979 | 17979 | 17979 | 17979 | 17979 |
| 2020-07-30 | 18187 | 18187 | 18187 | 18187 | 18187 | 18187 |
| 2020-07-31 | 18431 | 18431 | 18431 | 18431 | 18431 | 18431 |
| 2020-08-01 | 18677 | 18677 | 18677 | 18677 | 18677 | 18677 |
| 2020-08-02 | 18917 | 18917 | 18917 | 18917 | 18917 | 18917 |
| 2020-08-03 | 19160 | 19160 | 19160 | 19160 | 19160 | 19160 |
| 2020-08-04 | 19411 | 19411 | 19411 | 19411 | 19411 | 19411 |
| 2020-08-05 | 19667 | 19667 | 19667 | 19667 | 19667 | 19667 |
| 2020-08-06 | 19933 | 19933 | 19933 | 19933 | 19933 | 19933 |
| 2020-08-07 | 20201 | 20201 | 20201 | 20201 | 20201 | 20201 |
| 2020-08-08 | 20475 | 20475 | 20475 | 20475 | 20475 | 20475 |
| 2020-08-09 | 20757 | 20757 | 20757 | 20757 | 20757 | 20757 |
| 2020-08-10 | 21046 | 21046 | 21046 | 21046 | 21046 | 21046 |
| 2020-08-11 | 21343 | 21343 | 21343 | 21343 | 21343 | 21343 |
| 2020-08-12 | 21639 | 21639 | 21639 | 21639 | 21639 | 21639 |
| 2020-08-13 | 21954 | 21954 | 21954 | 21954 | 21954 | 21954 |
| 2020-08-14 | 22270 | 22270 | 22270 | 22270 | 22270 | 22270 |
| 2020-08-15 | 22596 | 22596 | 22596 | 22596 | 22596 | 22596 |
| 2020-08-16 | 22926 | 22926 | 22926 | 22926 | 22926 | 22926 |
| 2020-08-17 | 23265 | 23265 | 23265 | 23265 | 23265 | 23265 |
| 2020-08-18 | 23622 | 23622 | 23622 | 23622 | 23622 | 23622 |
| 2020-08-19 | 23977 | 23977 | 23977 | 23977 | 23977 | 23977 |
| 2020-08-20 | 24345 | 24345 | 24345 | 24345 | 24345 | 24345 |
| 2020-08-21 | 24720 | 24720 | 24720 | 24720 | 24720 | 24720 |
| 2020-08-22 | 25106 | 25106 | 25106 | 25106 | 25106 | 25106 |
| 2020-08-23 | 25498 | 25498 | 25498 | 25498 | 25498 | 25498 |
| 2020-08-24 | 25899 | 25899 | 25899 | 25899 | 25899 | 25899 |
| 2020-08-25 | 26306 | 26306 | 26306 | 26306 | 26306 | 26306 |
| 2020-08-26 | 26718 | 26718 | 26718 | 26718 | 26718 | 26718 |
| 2020-08-27 | 27146 | 27146 | 27146 | 27146 | 27146 | 27146 |
| 2020-08-28 | 27590 | 27590 | 27590 | 27590 | 27590 | 27590 |
| 2020-08-29 | 28034 | 28034 | 28034 | 28034 | 28034 | 28034 |
| 2020-08-30 | 28423 | 28423 | 28423 | 28423 | 28423 | 28423 |
| 2020-08-31 | 28826 | 28826 | 28826 | 28826 | 28826 | 28826 |
| 2020-09-01 | 29237 | 29237 | 29237 | 29237 | 29237 | 29237 |
| 2020-09-02 | 29639 | 29639 | 29639 | 29639 | 29639 | 29639 |
| 2020-09-03 | 30053 | 30053 | 30053 | 30053 | 30053 | 30053 |
| 2020-09-04 | 30465 | 30465 | 30465 | 30465 | 30465 | 30465 |
| 2020-09-05 | 30875 | 30875 | 30875 | 30875 | 30875 | 30875 |
| 2020-09-06 | 31282 | 31282 | 31282 | 31282 | 31282 | 31282 |
| 2020-09-07 | 31692 | 31692 | 31692 | 31692 | 31692 | 31692 |
| 2020-09-08 | 32099 | 32099 | 32099 | 32099 | 32099 | 32099 |
| 2020-09-09 | 32496 | 32496 | 32496 | 32496 | 32496 | 32496 |
| 2020-09-10 | 32896 | 32896 | 32896 | 32896 | 32896 | 32896 |
| 2020-09-11 | 33298 | 33298 | 33298 | 33298 | 33298 | 33298 |
| 2020-09-12 | 33703 | 33703 | 33703 | 33703 | 33703 | 33703 |
| 2020-09-13 | 34102 | 34102 | 34102 | 34102 | 34102 | 34102 |
| 2020-09-14 | 34501 | 34501 | 34501 | 34501 | 34501 | 34501 |
| 2020-09-15 | 34901 | 34901 | 34901 | 34901 | 34901 | 34901 |
| 2020-09-16 | 35299 | 35299 | 35299 | 35299 | 35299 | 35299 |
| 2020-09-17 | 35686 | 35686 | 35686 | 35686 | 35686 | 35686 |
| 2020-09-18 | 36076 | 36076 | 36076 | 36076 | 36076 | 36076 |
| 2020-09-19 | 36478 | 36478 | 36478 | 36478 | 36478 | 36478 |
| 2020-09-20 | 36868 | 36868 | 36868 | 36868 | 36868 | 36868 |
| 2020-09-21 | 37253 | 37253 | 37253 | 37253 | 37253 | 37253 |
| 2020-09-22 | 37643 | 37643 | 37643 | 37643 | 37643 | 37643 |
| 2020-09-23 | 38027 | 38027 | 38027 | 38027 | 38027 | 38027 |
| 2020-09-24 | 38416 | 38416 | 38416 | 38416 | 38416 | 38416 |
| 2020-09-25 | 38801 | 38801 | 38801 | 38801 | 38801 | 38801 |
| 2020-09-26 | 39186 | 39186 | 39186 | 39186 | 39186 | 39186 |
| 2020-09-27 | 39572 | 39572 | 39572 | 39572 | 39572 | 39572 |
| 2020-09-28 | 39952 | 39952 | 39952 | 39952 | 39952 | 39952 |
| 2020-09-29 | 40329 | 40329 | 40329 | 40329 | 40329 | 40329 |
| 2020-09-30 | 40706 | 40706 | 40706 | 40706 | 40706 | 40706 |

d. Milwaukee, adherence to NPIs is dynamic

Total number of confirmed cases

|  | No Vaccine | 25% | 50% | 60% | 75% | 100% Vaccinated |
| --- | --- | --- | --- | --- | --- | --- |
| 2020-03-15 | 0 | 0 | 0 | 0 | 0 | 0 |
| 2020-03-16 | 0 | 0 | 0 | 0 | 0 | 0 |
| 2020-03-17 | 0 | 0 | 0 | 0 | 0 | 0 |
| 2020-03-18 | 0 | 0 | 0 | 0 | 0 | 0 |
| 2020-03-19 | 3 | 3 | 3 | 3 | 3 | 3 |
| 2020-03-20 | 7 | 7 | 7 | 7 | 7 | 7 |
| 2020-03-21 | 12 | 12 | 12 | 12 | 12 | 12 |
| 2020-03-22 | 20 | 20 | 20 | 20 | 20 | 20 |
| 2020-03-23 | 28 | 28 | 28 | 28 | 28 | 28 |
| 2020-03-24 | 38 | 38 | 38 | 38 | 38 | 38 |
| 2020-03-25 | 51 | 51 | 51 | 51 | 51 | 51 |
| 2020-03-26 | 67 | 67 | 67 | 67 | 67 | 67 |
| 2020-03-27 | 88 | 88 | 88 | 88 | 88 | 88 |
| 2020-03-28 | 110 | 110 | 110 | 110 | 110 | 110 |
| 2020-03-29 | 140 | 140 | 140 | 140 | 140 | 140 |
| 2020-03-30 | 173 | 173 | 173 | 173 | 173 | 173 |
| 2020-03-31 | 212 | 212 | 212 | 212 | 212 | 212 |
| 2020-04-01 | 254 | 254 | 254 | 254 | 254 | 254 |
| 2020-04-02 | 302 | 302 | 302 | 302 | 302 | 302 |
| 2020-04-03 | 352 | 352 | 352 | 352 | 352 | 352 |
| 2020-04-04 | 404 | 404 | 404 | 404 | 404 | 404 |
| 2020-04-05 | 457 | 457 | 457 | 457 | 457 | 457 |
| 2020-04-06 | 509 | 509 | 509 | 509 | 509 | 509 |
| 2020-04-07 | 568 | 568 | 568 | 568 | 568 | 568 |
| 2020-04-08 | 631 | 631 | 631 | 631 | 631 | 631 |
| 2020-04-09 | 697 | 697 | 697 | 697 | 697 | 697 |
| 2020-04-10 | 769 | 769 | 769 | 769 | 769 | 769 |
| 2020-04-11 | 847 | 847 | 847 | 847 | 847 | 847 |
| 2020-04-12 | 925 | 925 | 925 | 925 | 925 | 925 |
| 2020-04-13 | 1007 | 1007 | 1007 | 1007 | 1007 | 1007 |
| 2020-04-14 | 1096 | 1096 | 1096 | 1096 | 1096 | 1096 |
| 2020-04-15 | 1185 | 1185 | 1185 | 1185 | 1185 | 1185 |
| 2020-04-16 | 1282 | 1282 | 1282 | 1282 | 1282 | 1282 |
| 2020-04-17 | 1382 | 1382 | 1382 | 1382 | 1382 | 1382 |
| 2020-04-18 | 1489 | 1489 | 1489 | 1489 | 1489 | 1489 |
| 2020-04-19 | 1597 | 1597 | 1597 | 1597 | 1597 | 1597 |
| 2020-04-20 | 1712 | 1712 | 1712 | 1712 | 1712 | 1712 |
| 2020-04-21 | 1836 | 1836 | 1836 | 1836 | 1836 | 1836 |
| 2020-04-22 | 1966 | 1966 | 1966 | 1966 | 1966 | 1966 |
| 2020-04-23 | 2037 | 2037 | 2037 | 2037 | 2037 | 2037 |
| 2020-04-24 | 2114 | 2114 | 2114 | 2114 | 2114 | 2114 |
| 2020-04-25 | 2191 | 2191 | 2191 | 2191 | 2191 | 2191 |
| 2020-04-26 | 2270 | 2270 | 2270 | 2270 | 2270 | 2270 |
| 2020-04-27 | 2350 | 2350 | 2350 | 2350 | 2350 | 2350 |
| 2020-04-28 | 2461 | 2461 | 2461 | 2461 | 2461 | 2461 |
| 2020-04-29 | 2567 | 2567 | 2567 | 2567 | 2567 | 2567 |
| 2020-04-30 | 2668 | 2668 | 2668 | 2668 | 2668 | 2668 |
| 2020-05-01 | 2766 | 2766 | 2766 | 2766 | 2766 | 2766 |
| 2020-05-02 | 2861 | 2861 | 2861 | 2861 | 2861 | 2861 |
| 2020-05-03 | 2950 | 2950 | 2950 | 2950 | 2950 | 2950 |
| 2020-05-04 | 3045 | 3045 | 3045 | 3045 | 3045 | 3045 |
| 2020-05-05 | 3140 | 3140 | 3140 | 3140 | 3140 | 3140 |
| 2020-05-06 | 3236 | 3236 | 3236 | 3236 | 3236 | 3236 |
| 2020-05-07 | 3333 | 3333 | 3333 | 3333 | 3333 | 3333 |
| 2020-05-08 | 3433 | 3433 | 3433 | 3433 | 3433 | 3433 |
| 2020-05-09 | 3531 | 3531 | 3531 | 3531 | 3531 | 3531 |
| 2020-05-10 | 3628 | 3628 | 3628 | 3628 | 3628 | 3628 |
| 2020-05-11 | 3726 | 3726 | 3726 | 3726 | 3726 | 3726 |
| 2020-05-12 | 3827 | 3827 | 3827 | 3827 | 3827 | 3827 |
| 2020-05-13 | 3924 | 3924 | 3924 | 3924 | 3924 | 3924 |
| 2020-05-14 | 4023 | 4023 | 4023 | 4023 | 4023 | 4023 |
| 2020-05-15 | 4123 | 4123 | 4123 | 4123 | 4123 | 4123 |
| 2020-05-16 | 4226 | 4226 | 4226 | 4226 | 4226 | 4226 |
| 2020-05-17 | 4325 | 4325 | 4325 | 4325 | 4325 | 4325 |
| 2020-05-18 | 4426 | 4426 | 4426 | 4426 | 4426 | 4426 |
| 2020-05-19 | 4524 | 4524 | 4524 | 4524 | 4524 | 4524 |
| 2020-05-20 | 4629 | 4629 | 4629 | 4629 | 4629 | 4629 |
| 2020-05-21 | 4745 | 4745 | 4745 | 4745 | 4745 | 4745 |
| 2020-05-22 | 4857 | 4857 | 4857 | 4857 | 4857 | 4857 |
| 2020-05-23 | 4974 | 4974 | 4974 | 4974 | 4974 | 4974 |
| 2020-05-24 | 5090 | 5090 | 5090 | 5090 | 5090 | 5090 |
| 2020-05-25 | 5208 | 5208 | 5208 | 5208 | 5208 | 5208 |
| 2020-05-26 | 5323 | 5323 | 5323 | 5323 | 5323 | 5323 |
| 2020-05-27 | 5438 | 5438 | 5438 | 5438 | 5438 | 5438 |
| 2020-05-28 | 5551 | 5551 | 5551 | 5551 | 5551 | 5551 |
| 2020-05-29 | 5701 | 5701 | 5701 | 5701 | 5701 | 5701 |
| 2020-05-30 | 5852 | 5852 | 5852 | 5852 | 5852 | 5852 |
| 2020-05-31 | 6002 | 6002 | 6002 | 6002 | 6002 | 6002 |
| 2020-06-01 | 6160 | 6160 | 6160 | 6160 | 6160 | 6160 |
| 2020-06-02 | 6324 | 6324 | 6324 | 6324 | 6324 | 6324 |
| 2020-06-03 | 6493 | 6493 | 6493 | 6493 | 6493 | 6493 |
| 2020-06-04 | 6675 | 6675 | 6675 | 6675 | 6675 | 6675 |
| 2020-06-05 | 6863 | 6863 | 6863 | 6863 | 6863 | 6863 |
| 2020-06-06 | 7062 | 7062 | 7062 | 7062 | 7062 | 7062 |
| 2020-06-07 | 7270 | 7270 | 7270 | 7270 | 7270 | 7270 |
| 2020-06-08 | 7492 | 7492 | 7492 | 7492 | 7492 | 7492 |
| 2020-06-09 | 7722 | 7722 | 7722 | 7722 | 7722 | 7722 |
| 2020-06-10 | 7906 | 7906 | 7906 | 7906 | 7906 | 7906 |
| 2020-06-11 | 8093 | 8093 | 8093 | 8093 | 8093 | 8093 |
| 2020-06-12 | 8291 | 8291 | 8291 | 8291 | 8291 | 8291 |
| 2020-06-13 | 8491 | 8491 | 8491 | 8491 | 8491 | 8491 |
| 2020-06-14 | 8694 | 8694 | 8694 | 8694 | 8694 | 8694 |
| 2020-06-15 | 8899 | 8899 | 8899 | 8899 | 8899 | 8899 |
| 2020-06-16 | 9104 | 9104 | 9104 | 9104 | 9104 | 9104 |
| 2020-06-17 | 9312 | 9312 | 9312 | 9312 | 9312 | 9312 |
| 2020-06-18 | 9518 | 9518 | 9518 | 9518 | 9518 | 9518 |
| 2020-06-19 | 9720 | 9720 | 9720 | 9720 | 9720 | 9720 |
| 2020-06-20 | 9918 | 9918 | 9918 | 9918 | 9918 | 9918 |
| 2020-06-21 | 10123 | 10123 | 10123 | 10123 | 10123 | 10123 |
| 2020-06-22 | 10330 | 10330 | 10330 | 10330 | 10330 | 10330 |
| 2020-06-23 | 10535 | 10535 | 10535 | 10535 | 10535 | 10535 |
| 2020-06-24 | 10738 | 10738 | 10738 | 10738 | 10738 | 10738 |
| 2020-06-25 | 10943 | 10943 | 10943 | 10943 | 10943 | 10943 |
| 2020-06-26 | 11151 | 11151 | 11151 | 11151 | 11151 | 11151 |
| 2020-06-27 | 11354 | 11354 | 11354 | 11354 | 11354 | 11354 |
| 2020-06-28 | 11557 | 11557 | 11557 | 11557 | 11557 | 11557 |
| 2020-06-29 | 11758 | 11758 | 11758 | 11758 | 11758 | 11758 |
| 2020-06-30 | 11964 | 11964 | 11964 | 11964 | 11964 | 11964 |
| 2020-07-01 | 12171 | 12171 | 12171 | 12171 | 12171 | 12171 |
| 2020-07-02 | 12375 | 12375 | 12375 | 12375 | 12375 | 12375 |
| 2020-07-03 | 12578 | 12578 | 12578 | 12578 | 12578 | 12578 |
| 2020-07-04 | 12786 | 12786 | 12786 | 12786 | 12786 | 12786 |
| 2020-07-05 | 12997 | 12997 | 12997 | 12997 | 12997 | 12997 |
| 2020-07-06 | 13204 | 13204 | 13204 | 13204 | 13204 | 13204 |
| 2020-07-07 | 13412 | 13412 | 13412 | 13412 | 13412 | 13412 |
| 2020-07-08 | 13615 | 13615 | 13615 | 13615 | 13615 | 13615 |
| 2020-07-09 | 13815 | 13815 | 13815 | 13815 | 13815 | 13815 |
| 2020-07-10 | 14024 | 14024 | 14024 | 14024 | 14024 | 14024 |
| 2020-07-11 | 14231 | 14231 | 14231 | 14231 | 14231 | 14231 |
| 2020-07-12 | 14440 | 14440 | 14440 | 14440 | 14440 | 14440 |
| 2020-07-13 | 14652 | 14652 | 14652 | 14652 | 14652 | 14652 |
| 2020-07-14 | 14860 | 14860 | 14860 | 14860 | 14860 | 14860 |
| 2020-07-15 | 15064 | 15064 | 15064 | 15064 | 15064 | 15064 |
| 2020-07-16 | 15271 | 15271 | 15271 | 15271 | 15271 | 15271 |
| 2020-07-17 | 15477 | 15477 | 15477 | 15477 | 15477 | 15477 |
| 2020-07-18 | 15685 | 15685 | 15685 | 15685 | 15685 | 15685 |
| 2020-07-19 | 15891 | 15891 | 15891 | 15891 | 15891 | 15891 |
| 2020-07-20 | 16098 | 16098 | 16098 | 16098 | 16098 | 16098 |
| 2020-07-21 | 16308 | 16308 | 16308 | 16308 | 16308 | 16308 |
| 2020-07-22 | 16518 | 16518 | 16518 | 16518 | 16518 | 16518 |
| 2020-07-23 | 16727 | 16727 | 16727 | 16727 | 16727 | 16727 |
| 2020-07-24 | 16936 | 16936 | 16936 | 16936 | 16936 | 16936 |
| 2020-07-25 | 17146 | 17146 | 17146 | 17146 | 17146 | 17146 |
| 2020-07-26 | 17355 | 17355 | 17355 | 17355 | 17355 | 17355 |
| 2020-07-27 | 17563 | 17563 | 17563 | 17563 | 17563 | 17563 |
| 2020-07-28 | 17774 | 17774 | 17774 | 17774 | 17774 | 17774 |
| 2020-07-29 | 17979 | 17979 | 17979 | 17979 | 17979 | 17979 |
| 2020-07-30 | 18187 | 18187 | 18187 | 18187 | 18187 | 18187 |
| 2020-07-31 | 18431 | 18431 | 18431 | 18431 | 18431 | 18431 |
| 2020-08-01 | 18677 | 18677 | 18677 | 18677 | 18677 | 18677 |
| 2020-08-02 | 18917 | 18917 | 18917 | 18917 | 18917 | 18917 |
| 2020-08-03 | 19160 | 19160 | 19160 | 19160 | 19160 | 19160 |
| 2020-08-04 | 19411 | 19411 | 19411 | 19411 | 19411 | 19411 |
| 2020-08-05 | 19667 | 19667 | 19667 | 19667 | 19667 | 19667 |
| 2020-08-06 | 19933 | 19933 | 19933 | 19933 | 19933 | 19933 |
| 2020-08-07 | 20201 | 20201 | 20201 | 20201 | 20201 | 20201 |
| 2020-08-08 | 20475 | 20475 | 20475 | 20475 | 20475 | 20475 |
| 2020-08-09 | 20757 | 20757 | 20757 | 20757 | 20757 | 20757 |
| 2020-08-10 | 21046 | 21046 | 21046 | 21046 | 21046 | 21046 |
| 2020-08-11 | 21343 | 21343 | 21343 | 21343 | 21343 | 21343 |
| 2020-08-12 | 21639 | 21639 | 21639 | 21639 | 21639 | 21639 |
| 2020-08-13 | 21954 | 21954 | 21954 | 21954 | 21954 | 21954 |
| 2020-08-14 | 22270 | 22270 | 22270 | 22270 | 22270 | 22270 |
| 2020-08-15 | 22596 | 22596 | 22596 | 22596 | 22596 | 22596 |
| 2020-08-16 | 22926 | 22926 | 22926 | 22926 | 22926 | 22926 |
| 2020-08-17 | 23265 | 23265 | 23265 | 23265 | 23265 | 23265 |
| 2020-08-18 | 23622 | 23622 | 23622 | 23622 | 23622 | 23622 |
| 2020-08-19 | 23977 | 23977 | 23977 | 23977 | 23977 | 23977 |
| 2020-08-20 | 24345 | 24345 | 24345 | 24345 | 24345 | 24345 |
| 2020-08-21 | 24720 | 24720 | 24720 | 24720 | 24720 | 24720 |
| 2020-08-22 | 25106 | 25106 | 25106 | 25106 | 25106 | 25106 |
| 2020-08-23 | 25498 | 25498 | 25498 | 25498 | 25498 | 25498 |
| 2020-08-24 | 25899 | 25899 | 25899 | 25899 | 25899 | 25899 |
| 2020-08-25 | 26306 | 26306 | 26306 | 26306 | 26306 | 26306 |
| 2020-08-26 | 26718 | 26718 | 26718 | 26718 | 26718 | 26718 |
| 2020-08-27 | 27146 | 27146 | 27146 | 27146 | 27146 | 27146 |
| 2020-08-28 | 27590 | 27590 | 27590 | 27590 | 27590 | 27590 |
| 2020-08-29 | 28034 | 28034 | 28034 | 28034 | 28034 | 28034 |
| 2020-08-30 | 28423 | 28423 | 28423 | 28423 | 28423 | 28423 |
| 2020-08-31 | 28826 | 28826 | 28826 | 28826 | 28826 | 28826 |
| 2020-09-01 | 29237 | 29237 | 29237 | 29237 | 29237 | 29237 |
| 2020-09-02 | 29639 | 29639 | 29639 | 29639 | 29639 | 29639 |
| 2020-09-03 | 30053 | 30053 | 30053 | 30053 | 30053 | 30053 |
| 2020-09-04 | 30465 | 30465 | 30465 | 30465 | 30465 | 30465 |
| 2020-09-05 | 30875 | 30875 | 30875 | 30875 | 30875 | 30875 |
| 2020-09-06 | 31282 | 31282 | 31282 | 31282 | 31282 | 31282 |
| 2020-09-07 | 31692 | 31692 | 31692 | 31692 | 31692 | 31692 |
| 2020-09-08 | 32099 | 32099 | 32099 | 32099 | 32099 | 32099 |
| 2020-09-09 | 32496 | 32496 | 32496 | 32496 | 32496 | 32496 |
| 2020-09-10 | 32896 | 32896 | 32896 | 32896 | 32896 | 32896 |
| 2020-09-11 | 33298 | 33298 | 33298 | 33298 | 33298 | 33298 |
| 2020-09-12 | 33703 | 33703 | 33703 | 33703 | 33703 | 33703 |
| 2020-09-13 | 34102 | 34102 | 34102 | 34102 | 34102 | 34102 |
| 2020-09-14 | 34501 | 34501 | 34501 | 34501 | 34501 | 34501 |
| 2020-09-15 | 34901 | 34901 | 34901 | 34901 | 34901 | 34901 |
| 2020-09-16 | 35299 | 35299 | 35299 | 35299 | 35299 | 35299 |
| 2020-09-17 | 35686 | 35686 | 35686 | 35686 | 35686 | 35686 |
| 2020-09-18 | 36076 | 36076 | 36076 | 36076 | 36076 | 36076 |
| 2020-09-19 | 36478 | 36478 | 36478 | 36478 | 36478 | 36478 |
| 2020-09-20 | 36868 | 36868 | 36868 | 36868 | 36868 | 36868 |
| 2020-09-21 | 37253 | 37253 | 37253 | 37253 | 37253 | 37253 |
| 2020-09-22 | 37643 | 37643 | 37643 | 37643 | 37643 | 37643 |
| 2020-09-23 | 38027 | 38027 | 38027 | 38027 | 38027 | 38027 |
| 2020-09-24 | 38416 | 38416 | 38416 | 38416 | 38416 | 38416 |
| 2020-09-25 | 38801 | 38801 | 38801 | 38801 | 38801 | 38801 |
| 2020-09-26 | 39186 | 39186 | 39186 | 39186 | 39186 | 39186 |
| 2020-09-27 | 39572 | 39572 | 39572 | 39572 | 39572 | 39572 |
| 2020-09-28 | 39952 | 39952 | 39952 | 39952 | 39952 | 39952 |
| 2020-09-29 | 40329 | 40329 | 40329 | 40329 | 40329 | 40329 |
| 2020-09-30 | 40706 | 40706 | 40706 | 40706 | 40706 | 40706 |

e. NYC, adherence to NPIs is fixed at a level of 85%

Total number of confirmed cases

|  | No Vaccine | 25% | 50% | 60% | 75% | 100% Vaccinated |
| --- | --- | --- | --- | --- | --- | --- |
| 2020-03-15 | 0 | 0 | 0 | 0 | 0 | 0 |
| 2020-03-16 | 0 | 0 | 0 | 0 | 0 | 0 |
| 2020-03-17 | 0 | 0 | 0 | 0 | 0 | 0 |
| 2020-03-18 | 0 | 0 | 0 | 0 | 0 | 0 |
| 2020-03-19 | 16 | 16 | 16 | 16 | 16 | 16 |
| 2020-03-20 | 136 | 136 | 136 | 136 | 136 | 136 |
| 2020-03-21 | 256 | 256 | 256 | 256 | 256 | 256 |
| 2020-03-22 | 398 | 398 | 398 | 398 | 398 | 398 |
| 2020-03-23 | 635 | 635 | 635 | 635 | 635 | 635 |
| 2020-03-24 | 970 | 970 | 970 | 970 | 970 | 970 |
| 2020-03-25 | 1416 | 1416 | 1416 | 1416 | 1416 | 1416 |
| 2020-03-26 | 2088 | 2088 | 2088 | 2088 | 2088 | 2088 |
| 2020-03-27 | 3019 | 3019 | 3019 | 3019 | 3019 | 3019 |
| 2020-03-28 | 4209 | 4209 | 4209 | 4209 | 4209 | 4209 |
| 2020-03-29 | 5744 | 5744 | 5744 | 5744 | 5744 | 5744 |
| 2020-03-30 | 7623 | 7623 | 7623 | 7623 | 7623 | 7623 |
| 2020-03-31 | 9794 | 9794 | 9794 | 9794 | 9794 | 9794 |
| 2020-04-01 | 12226 | 12226 | 12226 | 12226 | 12226 | 12226 |
| 2020-04-02 | 14803 | 14803 | 14803 | 14803 | 14803 | 14803 |
| 2020-04-03 | 17237 | 17237 | 17237 | 17237 | 17237 | 17237 |
| 2020-04-04 | 19264 | 19264 | 19264 | 19264 | 19264 | 19264 |
| 2020-04-05 | 21702 | 21702 | 21702 | 21702 | 21702 | 21702 |
| 2020-04-06 | 24464 | 24464 | 24464 | 24464 | 24464 | 24464 |
| 2020-04-07 | 27356 | 27356 | 27356 | 27356 | 27356 | 27356 |
| 2020-04-08 | 30399 | 30399 | 30399 | 30399 | 30399 | 30399 |
| 2020-04-09 | 33612 | 33612 | 33612 | 33612 | 33612 | 33612 |
| 2020-04-10 | 36926 | 36926 | 36926 | 36926 | 36926 | 36926 |
| 2020-04-11 | 40383 | 40383 | 40383 | 40383 | 40383 | 40383 |
| 2020-04-12 | 43962 | 43962 | 43962 | 43962 | 43962 | 43962 |
| 2020-04-13 | 47704 | 47704 | 47704 | 47704 | 47704 | 47704 |
| 2020-04-14 | 51558 | 51558 | 51558 | 51558 | 51558 | 51558 |
| 2020-04-15 | 55724 | 55724 | 55724 | 55724 | 55724 | 55724 |
| 2020-04-16 | 60041 | 60041 | 60041 | 60041 | 60041 | 60041 |
| 2020-04-17 | 64551 | 64551 | 64551 | 64551 | 64551 | 64551 |
| 2020-04-18 | 69293 | 69293 | 69293 | 69293 | 69293 | 69293 |
| 2020-04-19 | 74174 | 74174 | 74174 | 74174 | 74174 | 74174 |
| 2020-04-20 | 79294 | 79294 | 79294 | 79294 | 79294 | 79294 |
| 2020-04-21 | 84677 | 84677 | 84677 | 84677 | 84677 | 84677 |
| 2020-04-22 | 90188 | 90188 | 90188 | 90188 | 90188 | 90188 |
| 2020-04-23 | 96006 | 96006 | 96006 | 96006 | 96006 | 96006 |
| 2020-04-24 | 102121 | 102121 | 102121 | 102121 | 102121 | 102121 |
| 2020-04-25 | 108476 | 108476 | 108476 | 108476 | 108476 | 108476 |
| 2020-04-26 | 115583 | 115583 | 115583 | 115583 | 115583 | 115583 |
| 2020-04-27 | 122960 | 122960 | 122960 | 122960 | 122960 | 122960 |
| 2020-04-28 | 126975 | 126975 | 126975 | 126975 | 126975 | 126975 |
| 2020-04-29 | 131186 | 131186 | 131186 | 131186 | 131186 | 131186 |
| 2020-04-30 | 135585 | 135585 | 135585 | 135585 | 135585 | 135585 |
| 2020-05-01 | 139848 | 139848 | 139848 | 139848 | 139848 | 139848 |
| 2020-05-02 | 143941 | 143941 | 143941 | 143941 | 143941 | 143941 |
| 2020-05-03 | 147885 | 147885 | 147885 | 147885 | 147885 | 147885 |
| 2020-05-04 | 151528 | 151528 | 151528 | 151528 | 151528 | 151528 |
| 2020-05-05 | 154909 | 154909 | 154909 | 154909 | 154909 | 154909 |
| 2020-05-06 | 158114 | 158114 | 158114 | 158114 | 158114 | 158114 |
| 2020-05-07 | 161007 | 161007 | 161007 | 161007 | 161007 | 161007 |
| 2020-05-08 | 163601 | 163601 | 163601 | 163601 | 163601 | 163601 |
| 2020-05-09 | 166148 | 166148 | 166148 | 166148 | 166148 | 166148 |
| 2020-05-10 | 168612 | 168612 | 168612 | 168612 | 168612 | 168612 |
| 2020-05-11 | 170889 | 170889 | 170889 | 170889 | 170889 | 170889 |
| 2020-05-12 | 173053 | 173053 | 173053 | 173053 | 173053 | 173053 |
| 2020-05-13 | 175078 | 175078 | 175078 | 175078 | 175078 | 175078 |
| 2020-05-14 | 176994 | 176994 | 176994 | 176994 | 176994 | 176994 |
| 2020-05-15 | 178782 | 178782 | 178782 | 178782 | 178782 | 178782 |
| 2020-05-16 | 180446 | 180446 | 180446 | 180446 | 180446 | 180446 |
| 2020-05-17 | 182009 | 182009 | 182009 | 182009 | 182009 | 182009 |
| 2020-05-18 | 183465 | 183465 | 183465 | 183465 | 183465 | 183465 |
| 2020-05-19 | 184859 | 184859 | 184859 | 184859 | 184859 | 184859 |
| 2020-05-20 | 186173 | 186173 | 186173 | 186173 | 186173 | 186173 |
| 2020-05-21 | 187546 | 187546 | 187546 | 187546 | 187546 | 187546 |
| 2020-05-22 | 188824 | 188824 | 188824 | 188824 | 188824 | 188824 |
| 2020-05-23 | 190036 | 190036 | 190036 | 190036 | 190036 | 190036 |
| 2020-05-24 | 191206 | 191206 | 191206 | 191206 | 191206 | 191206 |
| 2020-05-25 | 192290 | 192290 | 192290 | 192290 | 192290 | 192290 |
| 2020-05-26 | 193291 | 193291 | 193291 | 193291 | 193291 | 193291 |
| 2020-05-27 | 194217 | 194217 | 194217 | 194217 | 194217 | 194217 |
| 2020-05-28 | 195088 | 195088 | 195088 | 195088 | 195088 | 195088 |
| 2020-05-29 | 195887 | 195887 | 195887 | 195887 | 195887 | 195887 |
| 2020-05-30 | 196657 | 196657 | 196657 | 196657 | 196657 | 196657 |
| 2020-05-31 | 197411 | 197411 | 197411 | 197411 | 197411 | 197411 |
| 2020-06-01 | 198095 | 198095 | 198095 | 198095 | 198095 | 198095 |
| 2020-06-02 | 198759 | 198759 | 198759 | 198759 | 198759 | 198759 |
| 2020-06-03 | 199378 | 199378 | 199378 | 199378 | 199378 | 199378 |
| 2020-06-04 | 199977 | 199977 | 199977 | 199977 | 199977 | 199977 |
| 2020-06-05 | 200537 | 200537 | 200537 | 200537 | 200537 | 200537 |
| 2020-06-06 | 201048 | 201048 | 201048 | 201048 | 201048 | 201048 |
| 2020-06-07 | 201534 | 201534 | 201534 | 201534 | 201534 | 201534 |
| 2020-06-08 | 201998 | 201998 | 201998 | 201998 | 201998 | 201998 |
| 2020-06-09 | 202433 | 202433 | 202433 | 202433 | 202433 | 202433 |
| 2020-06-10 | 202856 | 202856 | 202856 | 202856 | 202856 | 202856 |
| 2020-06-11 | 203261 | 203261 | 203261 | 203261 | 203261 | 203261 |
| 2020-06-12 | 203643 | 203643 | 203643 | 203643 | 203643 | 203643 |
| 2020-06-13 | 204004 | 204004 | 204004 | 204004 | 204004 | 204004 |
| 2020-06-14 | 204348 | 204348 | 204348 | 204348 | 204348 | 204348 |
| 2020-06-15 | 204669 | 204669 | 204669 | 204669 | 204669 | 204669 |
| 2020-06-16 | 204976 | 204976 | 204976 | 204976 | 204976 | 204976 |
| 2020-06-17 | 205263 | 205263 | 205263 | 205263 | 205263 | 205263 |
| 2020-06-18 | 205540 | 205540 | 205540 | 205540 | 205540 | 205540 |
| 2020-06-19 | 205812 | 205812 | 205812 | 205812 | 205812 | 205812 |
| 2020-06-20 | 206067 | 206067 | 206067 | 206067 | 206067 | 206067 |
| 2020-06-21 | 206310 | 206310 | 206310 | 206310 | 206310 | 206310 |
| 2020-06-22 | 206544 | 206544 | 206544 | 206544 | 206544 | 206544 |
| 2020-06-23 | 206858 | 206858 | 206858 | 206858 | 206858 | 206858 |
| 2020-06-24 | 207155 | 207155 | 207155 | 207155 | 207155 | 207155 |
| 2020-06-25 | 207452 | 207452 | 207452 | 207452 | 207452 | 207452 |
| 2020-06-26 | 207741 | 207741 | 207741 | 207741 | 207741 | 207741 |
| 2020-06-27 | 208025 | 208025 | 208025 | 208025 | 208025 | 208025 |
| 2020-06-28 | 208306 | 208306 | 208306 | 208306 | 208306 | 208306 |
| 2020-06-29 | 208599 | 208599 | 208599 | 208599 | 208599 | 208599 |
| 2020-06-30 | 208884 | 208884 | 208884 | 208884 | 208884 | 208884 |
| 2020-07-01 | 209187 | 209187 | 209187 | 209187 | 209187 | 209187 |
| 2020-07-02 | 209502 | 209502 | 209502 | 209502 | 209502 | 209502 |
| 2020-07-03 | 209809 | 209809 | 209809 | 209809 | 209809 | 209809 |
| 2020-07-04 | 210126 | 210126 | 210126 | 210126 | 210126 | 210126 |
| 2020-07-05 | 210438 | 210438 | 210438 | 210438 | 210438 | 210438 |
| 2020-07-06 | 210748 | 210748 | 210748 | 210748 | 210748 | 210748 |
| 2020-07-07 | 211054 | 211054 | 211054 | 211054 | 211054 | 211054 |
| 2020-07-08 | 211376 | 211376 | 211376 | 211376 | 211376 | 211376 |
| 2020-07-09 | 211704 | 211704 | 211704 | 211704 | 211704 | 211704 |
| 2020-07-10 | 212042 | 212042 | 212042 | 212042 | 212042 | 212042 |
| 2020-07-11 | 212380 | 212380 | 212380 | 212380 | 212380 | 212380 |
| 2020-07-12 | 212716 | 212716 | 212716 | 212716 | 212716 | 212716 |
| 2020-07-13 | 213052 | 213052 | 213052 | 213052 | 213052 | 213052 |
| 2020-07-14 | 213393 | 213393 | 213393 | 213393 | 213393 | 213393 |
| 2020-07-15 | 213733 | 213733 | 213733 | 213733 | 213733 | 213733 |
| 2020-07-16 | 214082 | 214082 | 214082 | 214082 | 214082 | 214082 |
| 2020-07-17 | 214314 | 214314 | 214314 | 214314 | 214314 | 214314 |
| 2020-07-18 | 214561 | 214561 | 214561 | 214561 | 214561 | 214561 |
| 2020-07-19 | 214811 | 214811 | 214811 | 214811 | 214811 | 214811 |
| 2020-07-20 | 215052 | 215052 | 215052 | 215052 | 215052 | 215052 |
| 2020-07-21 | 215296 | 215296 | 215296 | 215296 | 215296 | 215296 |
| 2020-07-22 | 215526 | 215526 | 215526 | 215526 | 215526 | 215526 |
| 2020-07-23 | 215742 | 215742 | 215742 | 215742 | 215742 | 215742 |
| 2020-07-24 | 215945 | 215945 | 215945 | 215945 | 215945 | 215945 |
| 2020-07-25 | 216138 | 216138 | 216138 | 216138 | 216138 | 216138 |
| 2020-07-26 | 216333 | 216333 | 216333 | 216333 | 216333 | 216333 |
| 2020-07-27 | 216513 | 216513 | 216513 | 216513 | 216513 | 216513 |
| 2020-07-28 | 216692 | 216692 | 216692 | 216692 | 216692 | 216692 |
| 2020-07-29 | 216872 | 216872 | 216872 | 216872 | 216872 | 216872 |
| 2020-07-30 | 217036 | 217036 | 217036 | 217036 | 217036 | 217036 |
| 2020-07-31 | 217196 | 217196 | 217196 | 217196 | 217196 | 217196 |
| 2020-08-01 | 217359 | 217359 | 217359 | 217359 | 217359 | 217359 |
| 2020-08-02 | 217513 | 217513 | 217513 | 217513 | 217513 | 217513 |
| 2020-08-03 | 217660 | 217660 | 217660 | 217660 | 217660 | 217660 |
| 2020-08-04 | 217808 | 217808 | 217808 | 217808 | 217808 | 217808 |
| 2020-08-05 | 217947 | 217947 | 217947 | 217947 | 217947 | 217947 |
| 2020-08-06 | 218081 | 218081 | 218081 | 218081 | 218081 | 218081 |
| 2020-08-07 | 218210 | 218210 | 218210 | 218210 | 218210 | 218210 |
| 2020-08-08 | 218337 | 218337 | 218337 | 218337 | 218337 | 218337 |
| 2020-08-09 | 218458 | 218458 | 218458 | 218458 | 218458 | 218458 |
| 2020-08-10 | 218586 | 218586 | 218586 | 218586 | 218586 | 218586 |
| 2020-08-11 | 218706 | 218706 | 218706 | 218706 | 218706 | 218706 |
| 2020-08-12 | 218822 | 218822 | 218822 | 218822 | 218822 | 218822 |
| 2020-08-13 | 218940 | 218940 | 218940 | 218940 | 218940 | 218940 |
| 2020-08-14 | 219048 | 219048 | 219048 | 219048 | 219048 | 219048 |
| 2020-08-15 | 219156 | 219156 | 219156 | 219156 | 219156 | 219156 |
| 2020-08-16 | 219265 | 219265 | 219265 | 219265 | 219265 | 219265 |
| 2020-08-17 | 219368 | 219368 | 219368 | 219368 | 219368 | 219368 |
| 2020-08-18 | 219475 | 219475 | 219475 | 219475 | 219475 | 219475 |
| 2020-08-19 | 219578 | 219578 | 219578 | 219578 | 219578 | 219578 |
| 2020-08-20 | 219685 | 219685 | 219685 | 219685 | 219685 | 219685 |
| 2020-08-21 | 219786 | 219786 | 219786 | 219786 | 219786 | 219786 |
| 2020-08-22 | 219882 | 219882 | 219882 | 219882 | 219882 | 219882 |
| 2020-08-23 | 219976 | 219976 | 219976 | 219976 | 219976 | 219976 |
| 2020-08-24 | 220079 | 220079 | 220079 | 220079 | 220079 | 220079 |
| 2020-08-25 | 220175 | 220175 | 220175 | 220175 | 220175 | 220175 |
| 2020-08-26 | 220274 | 220274 | 220274 | 220274 | 220274 | 220274 |
| 2020-08-27 | 220371 | 220371 | 220371 | 220371 | 220371 | 220371 |
| 2020-08-28 | 220472 | 220472 | 220472 | 220472 | 220472 | 220472 |
| 2020-08-29 | 220571 | 220571 | 220571 | 220571 | 220571 | 220571 |
| 2020-08-30 | 220662 | 220662 | 220662 | 220662 | 220662 | 220662 |
| 2020-08-31 | 220827 | 220827 | 220827 | 220827 | 220827 | 220827 |
| 2020-09-01 | 220984 | 220984 | 220984 | 220984 | 220984 | 220984 |
| 2020-09-02 | 221135 | 221135 | 221135 | 221135 | 221135 | 221135 |
| 2020-09-03 | 221304 | 221304 | 221304 | 221304 | 221304 | 221304 |
| 2020-09-04 | 221486 | 221486 | 221486 | 221486 | 221486 | 221486 |
| 2020-09-05 | 221678 | 221678 | 221678 | 221678 | 221678 | 221678 |
| 2020-09-06 | 221879 | 221879 | 221879 | 221879 | 221879 | 221879 |
| 2020-09-07 | 222095 | 222095 | 222095 | 222095 | 222095 | 222095 |
| 2020-09-08 | 222327 | 222327 | 222327 | 222327 | 222327 | 222327 |
| 2020-09-09 | 222576 | 222576 | 222576 | 222576 | 222576 | 222576 |
| 2020-09-10 | 222839 | 222839 | 222839 | 222839 | 222839 | 222839 |
| 2020-09-11 | 223110 | 223110 | 223110 | 223110 | 223110 | 223110 |
| 2020-09-12 | 223385 | 223385 | 223385 | 223385 | 223385 | 223385 |
| 2020-09-13 | 223697 | 223697 | 223697 | 223697 | 223697 | 223697 |
| 2020-09-14 | 224019 | 224019 | 224019 | 224019 | 224019 | 224019 |
| 2020-09-15 | 224346 | 224346 | 224346 | 224346 | 224346 | 224346 |
| 2020-09-16 | 224689 | 224689 | 224689 | 224689 | 224689 | 224689 |
| 2020-09-17 | 225068 | 225068 | 225068 | 225068 | 225068 | 225068 |
| 2020-09-18 | 225459 | 225459 | 225459 | 225459 | 225459 | 225459 |
| 2020-09-19 | 225875 | 225875 | 225875 | 225875 | 225875 | 225875 |
| 2020-09-20 | 226316 | 226316 | 226316 | 226316 | 226316 | 226316 |
| 2020-09-21 | 226777 | 226777 | 226777 | 226777 | 226777 | 226777 |
| 2020-09-22 | 227245 | 227245 | 227245 | 227245 | 227245 | 227245 |
| 2020-09-23 | 227758 | 227758 | 227758 | 227758 | 227758 | 227758 |
| 2020-09-24 | 228292 | 228292 | 228292 | 228292 | 228292 | 228292 |
| 2020-09-25 | 228831 | 228831 | 228831 | 228831 | 228831 | 228831 |
| 2020-09-26 | 229421 | 229421 | 229421 | 229421 | 229421 | 229421 |
| 2020-09-27 | 230029 | 230029 | 230029 | 230029 | 230029 | 230029 |
| 2020-09-28 | 230662 | 230662 | 230662 | 230662 | 230662 | 230662 |
| 2020-09-29 | 231338 | 231338 | 231338 | 231338 | 231338 | 231338 |
| 2020-09-30 | 231876 | 231876 | 231876 | 231876 | 231876 | 231876 |

f. NYC, adherence to NPIs is dynamic

Total number of confirmed cases

|  | No Vaccine | 25% | 50% | 60% | 75% | 100% Vaccinated |
| --- | --- | --- | --- | --- | --- | --- |
| 2020-03-15 | 0 | 0 | 0 | 0 | 0 | 0 |
| 2020-03-16 | 0 | 0 | 0 | 0 | 0 | 0 |
| 2020-03-17 | 0 | 0 | 0 | 0 | 0 | 0 |
| 2020-03-18 | 0 | 0 | 0 | 0 | 0 | 0 |
| 2020-03-19 | 16 | 16 | 16 | 16 | 16 | 16 |
| 2020-03-20 | 136 | 136 | 136 | 136 | 136 | 136 |
| 2020-03-21 | 256 | 256 | 256 | 256 | 256 | 256 |
| 2020-03-22 | 398 | 398 | 398 | 398 | 398 | 398 |
| 2020-03-23 | 635 | 635 | 635 | 635 | 635 | 635 |
| 2020-03-24 | 970 | 970 | 970 | 970 | 970 | 970 |
| 2020-03-25 | 1416 | 1416 | 1416 | 1416 | 1416 | 1416 |
| 2020-03-26 | 2088 | 2088 | 2088 | 2088 | 2088 | 2088 |
| 2020-03-27 | 3019 | 3019 | 3019 | 3019 | 3019 | 3019 |
| 2020-03-28 | 4209 | 4209 | 4209 | 4209 | 4209 | 4209 |
| 2020-03-29 | 5744 | 5744 | 5744 | 5744 | 5744 | 5744 |
| 2020-03-30 | 7623 | 7623 | 7623 | 7623 | 7623 | 7623 |
| 2020-03-31 | 9794 | 9794 | 9794 | 9794 | 9794 | 9794 |
| 2020-04-01 | 12226 | 12226 | 12226 | 12226 | 12226 | 12226 |
| 2020-04-02 | 14803 | 14803 | 14803 | 14803 | 14803 | 14803 |
| 2020-04-03 | 17237 | 17237 | 17237 | 17237 | 17237 | 17237 |
| 2020-04-04 | 19264 | 19264 | 19264 | 19264 | 19264 | 19264 |
| 2020-04-05 | 21702 | 21702 | 21702 | 21702 | 21702 | 21702 |
| 2020-04-06 | 24464 | 24464 | 24464 | 24464 | 24464 | 24464 |
| 2020-04-07 | 27356 | 27356 | 27356 | 27356 | 27356 | 27356 |
| 2020-04-08 | 30399 | 30399 | 30399 | 30399 | 30399 | 30399 |
| 2020-04-09 | 33612 | 33612 | 33612 | 33612 | 33612 | 33612 |
| 2020-04-10 | 36926 | 36926 | 36926 | 36926 | 36926 | 36926 |
| 2020-04-11 | 40383 | 40383 | 40383 | 40383 | 40383 | 40383 |
| 2020-04-12 | 43962 | 43962 | 43962 | 43962 | 43962 | 43962 |
| 2020-04-13 | 47704 | 47704 | 47704 | 47704 | 47704 | 47704 |
| 2020-04-14 | 51558 | 51558 | 51558 | 51558 | 51558 | 51558 |
| 2020-04-15 | 55724 | 55724 | 55724 | 55724 | 55724 | 55724 |
| 2020-04-16 | 60041 | 60041 | 60041 | 60041 | 60041 | 60041 |
| 2020-04-17 | 64551 | 64551 | 64551 | 64551 | 64551 | 64551 |
| 2020-04-18 | 69293 | 69293 | 69293 | 69293 | 69293 | 69293 |
| 2020-04-19 | 74174 | 74174 | 74174 | 74174 | 74174 | 74174 |
| 2020-04-20 | 79294 | 79294 | 79294 | 79294 | 79294 | 79294 |
| 2020-04-21 | 84677 | 84677 | 84677 | 84677 | 84677 | 84677 |
| 2020-04-22 | 90188 | 90188 | 90188 | 90188 | 90188 | 90188 |
| 2020-04-23 | 96006 | 96006 | 96006 | 96006 | 96006 | 96006 |
| 2020-04-24 | 102121 | 102121 | 102121 | 102121 | 102121 | 102121 |
| 2020-04-25 | 108476 | 108476 | 108476 | 108476 | 108476 | 108476 |
| 2020-04-26 | 115583 | 115583 | 115583 | 115583 | 115583 | 115583 |
| 2020-04-27 | 122960 | 122960 | 122960 | 122960 | 122960 | 122960 |
| 2020-04-28 | 126975 | 126975 | 126975 | 126975 | 126975 | 126975 |
| 2020-04-29 | 131186 | 131186 | 131186 | 131186 | 131186 | 131186 |
| 2020-04-30 | 135585 | 135585 | 135585 | 135585 | 135585 | 135585 |
| 2020-05-01 | 139848 | 139848 | 139848 | 139848 | 139848 | 139848 |
| 2020-05-02 | 143941 | 143941 | 143941 | 143941 | 143941 | 143941 |
| 2020-05-03 | 147885 | 147885 | 147885 | 147885 | 147885 | 147885 |
| 2020-05-04 | 151528 | 151528 | 151528 | 151528 | 151528 | 151528 |
| 2020-05-05 | 154909 | 154909 | 154909 | 154909 | 154909 | 154909 |
| 2020-05-06 | 158114 | 158114 | 158114 | 158114 | 158114 | 158114 |
| 2020-05-07 | 161007 | 161007 | 161007 | 161007 | 161007 | 161007 |
| 2020-05-08 | 163601 | 163601 | 163601 | 163601 | 163601 | 163601 |
| 2020-05-09 | 166148 | 166148 | 166148 | 166148 | 166148 | 166148 |
| 2020-05-10 | 168612 | 168612 | 168612 | 168612 | 168612 | 168612 |
| 2020-05-11 | 170889 | 170889 | 170889 | 170889 | 170889 | 170889 |
| 2020-05-12 | 173053 | 173053 | 173053 | 173053 | 173053 | 173053 |
| 2020-05-13 | 175078 | 175078 | 175078 | 175078 | 175078 | 175078 |
| 2020-05-14 | 176994 | 176994 | 176994 | 176994 | 176994 | 176994 |
| 2020-05-15 | 178782 | 178782 | 178782 | 178782 | 178782 | 178782 |
| 2020-05-16 | 180446 | 180446 | 180446 | 180446 | 180446 | 180446 |
| 2020-05-17 | 182009 | 182009 | 182009 | 182009 | 182009 | 182009 |
| 2020-05-18 | 183465 | 183465 | 183465 | 183465 | 183465 | 183465 |
| 2020-05-19 | 184859 | 184859 | 184859 | 184859 | 184859 | 184859 |
| 2020-05-20 | 186173 | 186173 | 186173 | 186173 | 186173 | 186173 |
| 2020-05-21 | 187546 | 187546 | 187546 | 187546 | 187546 | 187546 |
| 2020-05-22 | 188824 | 188824 | 188824 | 188824 | 188824 | 188824 |
| 2020-05-23 | 190036 | 190036 | 190036 | 190036 | 190036 | 190036 |
| 2020-05-24 | 191206 | 191206 | 191206 | 191206 | 191206 | 191206 |
| 2020-05-25 | 192290 | 192290 | 192290 | 192290 | 192290 | 192290 |
| 2020-05-26 | 193291 | 193291 | 193291 | 193291 | 193291 | 193291 |
| 2020-05-27 | 194217 | 194217 | 194217 | 194217 | 194217 | 194217 |
| 2020-05-28 | 195088 | 195088 | 195088 | 195088 | 195088 | 195088 |
| 2020-05-29 | 195887 | 195887 | 195887 | 195887 | 195887 | 195887 |
| 2020-05-30 | 196657 | 196657 | 196657 | 196657 | 196657 | 196657 |
| 2020-05-31 | 197411 | 197411 | 197411 | 197411 | 197411 | 197411 |
| 2020-06-01 | 198095 | 198095 | 198095 | 198095 | 198095 | 198095 |
| 2020-06-02 | 198759 | 198759 | 198759 | 198759 | 198759 | 198759 |
| 2020-06-03 | 199378 | 199378 | 199378 | 199378 | 199378 | 199378 |
| 2020-06-04 | 199977 | 199977 | 199977 | 199977 | 199977 | 199977 |
| 2020-06-05 | 200537 | 200537 | 200537 | 200537 | 200537 | 200537 |
| 2020-06-06 | 201048 | 201048 | 201048 | 201048 | 201048 | 201048 |
| 2020-06-07 | 201534 | 201534 | 201534 | 201534 | 201534 | 201534 |
| 2020-06-08 | 201998 | 201998 | 201998 | 201998 | 201998 | 201998 |
| 2020-06-09 | 202433 | 202433 | 202433 | 202433 | 202433 | 202433 |
| 2020-06-10 | 202856 | 202856 | 202856 | 202856 | 202856 | 202856 |
| 2020-06-11 | 203261 | 203261 | 203261 | 203261 | 203261 | 203261 |
| 2020-06-12 | 203643 | 203643 | 203643 | 203643 | 203643 | 203643 |
| 2020-06-13 | 204004 | 204004 | 204004 | 204004 | 204004 | 204004 |
| 2020-06-14 | 204348 | 204348 | 204348 | 204348 | 204348 | 204348 |
| 2020-06-15 | 204669 | 204669 | 204669 | 204669 | 204669 | 204669 |
| 2020-06-16 | 204976 | 204976 | 204976 | 204976 | 204976 | 204976 |
| 2020-06-17 | 205263 | 205263 | 205263 | 205263 | 205263 | 205263 |
| 2020-06-18 | 205540 | 205540 | 205540 | 205540 | 205540 | 205540 |
| 2020-06-19 | 205812 | 205812 | 205812 | 205812 | 205812 | 205812 |
| 2020-06-20 | 206067 | 206067 | 206067 | 206067 | 206067 | 206067 |
| 2020-06-21 | 206310 | 206310 | 206310 | 206310 | 206310 | 206310 |
| 2020-06-22 | 206544 | 206544 | 206544 | 206544 | 206544 | 206544 |
| 2020-06-23 | 206858 | 206858 | 206858 | 206858 | 206858 | 206858 |
| 2020-06-24 | 207155 | 207155 | 207155 | 207155 | 207155 | 207155 |
| 2020-06-25 | 207452 | 207452 | 207452 | 207452 | 207452 | 207452 |
| 2020-06-26 | 207741 | 207741 | 207741 | 207741 | 207741 | 207741 |
| 2020-06-27 | 208025 | 208025 | 208025 | 208025 | 208025 | 208025 |
| 2020-06-28 | 208306 | 208306 | 208306 | 208306 | 208306 | 208306 |
| 2020-06-29 | 208599 | 208599 | 208599 | 208599 | 208599 | 208599 |
| 2020-06-30 | 208884 | 208884 | 208884 | 208884 | 208884 | 208884 |
| 2020-07-01 | 209187 | 209187 | 209187 | 209187 | 209187 | 209187 |
| 2020-07-02 | 209502 | 209502 | 209502 | 209502 | 209502 | 209502 |
| 2020-07-03 | 209809 | 209809 | 209809 | 209809 | 209809 | 209809 |
| 2020-07-04 | 210126 | 210126 | 210126 | 210126 | 210126 | 210126 |
| 2020-07-05 | 210438 | 210438 | 210438 | 210438 | 210438 | 210438 |
| 2020-07-06 | 210748 | 210748 | 210748 | 210748 | 210748 | 210748 |
| 2020-07-07 | 211054 | 211054 | 211054 | 211054 | 211054 | 211054 |
| 2020-07-08 | 211376 | 211376 | 211376 | 211376 | 211376 | 211376 |
| 2020-07-09 | 211704 | 211704 | 211704 | 211704 | 211704 | 211704 |
| 2020-07-10 | 212042 | 212042 | 212042 | 212042 | 212042 | 212042 |
| 2020-07-11 | 212380 | 212380 | 212380 | 212380 | 212380 | 212380 |
| 2020-07-12 | 212716 | 212716 | 212716 | 212716 | 212716 | 212716 |
| 2020-07-13 | 213052 | 213052 | 213052 | 213052 | 213052 | 213052 |
| 2020-07-14 | 213393 | 213393 | 213393 | 213393 | 213393 | 213393 |
| 2020-07-15 | 213733 | 213733 | 213733 | 213733 | 213733 | 213733 |
| 2020-07-16 | 214082 | 214082 | 214082 | 214082 | 214082 | 214082 |
| 2020-07-17 | 214314 | 214314 | 214314 | 214314 | 214314 | 214314 |
| 2020-07-18 | 214561 | 214561 | 214561 | 214561 | 214561 | 214561 |
| 2020-07-19 | 214811 | 214811 | 214811 | 214811 | 214811 | 214811 |
| 2020-07-20 | 215052 | 215052 | 215052 | 215052 | 215052 | 215052 |
| 2020-07-21 | 215296 | 215296 | 215296 | 215296 | 215296 | 215296 |
| 2020-07-22 | 215526 | 215526 | 215526 | 215526 | 215526 | 215526 |
| 2020-07-23 | 215742 | 215742 | 215742 | 215742 | 215742 | 215742 |
| 2020-07-24 | 215945 | 215945 | 215945 | 215945 | 215945 | 215945 |
| 2020-07-25 | 216138 | 216138 | 216138 | 216138 | 216138 | 216138 |
| 2020-07-26 | 216333 | 216333 | 216333 | 216333 | 216333 | 216333 |
| 2020-07-27 | 216513 | 216513 | 216513 | 216513 | 216513 | 216513 |
| 2020-07-28 | 216692 | 216692 | 216692 | 216692 | 216692 | 216692 |
| 2020-07-29 | 216872 | 216872 | 216872 | 216872 | 216872 | 216872 |
| 2020-07-30 | 217036 | 217036 | 217036 | 217036 | 217036 | 217036 |
| 2020-07-31 | 217196 | 217196 | 217196 | 217196 | 217196 | 217196 |
| 2020-08-01 | 217359 | 217359 | 217359 | 217359 | 217359 | 217359 |
| 2020-08-02 | 217513 | 217513 | 217513 | 217513 | 217513 | 217513 |
| 2020-08-03 | 217660 | 217660 | 217660 | 217660 | 217660 | 217660 |
| 2020-08-04 | 217808 | 217808 | 217808 | 217808 | 217808 | 217808 |
| 2020-08-05 | 217947 | 217947 | 217947 | 217947 | 217947 | 217947 |
| 2020-08-06 | 218081 | 218081 | 218081 | 218081 | 218081 | 218081 |
| 2020-08-07 | 218210 | 218210 | 218210 | 218210 | 218210 | 218210 |
| 2020-08-08 | 218337 | 218337 | 218337 | 218337 | 218337 | 218337 |
| 2020-08-09 | 218458 | 218458 | 218458 | 218458 | 218458 | 218458 |
| 2020-08-10 | 218586 | 218586 | 218586 | 218586 | 218586 | 218586 |
| 2020-08-11 | 218706 | 218706 | 218706 | 218706 | 218706 | 218706 |
| 2020-08-12 | 218822 | 218822 | 218822 | 218822 | 218822 | 218822 |
| 2020-08-13 | 218940 | 218940 | 218940 | 218940 | 218940 | 218940 |
| 2020-08-14 | 219048 | 219048 | 219048 | 219048 | 219048 | 219048 |
| 2020-08-15 | 219156 | 219156 | 219156 | 219156 | 219156 | 219156 |
| 2020-08-16 | 219265 | 219265 | 219265 | 219265 | 219265 | 219265 |
| 2020-08-17 | 219368 | 219368 | 219368 | 219368 | 219368 | 219368 |
| 2020-08-18 | 219475 | 219475 | 219475 | 219475 | 219475 | 219475 |
| 2020-08-19 | 219578 | 219578 | 219578 | 219578 | 219578 | 219578 |
| 2020-08-20 | 219685 | 219685 | 219685 | 219685 | 219685 | 219685 |
| 2020-08-21 | 219786 | 219786 | 219786 | 219786 | 219786 | 219786 |
| 2020-08-22 | 219882 | 219882 | 219882 | 219882 | 219882 | 219882 |
| 2020-08-23 | 219976 | 219976 | 219976 | 219976 | 219976 | 219976 |
| 2020-08-24 | 220079 | 220079 | 220079 | 220079 | 220079 | 220079 |
| 2020-08-25 | 220175 | 220175 | 220175 | 220175 | 220175 | 220175 |
| 2020-08-26 | 220274 | 220274 | 220274 | 220274 | 220274 | 220274 |
| 2020-08-27 | 220371 | 220371 | 220371 | 220371 | 220371 | 220371 |
| 2020-08-28 | 220472 | 220472 | 220472 | 220472 | 220472 | 220472 |
| 2020-08-29 | 220571 | 220571 | 220571 | 220571 | 220571 | 220571 |
| 2020-08-30 | 220662 | 220662 | 220662 | 220662 | 220662 | 220662 |
| 2020-08-31 | 220827 | 220827 | 220827 | 220827 | 220827 | 220827 |
| 2020-09-01 | 220984 | 220984 | 220984 | 220984 | 220984 | 220984 |
| 2020-09-02 | 221135 | 221135 | 221135 | 221135 | 221135 | 221135 |
| 2020-09-03 | 221304 | 221304 | 221304 | 221304 | 221304 | 221304 |
| 2020-09-04 | 221486 | 221486 | 221486 | 221486 | 221486 | 221486 |
| 2020-09-05 | 221678 | 221678 | 221678 | 221678 | 221678 | 221678 |
| 2020-09-06 | 221879 | 221879 | 221879 | 221879 | 221879 | 221879 |
| 2020-09-07 | 222095 | 222095 | 222095 | 222095 | 222095 | 222095 |
| 2020-09-08 | 222327 | 222327 | 222327 | 222327 | 222327 | 222327 |
| 2020-09-09 | 222576 | 222576 | 222576 | 222576 | 222576 | 222576 |
| 2020-09-10 | 222839 | 222839 | 222839 | 222839 | 222839 | 222839 |
| 2020-09-11 | 223110 | 223110 | 223110 | 223110 | 223110 | 223110 |
| 2020-09-12 | 223385 | 223385 | 223385 | 223385 | 223385 | 223385 |
| 2020-09-13 | 223697 | 223697 | 223697 | 223697 | 223697 | 223697 |
| 2020-09-14 | 224019 | 224019 | 224019 | 224019 | 224019 | 224019 |
| 2020-09-15 | 224346 | 224346 | 224346 | 224346 | 224346 | 224346 |
| 2020-09-16 | 224689 | 224689 | 224689 | 224689 | 224689 | 224689 |
| 2020-09-17 | 225068 | 225068 | 225068 | 225068 | 225068 | 225068 |
| 2020-09-18 | 225459 | 225459 | 225459 | 225459 | 225459 | 225459 |
| 2020-09-19 | 225875 | 225875 | 225875 | 225875 | 225875 | 225875 |
| 2020-09-20 | 226316 | 226316 | 226316 | 226316 | 226316 | 226316 |
| 2020-09-21 | 226777 | 226777 | 226777 | 226777 | 226777 | 226777 |
| 2020-09-22 | 227245 | 227245 | 227245 | 227245 | 227245 | 227245 |
| 2020-09-23 | 227758 | 227758 | 227758 | 227758 | 227758 | 227758 |
| 2020-09-24 | 228292 | 228292 | 228292 | 228292 | 228292 | 228292 |
| 2020-09-25 | 228831 | 228831 | 228831 | 228831 | 228831 | 228831 |
| 2020-09-26 | 229421 | 229421 | 229421 | 229421 | 229421 | 229421 |
| 2020-09-27 | 230029 | 230029 | 230029 | 230029 | 230029 | 230029 |
| 2020-09-28 | 230662 | 230662 | 230662 | 230662 | 230662 | 230662 |
| 2020-09-29 | 231338 | 231338 | 231338 | 231338 | 231338 | 231338 |
| 2020-09-30 | 231876 | 231876 | 231876 | 231876 | 231876 | 231876 |

Supplement Figure 3. Impact of vaccination and adherence to nonpharmaceutical interventions on the number of cases over time when there is no drop in adherence after vaccination. In these experiments, we assumed that vaccine effectiveness is 90%, vaccine coverage is 50%, and daily vaccination capacity is 0.1% for the vaccination scenario.

a. Dane, vaccination

Total number of confirmed cases

|  | 75% Adherence | 70% | 65% | 60% | Dynamic Adherence |
| --- | --- | --- | --- | --- | --- |
| 2020-03-15 | 0 | 0 | 0 | 0 | 0 |
| 2020-03-16 | 0 | 0 | 0 | 0 | 0 |
| 2020-03-17 | 0 | 0 | 0 | 0 | 0 |
| 2020-03-18 | 0 | 0 | 0 | 0 | 0 |
| 2020-03-19 | 1 | 1 | 1 | 1 | 1 |
| 2020-03-20 | 3 | 3 | 3 | 3 | 3 |
| 2020-03-21 | 5 | 5 | 5 | 5 | 5 |
| 2020-03-22 | 9 | 9 | 9 | 9 | 9 |
| 2020-03-23 | 12 | 12 | 12 | 12 | 12 |
| 2020-03-24 | 17 | 17 | 17 | 17 | 17 |
| 2020-03-25 | 23 | 23 | 23 | 23 | 23 |
| 2020-03-26 | 30 | 30 | 30 | 30 | 30 |
| 2020-03-27 | 39 | 39 | 39 | 39 | 39 |
| 2020-03-28 | 51 | 51 | 51 | 51 | 51 |
| 2020-03-29 | 64 | 64 | 64 | 64 | 64 |
| 2020-03-30 | 79 | 79 | 79 | 79 | 79 |
| 2020-03-31 | 96 | 96 | 96 | 96 | 96 |
| 2020-04-01 | 116 | 116 | 116 | 116 | 116 |
| 2020-04-02 | 135 | 135 | 135 | 135 | 135 |
| 2020-04-03 | 155 | 155 | 155 | 155 | 155 |
| 2020-04-04 | 174 | 174 | 174 | 174 | 174 |
| 2020-04-05 | 193 | 193 | 193 | 193 | 193 |
| 2020-04-06 | 210 | 210 | 210 | 210 | 210 |
| 2020-04-07 | 225 | 225 | 225 | 225 | 225 |
| 2020-04-08 | 237 | 237 | 237 | 237 | 237 |
| 2020-04-09 | 244 | 244 | 244 | 244 | 244 |
| 2020-04-10 | 258 | 258 | 258 | 258 | 258 |
| 2020-04-11 | 272 | 272 | 272 | 272 | 272 |
| 2020-04-12 | 284 | 284 | 284 | 284 | 284 |
| 2020-04-13 | 295 | 295 | 295 | 295 | 295 |
| 2020-04-14 | 305 | 305 | 305 | 305 | 305 |
| 2020-04-15 | 314 | 314 | 314 | 314 | 314 |
| 2020-04-16 | 323 | 323 | 323 | 323 | 323 |
| 2020-04-17 | 332 | 332 | 332 | 332 | 332 |
| 2020-04-18 | 340 | 340 | 340 | 340 | 340 |
| 2020-04-19 | 348 | 348 | 348 | 348 | 348 |
| 2020-04-20 | 356 | 356 | 356 | 356 | 356 |
| 2020-04-21 | 363 | 363 | 363 | 363 | 363 |
| 2020-04-22 | 370 | 370 | 370 | 370 | 370 |
| 2020-04-23 | 377 | 377 | 377 | 377 | 377 |
| 2020-04-24 | 383 | 383 | 383 | 383 | 383 |
| 2020-04-25 | 390 | 390 | 390 | 390 | 390 |
| 2020-04-26 | 396 | 396 | 396 | 396 | 396 |
| 2020-04-27 | 402 | 402 | 402 | 402 | 402 |
| 2020-04-28 | 408 | 408 | 408 | 408 | 408 |
| 2020-04-29 | 414 | 414 | 414 | 414 | 414 |
| 2020-04-30 | 420 | 420 | 420 | 420 | 420 |
| 2020-05-01 | 425 | 425 | 425 | 425 | 425 |
| 2020-05-02 | 430 | 430 | 430 | 430 | 430 |
| 2020-05-03 | 435 | 435 | 435 | 435 | 435 |
| 2020-05-04 | 440 | 440 | 440 | 440 | 440 |
| 2020-05-05 | 445 | 445 | 445 | 445 | 445 |
| 2020-05-06 | 449 | 449 | 449 | 449 | 449 |
| 2020-05-07 | 454 | 454 | 454 | 454 | 454 |
| 2020-05-08 | 459 | 459 | 459 | 459 | 459 |
| 2020-05-09 | 463 | 463 | 463 | 463 | 463 |
| 2020-05-10 | 467 | 467 | 467 | 467 | 467 |
| 2020-05-11 | 472 | 472 | 472 | 472 | 472 |
| 2020-05-12 | 476 | 476 | 476 | 476 | 476 |
| 2020-05-13 | 479 | 479 | 479 | 479 | 479 |
| 2020-05-14 | 483 | 483 | 483 | 483 | 483 |
| 2020-05-15 | 487 | 487 | 487 | 487 | 487 |
| 2020-05-16 | 492 | 492 | 492 | 492 | 492 |
| 2020-05-17 | 497 | 497 | 497 | 497 | 497 |
| 2020-05-18 | 502 | 502 | 502 | 502 | 502 |
| 2020-05-19 | 507 | 507 | 507 | 507 | 507 |
| 2020-05-20 | 512 | 512 | 512 | 512 | 512 |
| 2020-05-21 | 519 | 519 | 519 | 519 | 519 |
| 2020-05-22 | 525 | 525 | 525 | 525 | 525 |
| 2020-05-23 | 531 | 531 | 531 | 531 | 531 |
| 2020-05-24 | 537 | 537 | 537 | 537 | 537 |
| 2020-05-25 | 544 | 544 | 544 | 544 | 544 |
| 2020-05-26 | 551 | 551 | 551 | 551 | 551 |
| 2020-05-27 | 558 | 558 | 558 | 558 | 558 |
| 2020-05-28 | 565 | 565 | 565 | 565 | 565 |
| 2020-05-29 | 574 | 574 | 574 | 574 | 574 |
| 2020-05-30 | 583 | 583 | 583 | 583 | 583 |
| 2020-05-31 | 593 | 593 | 593 | 593 | 593 |
| 2020-06-01 | 603 | 603 | 603 | 603 | 603 |
| 2020-06-02 | 614 | 614 | 614 | 614 | 614 |
| 2020-06-03 | 625 | 625 | 625 | 625 | 625 |
| 2020-06-04 | 636 | 636 | 636 | 636 | 636 |
| 2020-06-05 | 649 | 649 | 649 | 649 | 649 |
| 2020-06-06 | 662 | 662 | 662 | 662 | 662 |
| 2020-06-07 | 676 | 676 | 676 | 676 | 676 |
| 2020-06-08 | 691 | 691 | 691 | 691 | 691 |
| 2020-06-09 | 707 | 707 | 707 | 707 | 707 |
| 2020-06-10 | 723 | 723 | 723 | 723 | 723 |
| 2020-06-11 | 740 | 740 | 740 | 740 | 740 |
| 2020-06-12 | 759 | 759 | 759 | 759 | 759 |
| 2020-06-13 | 778 | 778 | 778 | 778 | 778 |
| 2020-06-14 | 797 | 797 | 797 | 797 | 797 |
| 2020-06-15 | 818 | 818 | 818 | 818 | 818 |
| 2020-06-16 | 840 | 840 | 840 | 840 | 840 |
| 2020-06-17 | 863 | 863 | 863 | 863 | 863 |
| 2020-06-18 | 887 | 887 | 887 | 887 | 887 |
| 2020-06-19 | 913 | 913 | 913 | 913 | 913 |
| 2020-06-20 | 940 | 940 | 940 | 940 | 940 |
| 2020-06-21 | 968 | 968 | 968 | 968 | 968 |
| 2020-06-22 | 997 | 997 | 997 | 997 | 997 |
| 2020-06-23 | 1028 | 1028 | 1028 | 1028 | 1028 |
| 2020-06-24 | 1061 | 1061 | 1061 | 1061 | 1061 |
| 2020-06-25 | 1096 | 1096 | 1096 | 1096 | 1096 |
| 2020-06-26 | 1132 | 1132 | 1132 | 1132 | 1132 |
| 2020-06-27 | 1169 | 1169 | 1169 | 1169 | 1169 |
| 2020-06-28 | 1207 | 1207 | 1207 | 1207 | 1207 |
| 2020-06-29 | 1249 | 1249 | 1249 | 1249 | 1249 |
| 2020-06-30 | 1292 | 1292 | 1292 | 1292 | 1292 |
| 2020-07-01 | 1338 | 1338 | 1338 | 1338 | 1338 |
| 2020-07-02 | 1385 | 1385 | 1385 | 1385 | 1385 |
| 2020-07-03 | 1434 | 1434 | 1434 | 1434 | 1434 |
| 2020-07-04 | 1486 | 1486 | 1486 | 1486 | 1486 |
| 2020-07-05 | 1541 | 1541 | 1541 | 1541 | 1541 |
| 2020-07-06 | 1597 | 1597 | 1597 | 1597 | 1597 |
| 2020-07-07 | 1657 | 1657 | 1657 | 1657 | 1657 |
| 2020-07-08 | 1721 | 1721 | 1721 | 1721 | 1721 |
| 2020-07-09 | 1787 | 1787 | 1787 | 1787 | 1787 |
| 2020-07-10 | 1855 | 1855 | 1855 | 1855 | 1855 |
| 2020-07-11 | 1927 | 1927 | 1927 | 1927 | 1927 |
| 2020-07-12 | 2003 | 2003 | 2003 | 2003 | 2003 |
| 2020-07-13 | 2082 | 2082 | 2082 | 2082 | 2082 |
| 2020-07-14 | 2165 | 2165 | 2165 | 2165 | 2165 |
| 2020-07-15 | 2251 | 2251 | 2251 | 2251 | 2251 |
| 2020-07-16 | 2320 | 2320 | 2320 | 2320 | 2320 |
| 2020-07-17 | 2392 | 2392 | 2392 | 2392 | 2392 |
| 2020-07-18 | 2467 | 2467 | 2467 | 2467 | 2467 |
| 2020-07-19 | 2541 | 2541 | 2541 | 2541 | 2541 |
| 2020-07-20 | 2616 | 2616 | 2616 | 2616 | 2616 |
| 2020-07-21 | 2692 | 2692 | 2692 | 2692 | 2692 |
| 2020-07-22 | 2768 | 2768 | 2768 | 2768 | 2768 |
| 2020-07-23 | 2846 | 2846 | 2846 | 2846 | 2846 |
| 2020-07-24 | 2922 | 2922 | 2922 | 2922 | 2922 |
| 2020-07-25 | 2999 | 2999 | 2999 | 2999 | 2999 |
| 2020-07-26 | 3073 | 3073 | 3073 | 3073 | 3073 |
| 2020-07-27 | 3147 | 3147 | 3147 | 3147 | 3147 |
| 2020-07-28 | 3222 | 3222 | 3222 | 3222 | 3222 |
| 2020-07-29 | 3299 | 3299 | 3299 | 3299 | 3299 |
| 2020-07-30 | 3376 | 3376 | 3376 | 3376 | 3376 |
| 2020-07-31 | 3453 | 3453 | 3453 | 3453 | 3453 |
| 2020-08-01 | 3528 | 3528 | 3528 | 3528 | 3528 |
| 2020-08-02 | 3605 | 3605 | 3605 | 3605 | 3605 |
| 2020-08-03 | 3682 | 3682 | 3682 | 3682 | 3682 |
| 2020-08-04 | 3757 | 3757 | 3757 | 3757 | 3757 |
| 2020-08-05 | 3832 | 3832 | 3832 | 3832 | 3832 |
| 2020-08-06 | 3908 | 3908 | 3908 | 3908 | 3908 |
| 2020-08-07 | 3983 | 3983 | 3983 | 3983 | 3983 |
| 2020-08-08 | 4060 | 4060 | 4060 | 4060 | 4060 |
| 2020-08-09 | 4137 | 4137 | 4137 | 4137 | 4137 |
| 2020-08-10 | 4213 | 4213 | 4213 | 4213 | 4213 |
| 2020-08-11 | 4290 | 4290 | 4290 | 4290 | 4290 |
| 2020-08-12 | 4364 | 4364 | 4364 | 4364 | 4364 |
| 2020-08-13 | 4439 | 4439 | 4439 | 4439 | 4439 |
| 2020-08-14 | 4516 | 4516 | 4516 | 4516 | 4516 |
| 2020-08-15 | 4592 | 4592 | 4592 | 4592 | 4592 |
| 2020-08-16 | 4669 | 4669 | 4669 | 4669 | 4669 |
| 2020-08-17 | 4747 | 4747 | 4747 | 4747 | 4747 |
| 2020-08-18 | 4823 | 4823 | 4823 | 4823 | 4823 |
| 2020-08-19 | 4900 | 4900 | 4900 | 4900 | 4900 |
| 2020-08-20 | 4977 | 4977 | 4977 | 4977 | 4977 |
| 2020-08-21 | 5055 | 5055 | 5055 | 5055 | 5055 |
| 2020-08-22 | 5132 | 5132 | 5132 | 5132 | 5132 |
| 2020-08-23 | 5210 | 5210 | 5210 | 5210 | 5210 |
| 2020-08-24 | 5288 | 5288 | 5288 | 5288 | 5288 |
| 2020-08-25 | 5366 | 5366 | 5366 | 5366 | 5366 |
| 2020-08-26 | 5443 | 5443 | 5443 | 5443 | 5443 |
| 2020-08-27 | 5520 | 5520 | 5520 | 5520 | 5520 |
| 2020-08-28 | 5597 | 5597 | 5597 | 5597 | 5597 |
| 2020-08-29 | 5675 | 5675 | 5675 | 5675 | 5675 |
| 2020-08-30 | 5751 | 5751 | 5751 | 5751 | 5751 |
| 2020-08-31 | 5829 | 5829 | 5829 | 5829 | 5829 |
| 2020-09-01 | 5906 | 5906 | 5906 | 5906 | 5906 |
| 2020-09-02 | 5983 | 5983 | 5983 | 5983 | 5983 |
| 2020-09-03 | 6061 | 6061 | 6061 | 6061 | 6061 |
| 2020-09-04 | 6137 | 6137 | 6137 | 6137 | 6137 |
| 2020-09-05 | 6213 | 6213 | 6213 | 6213 | 6213 |
| 2020-09-06 | 6290 | 6290 | 6290 | 6290 | 6290 |
| 2020-09-07 | 6368 | 6368 | 6368 | 6368 | 6368 |
| 2020-09-08 | 6445 | 6445 | 6445 | 6445 | 6445 |
| 2020-09-09 | 6523 | 6523 | 6523 | 6523 | 6523 |
| 2020-09-10 | 6601 | 6601 | 6601 | 6601 | 6601 |
| 2020-09-11 | 6676 | 6676 | 6676 | 6676 | 6676 |
| 2020-09-12 | 6754 | 6754 | 6754 | 6754 | 6754 |
| 2020-09-13 | 6831 | 6831 | 6831 | 6831 | 6831 |
| 2020-09-14 | 6908 | 6908 | 6908 | 6908 | 6908 |
| 2020-09-15 | 6985 | 6985 | 6985 | 6985 | 6985 |
| 2020-09-16 | 7075 | 7075 | 7075 | 7075 | 7075 |
| 2020-09-17 | 7164 | 7164 | 7164 | 7164 | 7164 |
| 2020-09-18 | 7254 | 7254 | 7254 | 7254 | 7254 |
| 2020-09-19 | 7346 | 7346 | 7346 | 7346 | 7346 |
| 2020-09-20 | 7439 | 7439 | 7439 | 7439 | 7439 |
| 2020-09-21 | 7533 | 7533 | 7533 | 7533 | 7533 |
| 2020-09-22 | 7631 | 7631 | 7631 | 7631 | 7631 |
| 2020-09-23 | 7731 | 7731 | 7731 | 7731 | 7731 |
| 2020-09-24 | 7833 | 7833 | 7833 | 7833 | 7833 |
| 2020-09-25 | 7940 | 7940 | 7940 | 7940 | 7940 |
| 2020-09-26 | 8048 | 8048 | 8048 | 8048 | 8048 |
| 2020-09-27 | 8157 | 8157 | 8157 | 8157 | 8157 |
| 2020-09-28 | 8270 | 8270 | 8270 | 8270 | 8270 |
| 2020-09-29 | 8386 | 8386 | 8386 | 8386 | 8386 |
| 2020-09-30 | 8504 | 8504 | 8504 | 8504 | 8504 |

b. Dane, No vaccination

Total number of confirmed cases

|  | 75% Adherence | 70% | 65% | 60% | Dynamic Adherence |
| --- | --- | --- | --- | --- | --- |
| 2020-03-15 | 0 | 0 | 0 | 0 | 0 |
| 2020-03-16 | 0 | 0 | 0 | 0 | 0 |
| 2020-03-17 | 0 | 0 | 0 | 0 | 0 |
| 2020-03-18 | 0 | 0 | 0 | 0 | 0 |
| 2020-03-19 | 1 | 1 | 1 | 1 | 1 |
| 2020-03-20 | 3 | 3 | 3 | 3 | 3 |
| 2020-03-21 | 5 | 5 | 5 | 5 | 5 |
| 2020-03-22 | 9 | 9 | 9 | 9 | 9 |
| 2020-03-23 | 12 | 12 | 12 | 12 | 12 |
| 2020-03-24 | 17 | 17 | 17 | 17 | 17 |
| 2020-03-25 | 23 | 23 | 23 | 23 | 23 |
| 2020-03-26 | 30 | 30 | 30 | 30 | 30 |
| 2020-03-27 | 39 | 39 | 39 | 39 | 39 |
| 2020-03-28 | 51 | 51 | 51 | 51 | 51 |
| 2020-03-29 | 64 | 64 | 64 | 64 | 64 |
| 2020-03-30 | 79 | 79 | 79 | 79 | 79 |
| 2020-03-31 | 96 | 96 | 96 | 96 | 96 |
| 2020-04-01 | 116 | 116 | 116 | 116 | 116 |
| 2020-04-02 | 135 | 135 | 135 | 135 | 135 |
| 2020-04-03 | 155 | 155 | 155 | 155 | 155 |
| 2020-04-04 | 174 | 174 | 174 | 174 | 174 |
| 2020-04-05 | 193 | 193 | 193 | 193 | 193 |
| 2020-04-06 | 210 | 210 | 210 | 210 | 210 |
| 2020-04-07 | 225 | 225 | 225 | 225 | 225 |
| 2020-04-08 | 237 | 237 | 237 | 237 | 237 |
| 2020-04-09 | 244 | 244 | 244 | 244 | 244 |
| 2020-04-10 | 258 | 258 | 258 | 258 | 258 |
| 2020-04-11 | 272 | 272 | 272 | 272 | 272 |
| 2020-04-12 | 284 | 284 | 284 | 284 | 284 |
| 2020-04-13 | 295 | 295 | 295 | 295 | 295 |
| 2020-04-14 | 305 | 305 | 305 | 305 | 305 |
| 2020-04-15 | 314 | 314 | 314 | 314 | 314 |
| 2020-04-16 | 323 | 323 | 323 | 323 | 323 |
| 2020-04-17 | 332 | 332 | 332 | 332 | 332 |
| 2020-04-18 | 340 | 340 | 340 | 340 | 340 |
| 2020-04-19 | 348 | 348 | 348 | 348 | 348 |
| 2020-04-20 | 356 | 356 | 356 | 356 | 356 |
| 2020-04-21 | 363 | 363 | 363 | 363 | 363 |
| 2020-04-22 | 370 | 370 | 370 | 370 | 370 |
| 2020-04-23 | 377 | 377 | 377 | 377 | 377 |
| 2020-04-24 | 383 | 383 | 383 | 383 | 383 |
| 2020-04-25 | 390 | 390 | 390 | 390 | 390 |
| 2020-04-26 | 396 | 396 | 396 | 396 | 396 |
| 2020-04-27 | 402 | 402 | 402 | 402 | 402 |
| 2020-04-28 | 408 | 408 | 408 | 408 | 408 |
| 2020-04-29 | 414 | 414 | 414 | 414 | 414 |
| 2020-04-30 | 420 | 420 | 420 | 420 | 420 |
| 2020-05-01 | 425 | 425 | 425 | 425 | 425 |
| 2020-05-02 | 430 | 430 | 430 | 430 | 430 |
| 2020-05-03 | 435 | 435 | 435 | 435 | 435 |
| 2020-05-04 | 440 | 440 | 440 | 440 | 440 |
| 2020-05-05 | 445 | 445 | 445 | 445 | 445 |
| 2020-05-06 | 449 | 449 | 449 | 449 | 449 |
| 2020-05-07 | 454 | 454 | 454 | 454 | 454 |
| 2020-05-08 | 459 | 459 | 459 | 459 | 459 |
| 2020-05-09 | 463 | 463 | 463 | 463 | 463 |
| 2020-05-10 | 467 | 467 | 467 | 467 | 467 |
| 2020-05-11 | 472 | 472 | 472 | 472 | 472 |
| 2020-05-12 | 476 | 476 | 476 | 476 | 476 |
| 2020-05-13 | 479 | 479 | 479 | 479 | 479 |
| 2020-05-14 | 483 | 483 | 483 | 483 | 483 |
| 2020-05-15 | 487 | 487 | 487 | 487 | 487 |
| 2020-05-16 | 492 | 492 | 492 | 492 | 492 |
| 2020-05-17 | 497 | 497 | 497 | 497 | 497 |
| 2020-05-18 | 502 | 502 | 502 | 502 | 502 |
| 2020-05-19 | 507 | 507 | 507 | 507 | 507 |
| 2020-05-20 | 512 | 512 | 512 | 512 | 512 |
| 2020-05-21 | 519 | 519 | 519 | 519 | 519 |
| 2020-05-22 | 525 | 525 | 525 | 525 | 525 |
| 2020-05-23 | 531 | 531 | 531 | 531 | 531 |
| 2020-05-24 | 537 | 537 | 537 | 537 | 537 |
| 2020-05-25 | 544 | 544 | 544 | 544 | 544 |
| 2020-05-26 | 551 | 551 | 551 | 551 | 551 |
| 2020-05-27 | 558 | 558 | 558 | 558 | 558 |
| 2020-05-28 | 565 | 565 | 565 | 565 | 565 |
| 2020-05-29 | 574 | 574 | 574 | 574 | 574 |
| 2020-05-30 | 583 | 583 | 583 | 583 | 583 |
| 2020-05-31 | 593 | 593 | 593 | 593 | 593 |
| 2020-06-01 | 603 | 603 | 603 | 603 | 603 |
| 2020-06-02 | 614 | 614 | 614 | 614 | 614 |
| 2020-06-03 | 625 | 625 | 625 | 625 | 625 |
| 2020-06-04 | 636 | 636 | 636 | 636 | 636 |
| 2020-06-05 | 649 | 649 | 649 | 649 | 649 |
| 2020-06-06 | 662 | 662 | 662 | 662 | 662 |
| 2020-06-07 | 676 | 676 | 676 | 676 | 676 |
| 2020-06-08 | 691 | 691 | 691 | 691 | 691 |
| 2020-06-09 | 707 | 707 | 707 | 707 | 707 |
| 2020-06-10 | 723 | 723 | 723 | 723 | 723 |
| 2020-06-11 | 740 | 740 | 740 | 740 | 740 |
| 2020-06-12 | 759 | 759 | 759 | 759 | 759 |
| 2020-06-13 | 778 | 778 | 778 | 778 | 778 |
| 2020-06-14 | 797 | 797 | 797 | 797 | 797 |
| 2020-06-15 | 818 | 818 | 818 | 818 | 818 |
| 2020-06-16 | 840 | 840 | 840 | 840 | 840 |
| 2020-06-17 | 863 | 863 | 863 | 863 | 863 |
| 2020-06-18 | 887 | 887 | 887 | 887 | 887 |
| 2020-06-19 | 913 | 913 | 913 | 913 | 913 |
| 2020-06-20 | 940 | 940 | 940 | 940 | 940 |
| 2020-06-21 | 968 | 968 | 968 | 968 | 968 |
| 2020-06-22 | 997 | 997 | 997 | 997 | 997 |
| 2020-06-23 | 1028 | 1028 | 1028 | 1028 | 1028 |
| 2020-06-24 | 1061 | 1061 | 1061 | 1061 | 1061 |
| 2020-06-25 | 1096 | 1096 | 1096 | 1096 | 1096 |
| 2020-06-26 | 1132 | 1132 | 1132 | 1132 | 1132 |
| 2020-06-27 | 1169 | 1169 | 1169 | 1169 | 1169 |
| 2020-06-28 | 1207 | 1207 | 1207 | 1207 | 1207 |
| 2020-06-29 | 1249 | 1249 | 1249 | 1249 | 1249 |
| 2020-06-30 | 1292 | 1292 | 1292 | 1292 | 1292 |
| 2020-07-01 | 1338 | 1338 | 1338 | 1338 | 1338 |
| 2020-07-02 | 1385 | 1385 | 1385 | 1385 | 1385 |
| 2020-07-03 | 1434 | 1434 | 1434 | 1434 | 1434 |
| 2020-07-04 | 1486 | 1486 | 1486 | 1486 | 1486 |
| 2020-07-05 | 1541 | 1541 | 1541 | 1541 | 1541 |
| 2020-07-06 | 1597 | 1597 | 1597 | 1597 | 1597 |
| 2020-07-07 | 1657 | 1657 | 1657 | 1657 | 1657 |
| 2020-07-08 | 1721 | 1721 | 1721 | 1721 | 1721 |
| 2020-07-09 | 1787 | 1787 | 1787 | 1787 | 1787 |
| 2020-07-10 | 1855 | 1855 | 1855 | 1855 | 1855 |
| 2020-07-11 | 1927 | 1927 | 1927 | 1927 | 1927 |
| 2020-07-12 | 2003 | 2003 | 2003 | 2003 | 2003 |
| 2020-07-13 | 2082 | 2082 | 2082 | 2082 | 2082 |
| 2020-07-14 | 2165 | 2165 | 2165 | 2165 | 2165 |
| 2020-07-15 | 2251 | 2251 | 2251 | 2251 | 2251 |
| 2020-07-16 | 2320 | 2320 | 2320 | 2320 | 2320 |
| 2020-07-17 | 2392 | 2392 | 2392 | 2392 | 2392 |
| 2020-07-18 | 2467 | 2467 | 2467 | 2467 | 2467 |
| 2020-07-19 | 2541 | 2541 | 2541 | 2541 | 2541 |
| 2020-07-20 | 2616 | 2616 | 2616 | 2616 | 2616 |
| 2020-07-21 | 2692 | 2692 | 2692 | 2692 | 2692 |
| 2020-07-22 | 2768 | 2768 | 2768 | 2768 | 2768 |
| 2020-07-23 | 2846 | 2846 | 2846 | 2846 | 2846 |
| 2020-07-24 | 2922 | 2922 | 2922 | 2922 | 2922 |
| 2020-07-25 | 2999 | 2999 | 2999 | 2999 | 2999 |
| 2020-07-26 | 3073 | 3073 | 3073 | 3073 | 3073 |
| 2020-07-27 | 3147 | 3147 | 3147 | 3147 | 3147 |
| 2020-07-28 | 3222 | 3222 | 3222 | 3222 | 3222 |
| 2020-07-29 | 3299 | 3299 | 3299 | 3299 | 3299 |
| 2020-07-30 | 3376 | 3376 | 3376 | 3376 | 3376 |
| 2020-07-31 | 3453 | 3453 | 3453 | 3453 | 3453 |
| 2020-08-01 | 3528 | 3528 | 3528 | 3528 | 3528 |
| 2020-08-02 | 3605 | 3605 | 3605 | 3605 | 3605 |
| 2020-08-03 | 3682 | 3682 | 3682 | 3682 | 3682 |
| 2020-08-04 | 3757 | 3757 | 3757 | 3757 | 3757 |
| 2020-08-05 | 3832 | 3832 | 3832 | 3832 | 3832 |
| 2020-08-06 | 3908 | 3908 | 3908 | 3908 | 3908 |
| 2020-08-07 | 3983 | 3983 | 3983 | 3983 | 3983 |
| 2020-08-08 | 4060 | 4060 | 4060 | 4060 | 4060 |
| 2020-08-09 | 4137 | 4137 | 4137 | 4137 | 4137 |
| 2020-08-10 | 4213 | 4213 | 4213 | 4213 | 4213 |
| 2020-08-11 | 4290 | 4290 | 4290 | 4290 | 4290 |
| 2020-08-12 | 4364 | 4364 | 4364 | 4364 | 4364 |
| 2020-08-13 | 4439 | 4439 | 4439 | 4439 | 4439 |
| 2020-08-14 | 4516 | 4516 | 4516 | 4516 | 4516 |
| 2020-08-15 | 4592 | 4592 | 4592 | 4592 | 4592 |
| 2020-08-16 | 4669 | 4669 | 4669 | 4669 | 4669 |
| 2020-08-17 | 4747 | 4747 | 4747 | 4747 | 4747 |
| 2020-08-18 | 4823 | 4823 | 4823 | 4823 | 4823 |
| 2020-08-19 | 4900 | 4900 | 4900 | 4900 | 4900 |
| 2020-08-20 | 4977 | 4977 | 4977 | 4977 | 4977 |
| 2020-08-21 | 5055 | 5055 | 5055 | 5055 | 5055 |
| 2020-08-22 | 5132 | 5132 | 5132 | 5132 | 5132 |
| 2020-08-23 | 5210 | 5210 | 5210 | 5210 | 5210 |
| 2020-08-24 | 5288 | 5288 | 5288 | 5288 | 5288 |
| 2020-08-25 | 5366 | 5366 | 5366 | 5366 | 5366 |
| 2020-08-26 | 5443 | 5443 | 5443 | 5443 | 5443 |
| 2020-08-27 | 5520 | 5520 | 5520 | 5520 | 5520 |
| 2020-08-28 | 5597 | 5597 | 5597 | 5597 | 5597 |
| 2020-08-29 | 5675 | 5675 | 5675 | 5675 | 5675 |
| 2020-08-30 | 5751 | 5751 | 5751 | 5751 | 5751 |
| 2020-08-31 | 5829 | 5829 | 5829 | 5829 | 5829 |
| 2020-09-01 | 5906 | 5906 | 5906 | 5906 | 5906 |
| 2020-09-02 | 5983 | 5983 | 5983 | 5983 | 5983 |
| 2020-09-03 | 6061 | 6061 | 6061 | 6061 | 6061 |
| 2020-09-04 | 6137 | 6137 | 6137 | 6137 | 6137 |
| 2020-09-05 | 6213 | 6213 | 6213 | 6213 | 6213 |
| 2020-09-06 | 6290 | 6290 | 6290 | 6290 | 6290 |
| 2020-09-07 | 6368 | 6368 | 6368 | 6368 | 6368 |
| 2020-09-08 | 6445 | 6445 | 6445 | 6445 | 6445 |
| 2020-09-09 | 6523 | 6523 | 6523 | 6523 | 6523 |
| 2020-09-10 | 6601 | 6601 | 6601 | 6601 | 6601 |
| 2020-09-11 | 6676 | 6676 | 6676 | 6676 | 6676 |
| 2020-09-12 | 6754 | 6754 | 6754 | 6754 | 6754 |
| 2020-09-13 | 6831 | 6831 | 6831 | 6831 | 6831 |
| 2020-09-14 | 6908 | 6908 | 6908 | 6908 | 6908 |
| 2020-09-15 | 6985 | 6985 | 6985 | 6985 | 6985 |
| 2020-09-16 | 7075 | 7075 | 7075 | 7075 | 7075 |
| 2020-09-17 | 7164 | 7164 | 7164 | 7164 | 7164 |
| 2020-09-18 | 7254 | 7254 | 7254 | 7254 | 7254 |
| 2020-09-19 | 7346 | 7346 | 7346 | 7346 | 7346 |
| 2020-09-20 | 7439 | 7439 | 7439 | 7439 | 7439 |
| 2020-09-21 | 7533 | 7533 | 7533 | 7533 | 7533 |
| 2020-09-22 | 7631 | 7631 | 7631 | 7631 | 7631 |
| 2020-09-23 | 7731 | 7731 | 7731 | 7731 | 7731 |
| 2020-09-24 | 7833 | 7833 | 7833 | 7833 | 7833 |
| 2020-09-25 | 7940 | 7940 | 7940 | 7940 | 7940 |
| 2020-09-26 | 8048 | 8048 | 8048 | 8048 | 8048 |
| 2020-09-27 | 8157 | 8157 | 8157 | 8157 | 8157 |
| 2020-09-28 | 8270 | 8270 | 8270 | 8270 | 8270 |
| 2020-09-29 | 8386 | 8386 | 8386 | 8386 | 8386 |
| 2020-09-30 | 8504 | 8504 | 8504 | 8504 | 8504 |

c. Milwaukee, vaccination

Total number of confirmed cases

|  | 75% Adherence | 70% | 65% | 60% | Dynamic Adherence |
| --- | --- | --- | --- | --- | --- |
| 2020-03-15 | 0 | 0 | 0 | 0 | 0 |
| 2020-03-16 | 0 | 0 | 0 | 0 | 0 |
| 2020-03-17 | 0 | 0 | 0 | 0 | 0 |
| 2020-03-18 | 0 | 0 | 0 | 0 | 0 |
| 2020-03-19 | 3 | 3 | 3 | 3 | 3 |
| 2020-03-20 | 7 | 7 | 7 | 7 | 7 |
| 2020-03-21 | 12 | 12 | 12 | 12 | 12 |
| 2020-03-22 | 20 | 20 | 20 | 20 | 20 |
| 2020-03-23 | 28 | 28 | 28 | 28 | 28 |
| 2020-03-24 | 38 | 38 | 38 | 38 | 38 |
| 2020-03-25 | 51 | 51 | 51 | 51 | 51 |
| 2020-03-26 | 67 | 67 | 67 | 67 | 67 |
| 2020-03-27 | 88 | 88 | 88 | 88 | 88 |
| 2020-03-28 | 110 | 110 | 110 | 110 | 110 |
| 2020-03-29 | 140 | 140 | 140 | 140 | 140 |
| 2020-03-30 | 173 | 173 | 173 | 173 | 173 |
| 2020-03-31 | 212 | 212 | 212 | 212 | 212 |
| 2020-04-01 | 254 | 254 | 254 | 254 | 254 |
| 2020-04-02 | 302 | 302 | 302 | 302 | 302 |
| 2020-04-03 | 352 | 352 | 352 | 352 | 352 |
| 2020-04-04 | 404 | 404 | 404 | 404 | 404 |
| 2020-04-05 | 457 | 457 | 457 | 457 | 457 |
| 2020-04-06 | 509 | 509 | 509 | 509 | 509 |
| 2020-04-07 | 568 | 568 | 568 | 568 | 568 |
| 2020-04-08 | 631 | 631 | 631 | 631 | 631 |
| 2020-04-09 | 697 | 697 | 697 | 697 | 697 |
| 2020-04-10 | 769 | 769 | 769 | 769 | 769 |
| 2020-04-11 | 847 | 847 | 847 | 847 | 847 |
| 2020-04-12 | 925 | 925 | 925 | 925 | 925 |
| 2020-04-13 | 1007 | 1007 | 1007 | 1007 | 1007 |
| 2020-04-14 | 1096 | 1096 | 1096 | 1096 | 1096 |
| 2020-04-15 | 1185 | 1185 | 1185 | 1185 | 1185 |
| 2020-04-16 | 1282 | 1282 | 1282 | 1282 | 1282 |
| 2020-04-17 | 1382 | 1382 | 1382 | 1382 | 1382 |
| 2020-04-18 | 1489 | 1489 | 1489 | 1489 | 1489 |
| 2020-04-19 | 1597 | 1597 | 1597 | 1597 | 1597 |
| 2020-04-20 | 1712 | 1712 | 1712 | 1712 | 1712 |
| 2020-04-21 | 1836 | 1836 | 1836 | 1836 | 1836 |
| 2020-04-22 | 1966 | 1966 | 1966 | 1966 | 1966 |
| 2020-04-23 | 2037 | 2037 | 2037 | 2037 | 2037 |
| 2020-04-24 | 2114 | 2114 | 2114 | 2114 | 2114 |
| 2020-04-25 | 2191 | 2191 | 2191 | 2191 | 2191 |
| 2020-04-26 | 2270 | 2270 | 2270 | 2270 | 2270 |
| 2020-04-27 | 2350 | 2350 | 2350 | 2350 | 2350 |
| 2020-04-28 | 2461 | 2461 | 2461 | 2461 | 2461 |
| 2020-04-29 | 2567 | 2567 | 2567 | 2567 | 2567 |
| 2020-04-30 | 2668 | 2668 | 2668 | 2668 | 2668 |
| 2020-05-01 | 2766 | 2766 | 2766 | 2766 | 2766 |
| 2020-05-02 | 2861 | 2861 | 2861 | 2861 | 2861 |
| 2020-05-03 | 2950 | 2950 | 2950 | 2950 | 2950 |
| 2020-05-04 | 3045 | 3045 | 3045 | 3045 | 3045 |
| 2020-05-05 | 3140 | 3140 | 3140 | 3140 | 3140 |
| 2020-05-06 | 3236 | 3236 | 3236 | 3236 | 3236 |
| 2020-05-07 | 3333 | 3333 | 3333 | 3333 | 3333 |
| 2020-05-08 | 3433 | 3433 | 3433 | 3433 | 3433 |
| 2020-05-09 | 3531 | 3531 | 3531 | 3531 | 3531 |
| 2020-05-10 | 3628 | 3628 | 3628 | 3628 | 3628 |
| 2020-05-11 | 3726 | 3726 | 3726 | 3726 | 3726 |
| 2020-05-12 | 3827 | 3827 | 3827 | 3827 | 3827 |
| 2020-05-13 | 3924 | 3924 | 3924 | 3924 | 3924 |
| 2020-05-14 | 4023 | 4023 | 4023 | 4023 | 4023 |
| 2020-05-15 | 4123 | 4123 | 4123 | 4123 | 4123 |
| 2020-05-16 | 4226 | 4226 | 4226 | 4226 | 4226 |
| 2020-05-17 | 4325 | 4325 | 4325 | 4325 | 4325 |
| 2020-05-18 | 4426 | 4426 | 4426 | 4426 | 4426 |
| 2020-05-19 | 4524 | 4524 | 4524 | 4524 | 4524 |
| 2020-05-20 | 4629 | 4629 | 4629 | 4629 | 4629 |
| 2020-05-21 | 4745 | 4745 | 4745 | 4745 | 4745 |
| 2020-05-22 | 4857 | 4857 | 4857 | 4857 | 4857 |
| 2020-05-23 | 4974 | 4974 | 4974 | 4974 | 4974 |
| 2020-05-24 | 5090 | 5090 | 5090 | 5090 | 5090 |
| 2020-05-25 | 5208 | 5208 | 5208 | 5208 | 5208 |
| 2020-05-26 | 5323 | 5323 | 5323 | 5323 | 5323 |
| 2020-05-27 | 5438 | 5438 | 5438 | 5438 | 5438 |
| 2020-05-28 | 5551 | 5551 | 5551 | 5551 | 5551 |
| 2020-05-29 | 5701 | 5701 | 5701 | 5701 | 5701 |
| 2020-05-30 | 5852 | 5852 | 5852 | 5852 | 5852 |
| 2020-05-31 | 6002 | 6002 | 6002 | 6002 | 6002 |
| 2020-06-01 | 6160 | 6160 | 6160 | 6160 | 6160 |
| 2020-06-02 | 6324 | 6324 | 6324 | 6324 | 6324 |
| 2020-06-03 | 6493 | 6493 | 6493 | 6493 | 6493 |
| 2020-06-04 | 6675 | 6675 | 6675 | 6675 | 6675 |
| 2020-06-05 | 6863 | 6863 | 6863 | 6863 | 6863 |
| 2020-06-06 | 7062 | 7062 | 7062 | 7062 | 7062 |
| 2020-06-07 | 7270 | 7270 | 7270 | 7270 | 7270 |
| 2020-06-08 | 7492 | 7492 | 7492 | 7492 | 7492 |
| 2020-06-09 | 7722 | 7722 | 7722 | 7722 | 7722 |
| 2020-06-10 | 7906 | 7906 | 7906 | 7906 | 7906 |
| 2020-06-11 | 8093 | 8093 | 8093 | 8093 | 8093 |
| 2020-06-12 | 8291 | 8291 | 8291 | 8291 | 8291 |
| 2020-06-13 | 8491 | 8491 | 8491 | 8491 | 8491 |
| 2020-06-14 | 8694 | 8694 | 8694 | 8694 | 8694 |
| 2020-06-15 | 8899 | 8899 | 8899 | 8899 | 8899 |
| 2020-06-16 | 9104 | 9104 | 9104 | 9104 | 9104 |
| 2020-06-17 | 9312 | 9312 | 9312 | 9312 | 9312 |
| 2020-06-18 | 9518 | 9518 | 9518 | 9518 | 9518 |
| 2020-06-19 | 9720 | 9720 | 9720 | 9720 | 9720 |
| 2020-06-20 | 9918 | 9918 | 9918 | 9918 | 9918 |
| 2020-06-21 | 10123 | 10123 | 10123 | 10123 | 10123 |
| 2020-06-22 | 10330 | 10330 | 10330 | 10330 | 10330 |
| 2020-06-23 | 10535 | 10535 | 10535 | 10535 | 10535 |
| 2020-06-24 | 10738 | 10738 | 10738 | 10738 | 10738 |
| 2020-06-25 | 10943 | 10943 | 10943 | 10943 | 10943 |
| 2020-06-26 | 11151 | 11151 | 11151 | 11151 | 11151 |
| 2020-06-27 | 11354 | 11354 | 11354 | 11354 | 11354 |
| 2020-06-28 | 11557 | 11557 | 11557 | 11557 | 11557 |
| 2020-06-29 | 11758 | 11758 | 11758 | 11758 | 11758 |
| 2020-06-30 | 11964 | 11964 | 11964 | 11964 | 11964 |
| 2020-07-01 | 12171 | 12171 | 12171 | 12171 | 12171 |
| 2020-07-02 | 12375 | 12375 | 12375 | 12375 | 12375 |
| 2020-07-03 | 12578 | 12578 | 12578 | 12578 | 12578 |
| 2020-07-04 | 12786 | 12786 | 12786 | 12786 | 12786 |
| 2020-07-05 | 12997 | 12997 | 12997 | 12997 | 12997 |
| 2020-07-06 | 13204 | 13204 | 13204 | 13204 | 13204 |
| 2020-07-07 | 13412 | 13412 | 13412 | 13412 | 13412 |
| 2020-07-08 | 13615 | 13615 | 13615 | 13615 | 13615 |
| 2020-07-09 | 13815 | 13815 | 13815 | 13815 | 13815 |
| 2020-07-10 | 14024 | 14024 | 14024 | 14024 | 14024 |
| 2020-07-11 | 14231 | 14231 | 14231 | 14231 | 14231 |
| 2020-07-12 | 14440 | 14440 | 14440 | 14440 | 14440 |
| 2020-07-13 | 14652 | 14652 | 14652 | 14652 | 14652 |
| 2020-07-14 | 14860 | 14860 | 14860 | 14860 | 14860 |
| 2020-07-15 | 15064 | 15064 | 15064 | 15064 | 15064 |
| 2020-07-16 | 15271 | 15271 | 15271 | 15271 | 15271 |
| 2020-07-17 | 15477 | 15477 | 15477 | 15477 | 15477 |
| 2020-07-18 | 15685 | 15685 | 15685 | 15685 | 15685 |
| 2020-07-19 | 15891 | 15891 | 15891 | 15891 | 15891 |
| 2020-07-20 | 16098 | 16098 | 16098 | 16098 | 16098 |
| 2020-07-21 | 16308 | 16308 | 16308 | 16308 | 16308 |
| 2020-07-22 | 16518 | 16518 | 16518 | 16518 | 16518 |
| 2020-07-23 | 16727 | 16727 | 16727 | 16727 | 16727 |
| 2020-07-24 | 16936 | 16936 | 16936 | 16936 | 16936 |
| 2020-07-25 | 17146 | 17146 | 17146 | 17146 | 17146 |
| 2020-07-26 | 17355 | 17355 | 17355 | 17355 | 17355 |
| 2020-07-27 | 17563 | 17563 | 17563 | 17563 | 17563 |
| 2020-07-28 | 17774 | 17774 | 17774 | 17774 | 17774 |
| 2020-07-29 | 17979 | 17979 | 17979 | 17979 | 17979 |
| 2020-07-30 | 18187 | 18187 | 18187 | 18187 | 18187 |
| 2020-07-31 | 18431 | 18431 | 18431 | 18431 | 18431 |
| 2020-08-01 | 18677 | 18677 | 18677 | 18677 | 18677 |
| 2020-08-02 | 18917 | 18917 | 18917 | 18917 | 18917 |
| 2020-08-03 | 19160 | 19160 | 19160 | 19160 | 19160 |
| 2020-08-04 | 19411 | 19411 | 19411 | 19411 | 19411 |
| 2020-08-05 | 19667 | 19667 | 19667 | 19667 | 19667 |
| 2020-08-06 | 19933 | 19933 | 19933 | 19933 | 19933 |
| 2020-08-07 | 20201 | 20201 | 20201 | 20201 | 20201 |
| 2020-08-08 | 20475 | 20475 | 20475 | 20475 | 20475 |
| 2020-08-09 | 20757 | 20757 | 20757 | 20757 | 20757 |
| 2020-08-10 | 21046 | 21046 | 21046 | 21046 | 21046 |
| 2020-08-11 | 21343 | 21343 | 21343 | 21343 | 21343 |
| 2020-08-12 | 21639 | 21639 | 21639 | 21639 | 21639 |
| 2020-08-13 | 21954 | 21954 | 21954 | 21954 | 21954 |
| 2020-08-14 | 22270 | 22270 | 22270 | 22270 | 22270 |
| 2020-08-15 | 22596 | 22596 | 22596 | 22596 | 22596 |
| 2020-08-16 | 22926 | 22926 | 22926 | 22926 | 22926 |
| 2020-08-17 | 23265 | 23265 | 23265 | 23265 | 23265 |
| 2020-08-18 | 23622 | 23622 | 23622 | 23622 | 23622 |
| 2020-08-19 | 23977 | 23977 | 23977 | 23977 | 23977 |
| 2020-08-20 | 24345 | 24345 | 24345 | 24345 | 24345 |
| 2020-08-21 | 24720 | 24720 | 24720 | 24720 | 24720 |
| 2020-08-22 | 25106 | 25106 | 25106 | 25106 | 25106 |
| 2020-08-23 | 25498 | 25498 | 25498 | 25498 | 25498 |
| 2020-08-24 | 25899 | 25899 | 25899 | 25899 | 25899 |
| 2020-08-25 | 26306 | 26306 | 26306 | 26306 | 26306 |
| 2020-08-26 | 26718 | 26718 | 26718 | 26718 | 26718 |
| 2020-08-27 | 27146 | 27146 | 27146 | 27146 | 27146 |
| 2020-08-28 | 27590 | 27590 | 27590 | 27590 | 27590 |
| 2020-08-29 | 28034 | 28034 | 28034 | 28034 | 28034 |
| 2020-08-30 | 28423 | 28423 | 28423 | 28423 | 28423 |
| 2020-08-31 | 28826 | 28826 | 28826 | 28826 | 28826 |
| 2020-09-01 | 29237 | 29237 | 29237 | 29237 | 29237 |
| 2020-09-02 | 29639 | 29639 | 29639 | 29639 | 29639 |
| 2020-09-03 | 30053 | 30053 | 30053 | 30053 | 30053 |
| 2020-09-04 | 30465 | 30465 | 30465 | 30465 | 30465 |
| 2020-09-05 | 30875 | 30875 | 30875 | 30875 | 30875 |
| 2020-09-06 | 31282 | 31282 | 31282 | 31282 | 31282 |
| 2020-09-07 | 31692 | 31692 | 31692 | 31692 | 31692 |
| 2020-09-08 | 32099 | 32099 | 32099 | 32099 | 32099 |
| 2020-09-09 | 32496 | 32496 | 32496 | 32496 | 32496 |
| 2020-09-10 | 32896 | 32896 | 32896 | 32896 | 32896 |
| 2020-09-11 | 33298 | 33298 | 33298 | 33298 | 33298 |
| 2020-09-12 | 33703 | 33703 | 33703 | 33703 | 33703 |
| 2020-09-13 | 34102 | 34102 | 34102 | 34102 | 34102 |
| 2020-09-14 | 34501 | 34501 | 34501 | 34501 | 34501 |
| 2020-09-15 | 34901 | 34901 | 34901 | 34901 | 34901 |
| 2020-09-16 | 35299 | 35299 | 35299 | 35299 | 35299 |
| 2020-09-17 | 35686 | 35686 | 35686 | 35686 | 35686 |
| 2020-09-18 | 36076 | 36076 | 36076 | 36076 | 36076 |
| 2020-09-19 | 36478 | 36478 | 36478 | 36478 | 36478 |
| 2020-09-20 | 36868 | 36868 | 36868 | 36868 | 36868 |
| 2020-09-21 | 37253 | 37253 | 37253 | 37253 | 37253 |
| 2020-09-22 | 37643 | 37643 | 37643 | 37643 | 37643 |
| 2020-09-23 | 38027 | 38027 | 38027 | 38027 | 38027 |
| 2020-09-24 | 38416 | 38416 | 38416 | 38416 | 38416 |
| 2020-09-25 | 38801 | 38801 | 38801 | 38801 | 38801 |
| 2020-09-26 | 39186 | 39186 | 39186 | 39186 | 39186 |
| 2020-09-27 | 39572 | 39572 | 39572 | 39572 | 39572 |
| 2020-09-28 | 39952 | 39952 | 39952 | 39952 | 39952 |
| 2020-09-29 | 40329 | 40329 | 40329 | 40329 | 40329 |
| 2020-09-30 | 40706 | 40706 | 40706 | 40706 | 40706 |

d. Milwaukee, no vaccination

Total number of confirmed cases

|  | 75% Adherence | 70% | 65% | 60% | Dynamic Adherence |
| --- | --- | --- | --- | --- | --- |
| 2020-03-15 | 0 | 0 | 0 | 0 | 0 |
| 2020-03-16 | 0 | 0 | 0 | 0 | 0 |
| 2020-03-17 | 0 | 0 | 0 | 0 | 0 |
| 2020-03-18 | 0 | 0 | 0 | 0 | 0 |
| 2020-03-19 | 3 | 3 | 3 | 3 | 3 |
| 2020-03-20 | 7 | 7 | 7 | 7 | 7 |
| 2020-03-21 | 12 | 12 | 12 | 12 | 12 |
| 2020-03-22 | 20 | 20 | 20 | 20 | 20 |
| 2020-03-23 | 28 | 28 | 28 | 28 | 28 |
| 2020-03-24 | 38 | 38 | 38 | 38 | 38 |
| 2020-03-25 | 51 | 51 | 51 | 51 | 51 |
| 2020-03-26 | 67 | 67 | 67 | 67 | 67 |
| 2020-03-27 | 88 | 88 | 88 | 88 | 88 |
| 2020-03-28 | 110 | 110 | 110 | 110 | 110 |
| 2020-03-29 | 140 | 140 | 140 | 140 | 140 |
| 2020-03-30 | 173 | 173 | 173 | 173 | 173 |
| 2020-03-31 | 212 | 212 | 212 | 212 | 212 |
| 2020-04-01 | 254 | 254 | 254 | 254 | 254 |
| 2020-04-02 | 302 | 302 | 302 | 302 | 302 |
| 2020-04-03 | 352 | 352 | 352 | 352 | 352 |
| 2020-04-04 | 404 | 404 | 404 | 404 | 404 |
| 2020-04-05 | 457 | 457 | 457 | 457 | 457 |
| 2020-04-06 | 509 | 509 | 509 | 509 | 509 |
| 2020-04-07 | 568 | 568 | 568 | 568 | 568 |
| 2020-04-08 | 631 | 631 | 631 | 631 | 631 |
| 2020-04-09 | 697 | 697 | 697 | 697 | 697 |
| 2020-04-10 | 769 | 769 | 769 | 769 | 769 |
| 2020-04-11 | 847 | 847 | 847 | 847 | 847 |
| 2020-04-12 | 925 | 925 | 925 | 925 | 925 |
| 2020-04-13 | 1007 | 1007 | 1007 | 1007 | 1007 |
| 2020-04-14 | 1096 | 1096 | 1096 | 1096 | 1096 |
| 2020-04-15 | 1185 | 1185 | 1185 | 1185 | 1185 |
| 2020-04-16 | 1282 | 1282 | 1282 | 1282 | 1282 |
| 2020-04-17 | 1382 | 1382 | 1382 | 1382 | 1382 |
| 2020-04-18 | 1489 | 1489 | 1489 | 1489 | 1489 |
| 2020-04-19 | 1597 | 1597 | 1597 | 1597 | 1597 |
| 2020-04-20 | 1712 | 1712 | 1712 | 1712 | 1712 |
| 2020-04-21 | 1836 | 1836 | 1836 | 1836 | 1836 |
| 2020-04-22 | 1966 | 1966 | 1966 | 1966 | 1966 |
| 2020-04-23 | 2037 | 2037 | 2037 | 2037 | 2037 |
| 2020-04-24 | 2114 | 2114 | 2114 | 2114 | 2114 |
| 2020-04-25 | 2191 | 2191 | 2191 | 2191 | 2191 |
| 2020-04-26 | 2270 | 2270 | 2270 | 2270 | 2270 |
| 2020-04-27 | 2350 | 2350 | 2350 | 2350 | 2350 |
| 2020-04-28 | 2461 | 2461 | 2461 | 2461 | 2461 |
| 2020-04-29 | 2567 | 2567 | 2567 | 2567 | 2567 |
| 2020-04-30 | 2668 | 2668 | 2668 | 2668 | 2668 |
| 2020-05-01 | 2766 | 2766 | 2766 | 2766 | 2766 |
| 2020-05-02 | 2861 | 2861 | 2861 | 2861 | 2861 |
| 2020-05-03 | 2950 | 2950 | 2950 | 2950 | 2950 |
| 2020-05-04 | 3045 | 3045 | 3045 | 3045 | 3045 |
| 2020-05-05 | 3140 | 3140 | 3140 | 3140 | 3140 |
| 2020-05-06 | 3236 | 3236 | 3236 | 3236 | 3236 |
| 2020-05-07 | 3333 | 3333 | 3333 | 3333 | 3333 |
| 2020-05-08 | 3433 | 3433 | 3433 | 3433 | 3433 |
| 2020-05-09 | 3531 | 3531 | 3531 | 3531 | 3531 |
| 2020-05-10 | 3628 | 3628 | 3628 | 3628 | 3628 |
| 2020-05-11 | 3726 | 3726 | 3726 | 3726 | 3726 |
| 2020-05-12 | 3827 | 3827 | 3827 | 3827 | 3827 |
| 2020-05-13 | 3924 | 3924 | 3924 | 3924 | 3924 |
| 2020-05-14 | 4023 | 4023 | 4023 | 4023 | 4023 |
| 2020-05-15 | 4123 | 4123 | 4123 | 4123 | 4123 |
| 2020-05-16 | 4226 | 4226 | 4226 | 4226 | 4226 |
| 2020-05-17 | 4325 | 4325 | 4325 | 4325 | 4325 |
| 2020-05-18 | 4426 | 4426 | 4426 | 4426 | 4426 |
| 2020-05-19 | 4524 | 4524 | 4524 | 4524 | 4524 |
| 2020-05-20 | 4629 | 4629 | 4629 | 4629 | 4629 |
| 2020-05-21 | 4745 | 4745 | 4745 | 4745 | 4745 |
| 2020-05-22 | 4857 | 4857 | 4857 | 4857 | 4857 |
| 2020-05-23 | 4974 | 4974 | 4974 | 4974 | 4974 |
| 2020-05-24 | 5090 | 5090 | 5090 | 5090 | 5090 |
| 2020-05-25 | 5208 | 5208 | 5208 | 5208 | 5208 |
| 2020-05-26 | 5323 | 5323 | 5323 | 5323 | 5323 |
| 2020-05-27 | 5438 | 5438 | 5438 | 5438 | 5438 |
| 2020-05-28 | 5551 | 5551 | 5551 | 5551 | 5551 |
| 2020-05-29 | 5701 | 5701 | 5701 | 5701 | 5701 |
| 2020-05-30 | 5852 | 5852 | 5852 | 5852 | 5852 |
| 2020-05-31 | 6002 | 6002 | 6002 | 6002 | 6002 |
| 2020-06-01 | 6160 | 6160 | 6160 | 6160 | 6160 |
| 2020-06-02 | 6324 | 6324 | 6324 | 6324 | 6324 |
| 2020-06-03 | 6493 | 6493 | 6493 | 6493 | 6493 |
| 2020-06-04 | 6675 | 6675 | 6675 | 6675 | 6675 |
| 2020-06-05 | 6863 | 6863 | 6863 | 6863 | 6863 |
| 2020-06-06 | 7062 | 7062 | 7062 | 7062 | 7062 |
| 2020-06-07 | 7270 | 7270 | 7270 | 7270 | 7270 |
| 2020-06-08 | 7492 | 7492 | 7492 | 7492 | 7492 |
| 2020-06-09 | 7722 | 7722 | 7722 | 7722 | 7722 |
| 2020-06-10 | 7906 | 7906 | 7906 | 7906 | 7906 |
| 2020-06-11 | 8093 | 8093 | 8093 | 8093 | 8093 |
| 2020-06-12 | 8291 | 8291 | 8291 | 8291 | 8291 |
| 2020-06-13 | 8491 | 8491 | 8491 | 8491 | 8491 |
| 2020-06-14 | 8694 | 8694 | 8694 | 8694 | 8694 |
| 2020-06-15 | 8899 | 8899 | 8899 | 8899 | 8899 |
| 2020-06-16 | 9104 | 9104 | 9104 | 9104 | 9104 |
| 2020-06-17 | 9312 | 9312 | 9312 | 9312 | 9312 |
| 2020-06-18 | 9518 | 9518 | 9518 | 9518 | 9518 |
| 2020-06-19 | 9720 | 9720 | 9720 | 9720 | 9720 |
| 2020-06-20 | 9918 | 9918 | 9918 | 9918 | 9918 |
| 2020-06-21 | 10123 | 10123 | 10123 | 10123 | 10123 |
| 2020-06-22 | 10330 | 10330 | 10330 | 10330 | 10330 |
| 2020-06-23 | 10535 | 10535 | 10535 | 10535 | 10535 |
| 2020-06-24 | 10738 | 10738 | 10738 | 10738 | 10738 |
| 2020-06-25 | 10943 | 10943 | 10943 | 10943 | 10943 |
| 2020-06-26 | 11151 | 11151 | 11151 | 11151 | 11151 |
| 2020-06-27 | 11354 | 11354 | 11354 | 11354 | 11354 |
| 2020-06-28 | 11557 | 11557 | 11557 | 11557 | 11557 |
| 2020-06-29 | 11758 | 11758 | 11758 | 11758 | 11758 |
| 2020-06-30 | 11964 | 11964 | 11964 | 11964 | 11964 |
| 2020-07-01 | 12171 | 12171 | 12171 | 12171 | 12171 |
| 2020-07-02 | 12375 | 12375 | 12375 | 12375 | 12375 |
| 2020-07-03 | 12578 | 12578 | 12578 | 12578 | 12578 |
| 2020-07-04 | 12786 | 12786 | 12786 | 12786 | 12786 |
| 2020-07-05 | 12997 | 12997 | 12997 | 12997 | 12997 |
| 2020-07-06 | 13204 | 13204 | 13204 | 13204 | 13204 |
| 2020-07-07 | 13412 | 13412 | 13412 | 13412 | 13412 |
| 2020-07-08 | 13615 | 13615 | 13615 | 13615 | 13615 |
| 2020-07-09 | 13815 | 13815 | 13815 | 13815 | 13815 |
| 2020-07-10 | 14024 | 14024 | 14024 | 14024 | 14024 |
| 2020-07-11 | 14231 | 14231 | 14231 | 14231 | 14231 |
| 2020-07-12 | 14440 | 14440 | 14440 | 14440 | 14440 |
| 2020-07-13 | 14652 | 14652 | 14652 | 14652 | 14652 |
| 2020-07-14 | 14860 | 14860 | 14860 | 14860 | 14860 |
| 2020-07-15 | 15064 | 15064 | 15064 | 15064 | 15064 |
| 2020-07-16 | 15271 | 15271 | 15271 | 15271 | 15271 |
| 2020-07-17 | 15477 | 15477 | 15477 | 15477 | 15477 |
| 2020-07-18 | 15685 | 15685 | 15685 | 15685 | 15685 |
| 2020-07-19 | 15891 | 15891 | 15891 | 15891 | 15891 |
| 2020-07-20 | 16098 | 16098 | 16098 | 16098 | 16098 |
| 2020-07-21 | 16308 | 16308 | 16308 | 16308 | 16308 |
| 2020-07-22 | 16518 | 16518 | 16518 | 16518 | 16518 |
| 2020-07-23 | 16727 | 16727 | 16727 | 16727 | 16727 |
| 2020-07-24 | 16936 | 16936 | 16936 | 16936 | 16936 |
| 2020-07-25 | 17146 | 17146 | 17146 | 17146 | 17146 |
| 2020-07-26 | 17355 | 17355 | 17355 | 17355 | 17355 |
| 2020-07-27 | 17563 | 17563 | 17563 | 17563 | 17563 |
| 2020-07-28 | 17774 | 17774 | 17774 | 17774 | 17774 |
| 2020-07-29 | 17979 | 17979 | 17979 | 17979 | 17979 |
| 2020-07-30 | 18187 | 18187 | 18187 | 18187 | 18187 |
| 2020-07-31 | 18431 | 18431 | 18431 | 18431 | 18431 |
| 2020-08-01 | 18677 | 18677 | 18677 | 18677 | 18677 |
| 2020-08-02 | 18917 | 18917 | 18917 | 18917 | 18917 |
| 2020-08-03 | 19160 | 19160 | 19160 | 19160 | 19160 |
| 2020-08-04 | 19411 | 19411 | 19411 | 19411 | 19411 |
| 2020-08-05 | 19667 | 19667 | 19667 | 19667 | 19667 |
| 2020-08-06 | 19933 | 19933 | 19933 | 19933 | 19933 |
| 2020-08-07 | 20201 | 20201 | 20201 | 20201 | 20201 |
| 2020-08-08 | 20475 | 20475 | 20475 | 20475 | 20475 |
| 2020-08-09 | 20757 | 20757 | 20757 | 20757 | 20757 |
| 2020-08-10 | 21046 | 21046 | 21046 | 21046 | 21046 |
| 2020-08-11 | 21343 | 21343 | 21343 | 21343 | 21343 |
| 2020-08-12 | 21639 | 21639 | 21639 | 21639 | 21639 |
| 2020-08-13 | 21954 | 21954 | 21954 | 21954 | 21954 |
| 2020-08-14 | 22270 | 22270 | 22270 | 22270 | 22270 |
| 2020-08-15 | 22596 | 22596 | 22596 | 22596 | 22596 |
| 2020-08-16 | 22926 | 22926 | 22926 | 22926 | 22926 |
| 2020-08-17 | 23265 | 23265 | 23265 | 23265 | 23265 |
| 2020-08-18 | 23622 | 23622 | 23622 | 23622 | 23622 |
| 2020-08-19 | 23977 | 23977 | 23977 | 23977 | 23977 |
| 2020-08-20 | 24345 | 24345 | 24345 | 24345 | 24345 |
| 2020-08-21 | 24720 | 24720 | 24720 | 24720 | 24720 |
| 2020-08-22 | 25106 | 25106 | 25106 | 25106 | 25106 |
| 2020-08-23 | 25498 | 25498 | 25498 | 25498 | 25498 |
| 2020-08-24 | 25899 | 25899 | 25899 | 25899 | 25899 |
| 2020-08-25 | 26306 | 26306 | 26306 | 26306 | 26306 |
| 2020-08-26 | 26718 | 26718 | 26718 | 26718 | 26718 |
| 2020-08-27 | 27146 | 27146 | 27146 | 27146 | 27146 |
| 2020-08-28 | 27590 | 27590 | 27590 | 27590 | 27590 |
| 2020-08-29 | 28034 | 28034 | 28034 | 28034 | 28034 |
| 2020-08-30 | 28423 | 28423 | 28423 | 28423 | 28423 |
| 2020-08-31 | 28826 | 28826 | 28826 | 28826 | 28826 |
| 2020-09-01 | 29237 | 29237 | 29237 | 29237 | 29237 |
| 2020-09-02 | 29639 | 29639 | 29639 | 29639 | 29639 |
| 2020-09-03 | 30053 | 30053 | 30053 | 30053 | 30053 |
| 2020-09-04 | 30465 | 30465 | 30465 | 30465 | 30465 |
| 2020-09-05 | 30875 | 30875 | 30875 | 30875 | 30875 |
| 2020-09-06 | 31282 | 31282 | 31282 | 31282 | 31282 |
| 2020-09-07 | 31692 | 31692 | 31692 | 31692 | 31692 |
| 2020-09-08 | 32099 | 32099 | 32099 | 32099 | 32099 |
| 2020-09-09 | 32496 | 32496 | 32496 | 32496 | 32496 |
| 2020-09-10 | 32896 | 32896 | 32896 | 32896 | 32896 |
| 2020-09-11 | 33298 | 33298 | 33298 | 33298 | 33298 |
| 2020-09-12 | 33703 | 33703 | 33703 | 33703 | 33703 |
| 2020-09-13 | 34102 | 34102 | 34102 | 34102 | 34102 |
| 2020-09-14 | 34501 | 34501 | 34501 | 34501 | 34501 |
| 2020-09-15 | 34901 | 34901 | 34901 | 34901 | 34901 |
| 2020-09-16 | 35299 | 35299 | 35299 | 35299 | 35299 |
| 2020-09-17 | 35686 | 35686 | 35686 | 35686 | 35686 |
| 2020-09-18 | 36076 | 36076 | 36076 | 36076 | 36076 |
| 2020-09-19 | 36478 | 36478 | 36478 | 36478 | 36478 |
| 2020-09-20 | 36868 | 36868 | 36868 | 36868 | 36868 |
| 2020-09-21 | 37253 | 37253 | 37253 | 37253 | 37253 |
| 2020-09-22 | 37643 | 37643 | 37643 | 37643 | 37643 |
| 2020-09-23 | 38027 | 38027 | 38027 | 38027 | 38027 |
| 2020-09-24 | 38416 | 38416 | 38416 | 38416 | 38416 |
| 2020-09-25 | 38801 | 38801 | 38801 | 38801 | 38801 |
| 2020-09-26 | 39186 | 39186 | 39186 | 39186 | 39186 |
| 2020-09-27 | 39572 | 39572 | 39572 | 39572 | 39572 |
| 2020-09-28 | 39952 | 39952 | 39952 | 39952 | 39952 |
| 2020-09-29 | 40329 | 40329 | 40329 | 40329 | 40329 |
| 2020-09-30 | 40706 | 40706 | 40706 | 40706 | 40706 |

e. NYC, vaccination

Total number of confirmed cases

|  | 90% Adherence | 85% | 80% | 75% | Dynamic Adherence |
| --- | --- | --- | --- | --- | --- |
| 2020-03-15 | 0 | 0 | 0 | 0 | 0 |
| 2020-03-16 | 0 | 0 | 0 | 0 | 0 |
| 2020-03-17 | 0 | 0 | 0 | 0 | 0 |
| 2020-03-18 | 0 | 0 | 0 | 0 | 0 |
| 2020-03-19 | 16 | 16 | 16 | 16 | 16 |
| 2020-03-20 | 136 | 136 | 136 | 136 | 136 |
| 2020-03-21 | 256 | 256 | 256 | 256 | 256 |
| 2020-03-22 | 398 | 398 | 398 | 398 | 398 |
| 2020-03-23 | 635 | 635 | 635 | 635 | 635 |
| 2020-03-24 | 970 | 970 | 970 | 970 | 970 |
| 2020-03-25 | 1416 | 1416 | 1416 | 1416 | 1416 |
| 2020-03-26 | 2088 | 2088 | 2088 | 2088 | 2088 |
| 2020-03-27 | 3019 | 3019 | 3019 | 3019 | 3019 |
| 2020-03-28 | 4209 | 4209 | 4209 | 4209 | 4209 |
| 2020-03-29 | 5744 | 5744 | 5744 | 5744 | 5744 |
| 2020-03-30 | 7623 | 7623 | 7623 | 7623 | 7623 |
| 2020-03-31 | 9794 | 9794 | 9794 | 9794 | 9794 |
| 2020-04-01 | 12226 | 12226 | 12226 | 12226 | 12226 |
| 2020-04-02 | 14803 | 14803 | 14803 | 14803 | 14803 |
| 2020-04-03 | 17237 | 17237 | 17237 | 17237 | 17237 |
| 2020-04-04 | 19264 | 19264 | 19264 | 19264 | 19264 |
| 2020-04-05 | 21702 | 21702 | 21702 | 21702 | 21702 |
| 2020-04-06 | 24464 | 24464 | 24464 | 24464 | 24464 |
| 2020-04-07 | 27356 | 27356 | 27356 | 27356 | 27356 |
| 2020-04-08 | 30399 | 30399 | 30399 | 30399 | 30399 |
| 2020-04-09 | 33612 | 33612 | 33612 | 33612 | 33612 |
| 2020-04-10 | 36926 | 36926 | 36926 | 36926 | 36926 |
| 2020-04-11 | 40383 | 40383 | 40383 | 40383 | 40383 |
| 2020-04-12 | 43962 | 43962 | 43962 | 43962 | 43962 |
| 2020-04-13 | 47704 | 47704 | 47704 | 47704 | 47704 |
| 2020-04-14 | 51558 | 51558 | 51558 | 51558 | 51558 |
| 2020-04-15 | 55724 | 55724 | 55724 | 55724 | 55724 |
| 2020-04-16 | 60041 | 60041 | 60041 | 60041 | 60041 |
| 2020-04-17 | 64551 | 64551 | 64551 | 64551 | 64551 |
| 2020-04-18 | 69293 | 69293 | 69293 | 69293 | 69293 |
| 2020-04-19 | 74174 | 74174 | 74174 | 74174 | 74174 |
| 2020-04-20 | 79294 | 79294 | 79294 | 79294 | 79294 |
| 2020-04-21 | 84677 | 84677 | 84677 | 84677 | 84677 |
| 2020-04-22 | 90188 | 90188 | 90188 | 90188 | 90188 |
| 2020-04-23 | 96006 | 96006 | 96006 | 96006 | 96006 |
| 2020-04-24 | 102121 | 102121 | 102121 | 102121 | 102121 |
| 2020-04-25 | 108476 | 108476 | 108476 | 108476 | 108476 |
| 2020-04-26 | 115583 | 115583 | 115583 | 115583 | 115583 |
| 2020-04-27 | 122960 | 122960 | 122960 | 122960 | 122960 |
| 2020-04-28 | 126975 | 126975 | 126975 | 126975 | 126975 |
| 2020-04-29 | 131186 | 131186 | 131186 | 131186 | 131186 |
| 2020-04-30 | 135585 | 135585 | 135585 | 135585 | 135585 |
| 2020-05-01 | 139848 | 139848 | 139848 | 139848 | 139848 |
| 2020-05-02 | 143941 | 143941 | 143941 | 143941 | 143941 |
| 2020-05-03 | 147885 | 147885 | 147885 | 147885 | 147885 |
| 2020-05-04 | 151528 | 151528 | 151528 | 151528 | 151528 |
| 2020-05-05 | 154909 | 154909 | 154909 | 154909 | 154909 |
| 2020-05-06 | 158114 | 158114 | 158114 | 158114 | 158114 |
| 2020-05-07 | 161007 | 161007 | 161007 | 161007 | 161007 |
| 2020-05-08 | 163601 | 163601 | 163601 | 163601 | 163601 |
| 2020-05-09 | 166148 | 166148 | 166148 | 166148 | 166148 |
| 2020-05-10 | 168612 | 168612 | 168612 | 168612 | 168612 |
| 2020-05-11 | 170889 | 170889 | 170889 | 170889 | 170889 |
| 2020-05-12 | 173053 | 173053 | 173053 | 173053 | 173053 |
| 2020-05-13 | 175078 | 175078 | 175078 | 175078 | 175078 |
| 2020-05-14 | 176994 | 176994 | 176994 | 176994 | 176994 |
| 2020-05-15 | 178782 | 178782 | 178782 | 178782 | 178782 |
| 2020-05-16 | 180446 | 180446 | 180446 | 180446 | 180446 |
| 2020-05-17 | 182009 | 182009 | 182009 | 182009 | 182009 |
| 2020-05-18 | 183465 | 183465 | 183465 | 183465 | 183465 |
| 2020-05-19 | 184859 | 184859 | 184859 | 184859 | 184859 |
| 2020-05-20 | 186173 | 186173 | 186173 | 186173 | 186173 |
| 2020-05-21 | 187546 | 187546 | 187546 | 187546 | 187546 |
| 2020-05-22 | 188824 | 188824 | 188824 | 188824 | 188824 |
| 2020-05-23 | 190036 | 190036 | 190036 | 190036 | 190036 |
| 2020-05-24 | 191206 | 191206 | 191206 | 191206 | 191206 |
| 2020-05-25 | 192290 | 192290 | 192290 | 192290 | 192290 |
| 2020-05-26 | 193291 | 193291 | 193291 | 193291 | 193291 |
| 2020-05-27 | 194217 | 194217 | 194217 | 194217 | 194217 |
| 2020-05-28 | 195088 | 195088 | 195088 | 195088 | 195088 |
| 2020-05-29 | 195887 | 195887 | 195887 | 195887 | 195887 |
| 2020-05-30 | 196657 | 196657 | 196657 | 196657 | 196657 |
| 2020-05-31 | 197411 | 197411 | 197411 | 197411 | 197411 |
| 2020-06-01 | 198095 | 198095 | 198095 | 198095 | 198095 |
| 2020-06-02 | 198759 | 198759 | 198759 | 198759 | 198759 |
| 2020-06-03 | 199378 | 199378 | 199378 | 199378 | 199378 |
| 2020-06-04 | 199977 | 199977 | 199977 | 199977 | 199977 |
| 2020-06-05 | 200537 | 200537 | 200537 | 200537 | 200537 |
| 2020-06-06 | 201048 | 201048 | 201048 | 201048 | 201048 |
| 2020-06-07 | 201534 | 201534 | 201534 | 201534 | 201534 |
| 2020-06-08 | 201998 | 201998 | 201998 | 201998 | 201998 |
| 2020-06-09 | 202433 | 202433 | 202433 | 202433 | 202433 |
| 2020-06-10 | 202856 | 202856 | 202856 | 202856 | 202856 |
| 2020-06-11 | 203261 | 203261 | 203261 | 203261 | 203261 |
| 2020-06-12 | 203643 | 203643 | 203643 | 203643 | 203643 |
| 2020-06-13 | 204004 | 204004 | 204004 | 204004 | 204004 |
| 2020-06-14 | 204348 | 204348 | 204348 | 204348 | 204348 |
| 2020-06-15 | 204669 | 204669 | 204669 | 204669 | 204669 |
| 2020-06-16 | 204976 | 204976 | 204976 | 204976 | 204976 |
| 2020-06-17 | 205263 | 205263 | 205263 | 205263 | 205263 |
| 2020-06-18 | 205540 | 205540 | 205540 | 205540 | 205540 |
| 2020-06-19 | 205812 | 205812 | 205812 | 205812 | 205812 |
| 2020-06-20 | 206067 | 206067 | 206067 | 206067 | 206067 |
| 2020-06-21 | 206310 | 206310 | 206310 | 206310 | 206310 |
| 2020-06-22 | 206544 | 206544 | 206544 | 206544 | 206544 |
| 2020-06-23 | 206858 | 206858 | 206858 | 206858 | 206858 |
| 2020-06-24 | 207155 | 207155 | 207155 | 207155 | 207155 |
| 2020-06-25 | 207452 | 207452 | 207452 | 207452 | 207452 |
| 2020-06-26 | 207741 | 207741 | 207741 | 207741 | 207741 |
| 2020-06-27 | 208025 | 208025 | 208025 | 208025 | 208025 |
| 2020-06-28 | 208306 | 208306 | 208306 | 208306 | 208306 |
| 2020-06-29 | 208599 | 208599 | 208599 | 208599 | 208599 |
| 2020-06-30 | 208884 | 208884 | 208884 | 208884 | 208884 |
| 2020-07-01 | 209187 | 209187 | 209187 | 209187 | 209187 |
| 2020-07-02 | 209502 | 209502 | 209502 | 209502 | 209502 |
| 2020-07-03 | 209809 | 209809 | 209809 | 209809 | 209809 |
| 2020-07-04 | 210126 | 210126 | 210126 | 210126 | 210126 |
| 2020-07-05 | 210438 | 210438 | 210438 | 210438 | 210438 |
| 2020-07-06 | 210748 | 210748 | 210748 | 210748 | 210748 |
| 2020-07-07 | 211054 | 211054 | 211054 | 211054 | 211054 |
| 2020-07-08 | 211376 | 211376 | 211376 | 211376 | 211376 |
| 2020-07-09 | 211704 | 211704 | 211704 | 211704 | 211704 |
| 2020-07-10 | 212042 | 212042 | 212042 | 212042 | 212042 |
| 2020-07-11 | 212380 | 212380 | 212380 | 212380 | 212380 |
| 2020-07-12 | 212716 | 212716 | 212716 | 212716 | 212716 |
| 2020-07-13 | 213052 | 213052 | 213052 | 213052 | 213052 |
| 2020-07-14 | 213393 | 213393 | 213393 | 213393 | 213393 |
| 2020-07-15 | 213733 | 213733 | 213733 | 213733 | 213733 |
| 2020-07-16 | 214082 | 214082 | 214082 | 214082 | 214082 |
| 2020-07-17 | 214314 | 214314 | 214314 | 214314 | 214314 |
| 2020-07-18 | 214561 | 214561 | 214561 | 214561 | 214561 |
| 2020-07-19 | 214811 | 214811 | 214811 | 214811 | 214811 |
| 2020-07-20 | 215052 | 215052 | 215052 | 215052 | 215052 |
| 2020-07-21 | 215296 | 215296 | 215296 | 215296 | 215296 |
| 2020-07-22 | 215526 | 215526 | 215526 | 215526 | 215526 |
| 2020-07-23 | 215742 | 215742 | 215742 | 215742 | 215742 |
| 2020-07-24 | 215945 | 215945 | 215945 | 215945 | 215945 |
| 2020-07-25 | 216138 | 216138 | 216138 | 216138 | 216138 |
| 2020-07-26 | 216333 | 216333 | 216333 | 216333 | 216333 |
| 2020-07-27 | 216513 | 216513 | 216513 | 216513 | 216513 |
| 2020-07-28 | 216692 | 216692 | 216692 | 216692 | 216692 |
| 2020-07-29 | 216872 | 216872 | 216872 | 216872 | 216872 |
| 2020-07-30 | 217036 | 217036 | 217036 | 217036 | 217036 |
| 2020-07-31 | 217196 | 217196 | 217196 | 217196 | 217196 |
| 2020-08-01 | 217359 | 217359 | 217359 | 217359 | 217359 |
| 2020-08-02 | 217513 | 217513 | 217513 | 217513 | 217513 |
| 2020-08-03 | 217660 | 217660 | 217660 | 217660 | 217660 |
| 2020-08-04 | 217808 | 217808 | 217808 | 217808 | 217808 |
| 2020-08-05 | 217947 | 217947 | 217947 | 217947 | 217947 |
| 2020-08-06 | 218081 | 218081 | 218081 | 218081 | 218081 |
| 2020-08-07 | 218210 | 218210 | 218210 | 218210 | 218210 |
| 2020-08-08 | 218337 | 218337 | 218337 | 218337 | 218337 |
| 2020-08-09 | 218458 | 218458 | 218458 | 218458 | 218458 |
| 2020-08-10 | 218586 | 218586 | 218586 | 218586 | 218586 |
| 2020-08-11 | 218706 | 218706 | 218706 | 218706 | 218706 |
| 2020-08-12 | 218822 | 218822 | 218822 | 218822 | 218822 |
| 2020-08-13 | 218940 | 218940 | 218940 | 218940 | 218940 |
| 2020-08-14 | 219048 | 219048 | 219048 | 219048 | 219048 |
| 2020-08-15 | 219156 | 219156 | 219156 | 219156 | 219156 |
| 2020-08-16 | 219265 | 219265 | 219265 | 219265 | 219265 |
| 2020-08-17 | 219368 | 219368 | 219368 | 219368 | 219368 |
| 2020-08-18 | 219475 | 219475 | 219475 | 219475 | 219475 |
| 2020-08-19 | 219578 | 219578 | 219578 | 219578 | 219578 |
| 2020-08-20 | 219685 | 219685 | 219685 | 219685 | 219685 |
| 2020-08-21 | 219786 | 219786 | 219786 | 219786 | 219786 |
| 2020-08-22 | 219882 | 219882 | 219882 | 219882 | 219882 |
| 2020-08-23 | 219976 | 219976 | 219976 | 219976 | 219976 |
| 2020-08-24 | 220079 | 220079 | 220079 | 220079 | 220079 |
| 2020-08-25 | 220175 | 220175 | 220175 | 220175 | 220175 |
| 2020-08-26 | 220274 | 220274 | 220274 | 220274 | 220274 |
| 2020-08-27 | 220371 | 220371 | 220371 | 220371 | 220371 |
| 2020-08-28 | 220472 | 220472 | 220472 | 220472 | 220472 |
| 2020-08-29 | 220571 | 220571 | 220571 | 220571 | 220571 |
| 2020-08-30 | 220662 | 220662 | 220662 | 220662 | 220662 |
| 2020-08-31 | 220827 | 220827 | 220827 | 220827 | 220827 |
| 2020-09-01 | 220984 | 220984 | 220984 | 220984 | 220984 |
| 2020-09-02 | 221135 | 221135 | 221135 | 221135 | 221135 |
| 2020-09-03 | 221304 | 221304 | 221304 | 221304 | 221304 |
| 2020-09-04 | 221486 | 221486 | 221486 | 221486 | 221486 |
| 2020-09-05 | 221678 | 221678 | 221678 | 221678 | 221678 |
| 2020-09-06 | 221879 | 221879 | 221879 | 221879 | 221879 |
| 2020-09-07 | 222095 | 222095 | 222095 | 222095 | 222095 |
| 2020-09-08 | 222327 | 222327 | 222327 | 222327 | 222327 |
| 2020-09-09 | 222576 | 222576 | 222576 | 222576 | 222576 |
| 2020-09-10 | 222839 | 222839 | 222839 | 222839 | 222839 |
| 2020-09-11 | 223110 | 223110 | 223110 | 223110 | 223110 |
| 2020-09-12 | 223385 | 223385 | 223385 | 223385 | 223385 |
| 2020-09-13 | 223697 | 223697 | 223697 | 223697 | 223697 |
| 2020-09-14 | 224019 | 224019 | 224019 | 224019 | 224019 |
| 2020-09-15 | 224346 | 224346 | 224346 | 224346 | 224346 |
| 2020-09-16 | 224689 | 224689 | 224689 | 224689 | 224689 |
| 2020-09-17 | 225068 | 225068 | 225068 | 225068 | 225068 |
| 2020-09-18 | 225459 | 225459 | 225459 | 225459 | 225459 |
| 2020-09-19 | 225875 | 225875 | 225875 | 225875 | 225875 |
| 2020-09-20 | 226316 | 226316 | 226316 | 226316 | 226316 |
| 2020-09-21 | 226777 | 226777 | 226777 | 226777 | 226777 |
| 2020-09-22 | 227245 | 227245 | 227245 | 227245 | 227245 |
| 2020-09-23 | 227758 | 227758 | 227758 | 227758 | 227758 |
| 2020-09-24 | 228292 | 228292 | 228292 | 228292 | 228292 |
| 2020-09-25 | 228831 | 228831 | 228831 | 228831 | 228831 |
| 2020-09-26 | 229421 | 229421 | 229421 | 229421 | 229421 |
| 2020-09-27 | 230029 | 230029 | 230029 | 230029 | 230029 |
| 2020-09-28 | 230662 | 230662 | 230662 | 230662 | 230662 |
| 2020-09-29 | 231338 | 231338 | 231338 | 231338 | 231338 |
| 2020-09-30 | 231876 | 231876 | 231876 | 231876 | 231876 |

f. NYC, no vaccination

Total number of confirmed cases

|  | 90% Adherence | 85% | 80% | 75% | Dynamic Adherence |
| --- | --- | --- | --- | --- | --- |
| 2020-03-15 | 0 | 0 | 0 | 0 | 0 |
| 2020-03-16 | 0 | 0 | 0 | 0 | 0 |
| 2020-03-17 | 0 | 0 | 0 | 0 | 0 |
| 2020-03-18 | 0 | 0 | 0 | 0 | 0 |
| 2020-03-19 | 16 | 16 | 16 | 16 | 16 |
| 2020-03-20 | 136 | 136 | 136 | 136 | 136 |
| 2020-03-21 | 256 | 256 | 256 | 256 | 256 |
| 2020-03-22 | 398 | 398 | 398 | 398 | 398 |
| 2020-03-23 | 635 | 635 | 635 | 635 | 635 |
| 2020-03-24 | 970 | 970 | 970 | 970 | 970 |
| 2020-03-25 | 1416 | 1416 | 1416 | 1416 | 1416 |
| 2020-03-26 | 2088 | 2088 | 2088 | 2088 | 2088 |
| 2020-03-27 | 3019 | 3019 | 3019 | 3019 | 3019 |
| 2020-03-28 | 4209 | 4209 | 4209 | 4209 | 4209 |
| 2020-03-29 | 5744 | 5744 | 5744 | 5744 | 5744 |
| 2020-03-30 | 7623 | 7623 | 7623 | 7623 | 7623 |
| 2020-03-31 | 9794 | 9794 | 9794 | 9794 | 9794 |
| 2020-04-01 | 12226 | 12226 | 12226 | 12226 | 12226 |
| 2020-04-02 | 14803 | 14803 | 14803 | 14803 | 14803 |
| 2020-04-03 | 17237 | 17237 | 17237 | 17237 | 17237 |
| 2020-04-04 | 19264 | 19264 | 19264 | 19264 | 19264 |
| 2020-04-05 | 21702 | 21702 | 21702 | 21702 | 21702 |
| 2020-04-06 | 24464 | 24464 | 24464 | 24464 | 24464 |
| 2020-04-07 | 27356 | 27356 | 27356 | 27356 | 27356 |
| 2020-04-08 | 30399 | 30399 | 30399 | 30399 | 30399 |
| 2020-04-09 | 33612 | 33612 | 33612 | 33612 | 33612 |
| 2020-04-10 | 36926 | 36926 | 36926 | 36926 | 36926 |
| 2020-04-11 | 40383 | 40383 | 40383 | 40383 | 40383 |
| 2020-04-12 | 43962 | 43962 | 43962 | 43962 | 43962 |
| 2020-04-13 | 47704 | 47704 | 47704 | 47704 | 47704 |
| 2020-04-14 | 51558 | 51558 | 51558 | 51558 | 51558 |
| 2020-04-15 | 55724 | 55724 | 55724 | 55724 | 55724 |
| 2020-04-16 | 60041 | 60041 | 60041 | 60041 | 60041 |
| 2020-04-17 | 64551 | 64551 | 64551 | 64551 | 64551 |
| 2020-04-18 | 69293 | 69293 | 69293 | 69293 | 69293 |
| 2020-04-19 | 74174 | 74174 | 74174 | 74174 | 74174 |
| 2020-04-20 | 79294 | 79294 | 79294 | 79294 | 79294 |
| 2020-04-21 | 84677 | 84677 | 84677 | 84677 | 84677 |
| 2020-04-22 | 90188 | 90188 | 90188 | 90188 | 90188 |
| 2020-04-23 | 96006 | 96006 | 96006 | 96006 | 96006 |
| 2020-04-24 | 102121 | 102121 | 102121 | 102121 | 102121 |
| 2020-04-25 | 108476 | 108476 | 108476 | 108476 | 108476 |
| 2020-04-26 | 115583 | 115583 | 115583 | 115583 | 115583 |
| 2020-04-27 | 122960 | 122960 | 122960 | 122960 | 122960 |
| 2020-04-28 | 126975 | 126975 | 126975 | 126975 | 126975 |
| 2020-04-29 | 131186 | 131186 | 131186 | 131186 | 131186 |
| 2020-04-30 | 135585 | 135585 | 135585 | 135585 | 135585 |
| 2020-05-01 | 139848 | 139848 | 139848 | 139848 | 139848 |
| 2020-05-02 | 143941 | 143941 | 143941 | 143941 | 143941 |
| 2020-05-03 | 147885 | 147885 | 147885 | 147885 | 147885 |
| 2020-05-04 | 151528 | 151528 | 151528 | 151528 | 151528 |
| 2020-05-05 | 154909 | 154909 | 154909 | 154909 | 154909 |
| 2020-05-06 | 158114 | 158114 | 158114 | 158114 | 158114 |
| 2020-05-07 | 161007 | 161007 | 161007 | 161007 | 161007 |
| 2020-05-08 | 163601 | 163601 | 163601 | 163601 | 163601 |
| 2020-05-09 | 166148 | 166148 | 166148 | 166148 | 166148 |
| 2020-05-10 | 168612 | 168612 | 168612 | 168612 | 168612 |
| 2020-05-11 | 170889 | 170889 | 170889 | 170889 | 170889 |
| 2020-05-12 | 173053 | 173053 | 173053 | 173053 | 173053 |
| 2020-05-13 | 175078 | 175078 | 175078 | 175078 | 175078 |
| 2020-05-14 | 176994 | 176994 | 176994 | 176994 | 176994 |
| 2020-05-15 | 178782 | 178782 | 178782 | 178782 | 178782 |
| 2020-05-16 | 180446 | 180446 | 180446 | 180446 | 180446 |
| 2020-05-17 | 182009 | 182009 | 182009 | 182009 | 182009 |
| 2020-05-18 | 183465 | 183465 | 183465 | 183465 | 183465 |
| 2020-05-19 | 184859 | 184859 | 184859 | 184859 | 184859 |
| 2020-05-20 | 186173 | 186173 | 186173 | 186173 | 186173 |
| 2020-05-21 | 187546 | 187546 | 187546 | 187546 | 187546 |
| 2020-05-22 | 188824 | 188824 | 188824 | 188824 | 188824 |
| 2020-05-23 | 190036 | 190036 | 190036 | 190036 | 190036 |
| 2020-05-24 | 191206 | 191206 | 191206 | 191206 | 191206 |
| 2020-05-25 | 192290 | 192290 | 192290 | 192290 | 192290 |
| 2020-05-26 | 193291 | 193291 | 193291 | 193291 | 193291 |
| 2020-05-27 | 194217 | 194217 | 194217 | 194217 | 194217 |
| 2020-05-28 | 195088 | 195088 | 195088 | 195088 | 195088 |
| 2020-05-29 | 195887 | 195887 | 195887 | 195887 | 195887 |
| 2020-05-30 | 196657 | 196657 | 196657 | 196657 | 196657 |
| 2020-05-31 | 197411 | 197411 | 197411 | 197411 | 197411 |
| 2020-06-01 | 198095 | 198095 | 198095 | 198095 | 198095 |
| 2020-06-02 | 198759 | 198759 | 198759 | 198759 | 198759 |
| 2020-06-03 | 199378 | 199378 | 199378 | 199378 | 199378 |
| 2020-06-04 | 199977 | 199977 | 199977 | 199977 | 199977 |
| 2020-06-05 | 200537 | 200537 | 200537 | 200537 | 200537 |
| 2020-06-06 | 201048 | 201048 | 201048 | 201048 | 201048 |
| 2020-06-07 | 201534 | 201534 | 201534 | 201534 | 201534 |
| 2020-06-08 | 201998 | 201998 | 201998 | 201998 | 201998 |
| 2020-06-09 | 202433 | 202433 | 202433 | 202433 | 202433 |
| 2020-06-10 | 202856 | 202856 | 202856 | 202856 | 202856 |
| 2020-06-11 | 203261 | 203261 | 203261 | 203261 | 203261 |
| 2020-06-12 | 203643 | 203643 | 203643 | 203643 | 203643 |
| 2020-06-13 | 204004 | 204004 | 204004 | 204004 | 204004 |
| 2020-06-14 | 204348 | 204348 | 204348 | 204348 | 204348 |
| 2020-06-15 | 204669 | 204669 | 204669 | 204669 | 204669 |
| 2020-06-16 | 204976 | 204976 | 204976 | 204976 | 204976 |
| 2020-06-17 | 205263 | 205263 | 205263 | 205263 | 205263 |
| 2020-06-18 | 205540 | 205540 | 205540 | 205540 | 205540 |
| 2020-06-19 | 205812 | 205812 | 205812 | 205812 | 205812 |
| 2020-06-20 | 206067 | 206067 | 206067 | 206067 | 206067 |
| 2020-06-21 | 206310 | 206310 | 206310 | 206310 | 206310 |
| 2020-06-22 | 206544 | 206544 | 206544 | 206544 | 206544 |
| 2020-06-23 | 206858 | 206858 | 206858 | 206858 | 206858 |
| 2020-06-24 | 207155 | 207155 | 207155 | 207155 | 207155 |
| 2020-06-25 | 207452 | 207452 | 207452 | 207452 | 207452 |
| 2020-06-26 | 207741 | 207741 | 207741 | 207741 | 207741 |
| 2020-06-27 | 208025 | 208025 | 208025 | 208025 | 208025 |
| 2020-06-28 | 208306 | 208306 | 208306 | 208306 | 208306 |
| 2020-06-29 | 208599 | 208599 | 208599 | 208599 | 208599 |
| 2020-06-30 | 208884 | 208884 | 208884 | 208884 | 208884 |
| 2020-07-01 | 209187 | 209187 | 209187 | 209187 | 209187 |
| 2020-07-02 | 209502 | 209502 | 209502 | 209502 | 209502 |
| 2020-07-03 | 209809 | 209809 | 209809 | 209809 | 209809 |
| 2020-07-04 | 210126 | 210126 | 210126 | 210126 | 210126 |
| 2020-07-05 | 210438 | 210438 | 210438 | 210438 | 210438 |
| 2020-07-06 | 210748 | 210748 | 210748 | 210748 | 210748 |
| 2020-07-07 | 211054 | 211054 | 211054 | 211054 | 211054 |
| 2020-07-08 | 211376 | 211376 | 211376 | 211376 | 211376 |
| 2020-07-09 | 211704 | 211704 | 211704 | 211704 | 211704 |
| 2020-07-10 | 212042 | 212042 | 212042 | 212042 | 212042 |
| 2020-07-11 | 212380 | 212380 | 212380 | 212380 | 212380 |
| 2020-07-12 | 212716 | 212716 | 212716 | 212716 | 212716 |
| 2020-07-13 | 213052 | 213052 | 213052 | 213052 | 213052 |
| 2020-07-14 | 213393 | 213393 | 213393 | 213393 | 213393 |
| 2020-07-15 | 213733 | 213733 | 213733 | 213733 | 213733 |
| 2020-07-16 | 214082 | 214082 | 214082 | 214082 | 214082 |
| 2020-07-17 | 214314 | 214314 | 214314 | 214314 | 214314 |
| 2020-07-18 | 214561 | 214561 | 214561 | 214561 | 214561 |
| 2020-07-19 | 214811 | 214811 | 214811 | 214811 | 214811 |
| 2020-07-20 | 215052 | 215052 | 215052 | 215052 | 215052 |
| 2020-07-21 | 215296 | 215296 | 215296 | 215296 | 215296 |
| 2020-07-22 | 215526 | 215526 | 215526 | 215526 | 215526 |
| 2020-07-23 | 215742 | 215742 | 215742 | 215742 | 215742 |
| 2020-07-24 | 215945 | 215945 | 215945 | 215945 | 215945 |
| 2020-07-25 | 216138 | 216138 | 216138 | 216138 | 216138 |
| 2020-07-26 | 216333 | 216333 | 216333 | 216333 | 216333 |
| 2020-07-27 | 216513 | 216513 | 216513 | 216513 | 216513 |
| 2020-07-28 | 216692 | 216692 | 216692 | 216692 | 216692 |
| 2020-07-29 | 216872 | 216872 | 216872 | 216872 | 216872 |
| 2020-07-30 | 217036 | 217036 | 217036 | 217036 | 217036 |
| 2020-07-31 | 217196 | 217196 | 217196 | 217196 | 217196 |
| 2020-08-01 | 217359 | 217359 | 217359 | 217359 | 217359 |
| 2020-08-02 | 217513 | 217513 | 217513 | 217513 | 217513 |
| 2020-08-03 | 217660 | 217660 | 217660 | 217660 | 217660 |
| 2020-08-04 | 217808 | 217808 | 217808 | 217808 | 217808 |
| 2020-08-05 | 217947 | 217947 | 217947 | 217947 | 217947 |
| 2020-08-06 | 218081 | 218081 | 218081 | 218081 | 218081 |
| 2020-08-07 | 218210 | 218210 | 218210 | 218210 | 218210 |
| 2020-08-08 | 218337 | 218337 | 218337 | 218337 | 218337 |
| 2020-08-09 | 218458 | 218458 | 218458 | 218458 | 218458 |
| 2020-08-10 | 218586 | 218586 | 218586 | 218586 | 218586 |
| 2020-08-11 | 218706 | 218706 | 218706 | 218706 | 218706 |
| 2020-08-12 | 218822 | 218822 | 218822 | 218822 | 218822 |
| 2020-08-13 | 218940 | 218940 | 218940 | 218940 | 218940 |
| 2020-08-14 | 219048 | 219048 | 219048 | 219048 | 219048 |
| 2020-08-15 | 219156 | 219156 | 219156 | 219156 | 219156 |
| 2020-08-16 | 219265 | 219265 | 219265 | 219265 | 219265 |
| 2020-08-17 | 219368 | 219368 | 219368 | 219368 | 219368 |
| 2020-08-18 | 219475 | 219475 | 219475 | 219475 | 219475 |
| 2020-08-19 | 219578 | 219578 | 219578 | 219578 | 219578 |
| 2020-08-20 | 219685 | 219685 | 219685 | 219685 | 219685 |
| 2020-08-21 | 219786 | 219786 | 219786 | 219786 | 219786 |
| 2020-08-22 | 219882 | 219882 | 219882 | 219882 | 219882 |
| 2020-08-23 | 219976 | 219976 | 219976 | 219976 | 219976 |
| 2020-08-24 | 220079 | 220079 | 220079 | 220079 | 220079 |
| 2020-08-25 | 220175 | 220175 | 220175 | 220175 | 220175 |
| 2020-08-26 | 220274 | 220274 | 220274 | 220274 | 220274 |
| 2020-08-27 | 220371 | 220371 | 220371 | 220371 | 220371 |
| 2020-08-28 | 220472 | 220472 | 220472 | 220472 | 220472 |
| 2020-08-29 | 220571 | 220571 | 220571 | 220571 | 220571 |
| 2020-08-30 | 220662 | 220662 | 220662 | 220662 | 220662 |
| 2020-08-31 | 220827 | 220827 | 220827 | 220827 | 220827 |
| 2020-09-01 | 220984 | 220984 | 220984 | 220984 | 220984 |
| 2020-09-02 | 221135 | 221135 | 221135 | 221135 | 221135 |
| 2020-09-03 | 221304 | 221304 | 221304 | 221304 | 221304 |
| 2020-09-04 | 221486 | 221486 | 221486 | 221486 | 221486 |
| 2020-09-05 | 221678 | 221678 | 221678 | 221678 | 221678 |
| 2020-09-06 | 221879 | 221879 | 221879 | 221879 | 221879 |
| 2020-09-07 | 222095 | 222095 | 222095 | 222095 | 222095 |
| 2020-09-08 | 222327 | 222327 | 222327 | 222327 | 222327 |
| 2020-09-09 | 222576 | 222576 | 222576 | 222576 | 222576 |
| 2020-09-10 | 222839 | 222839 | 222839 | 222839 | 222839 |
| 2020-09-11 | 223110 | 223110 | 223110 | 223110 | 223110 |
| 2020-09-12 | 223385 | 223385 | 223385 | 223385 | 223385 |
| 2020-09-13 | 223697 | 223697 | 223697 | 223697 | 223697 |
| 2020-09-14 | 224019 | 224019 | 224019 | 224019 | 224019 |
| 2020-09-15 | 224346 | 224346 | 224346 | 224346 | 224346 |
| 2020-09-16 | 224689 | 224689 | 224689 | 224689 | 224689 |
| 2020-09-17 | 225068 | 225068 | 225068 | 225068 | 225068 |
| 2020-09-18 | 225459 | 225459 | 225459 | 225459 | 225459 |
| 2020-09-19 | 225875 | 225875 | 225875 | 225875 | 225875 |
| 2020-09-20 | 226316 | 226316 | 226316 | 226316 | 226316 |
| 2020-09-21 | 226777 | 226777 | 226777 | 226777 | 226777 |
| 2020-09-22 | 227245 | 227245 | 227245 | 227245 | 227245 |
| 2020-09-23 | 227758 | 227758 | 227758 | 227758 | 227758 |
| 2020-09-24 | 228292 | 228292 | 228292 | 228292 | 228292 |
| 2020-09-25 | 228831 | 228831 | 228831 | 228831 | 228831 |
| 2020-09-26 | 229421 | 229421 | 229421 | 229421 | 229421 |
| 2020-09-27 | 230029 | 230029 | 230029 | 230029 | 230029 |
| 2020-09-28 | 230662 | 230662 | 230662 | 230662 | 230662 |
| 2020-09-29 | 231338 | 231338 | 231338 | 231338 | 231338 |
| 2020-09-30 | 231876 | 231876 | 231876 | 231876 | 231876 |

#### Section C.2 Sensitivity analysis on the modeling of vaccination effect

COVAM assumes that the vaccination is effective by simply making a proportion of the vaccinated individuals becoming immune to SARS-CoV-2 therefore do not become contagious and therefore do not spread the virus to others. For example, 90% effectiveness implies that 9 out of 10 individuals become immune and 1 out of 10 remains susceptible. An alternative way of modeling effectiveness is to assume that vaccine does not eliminate the risk of infection for vaccinated individuals but reduces the probability of these individuals becoming infected by contagious individuals. In this alternative scenario, vaccinated individuals do not become completely immune, instead, their risk of being infected is reduced by 90%. Again, we conducted this experiment only for NYC. We did not change any parameters except this structural implementation.

Below, we first present the results of the validation, which show that COVAM was able to replicate the observed cumulative number of COVID-19 cases using the new set of values for adherence to NPIs. We then present the results of our experiments.

#### Supplement Figure 4. Comparison of model predictions to actual NYC data when vaccination effectiveness is modeled as a reduction in probability of becoming infected

Cumulative number of confiemed cases

|  | NYC- actual confirmed cases | Alagoz model results |
| --- | --- | --- |
| 2020-03-13 | 137 | 970 |
| 2020-03-14 | 185 | 1416 |
| 2020-03-15 | 269 | 2088 |
| 2020-03-16 | 464 | 3019 |
| 2020-03-17 | 923 | 4209 |
| 2020-03-18 | 2009 | 5744 |
| 2020-03-19 | 3954 | 7623 |
| 2020-03-20 | 5683 | 9794 |
| 2020-03-21 | 8115 | 12226 |
| 2020-03-22 | 10764 | 14803 |
| 2020-03-23 | 13119 | 17237 |
| 2020-03-24 | 15597 | 19264 |
| 2020-03-25 | 17856 | 21702 |
| 2020-03-26 | 23112 | 24464 |
| 2020-03-27 | 26697 | 27356 |
| 2020-03-28 | 30765 | 30399 |
| 2020-03-29 | 33474 | 33612 |
| 2020-03-30 | 38087 | 36926 |
| 2020-03-31 | 41771 | 40383 |
| 2020-04-01 | 47439 | 43962 |
| 2020-04-02 | 49707 | 47704 |
| 2020-04-03 | 57160 | 51558 |
| 2020-04-04 | 60850 | 55724 |
| 2020-04-05 | 64955 | 60041 |
| 2020-04-06 | 68776 | 64551 |
| 2020-04-07 | 74601 | 69293 |
| 2020-04-08 | 80204 | 74174 |
| 2020-04-09 | 87725 | 79294 |
| 2020-04-10 | 94409 | 84677 |
| 2020-04-11 | 98715 | 90188 |
| 2020-04-12 | 104410 | 96006 |
| 2020-04-13 | 107263 | 102121 |
| 2020-04-14 | 111424 | 108476 |
| 2020-04-15 | 117565 | 115583 |
| 2020-04-16 | 122148 | 122960 |
| 2020-04-17 | 126368 | 126975 |
| 2020-04-18 | 129788 | 131186 |
| 2020-04-19 | 132467 | 135585 |
| 2020-04-20 | 134874 | 139848 |
| 2020-04-21 | 138435 | 143941 |
| 2020-04-22 | 141754 | 147885 |
| 2020-04-23 | 146139 | 151528 |
| 2020-04-24 | 150576 | 154909 |
| 2020-04-25 | 153204 | 158114 |
| 2020-04-26 | 156100 | 161007 |
| 2020-04-27 | 157713 | 163601 |
| 2020-04-28 | 159865 | 166148 |
| 2020-04-29 | 162212 | 168612 |
| 2020-04-30 | 164505 | 170889 |
| 2020-05-01 | 166883 | 173053 |
| 2020-05-02 | 168845 | 175078 |
| 2020-05-03 | 170534 | 176994 |
| 2020-05-04 | 171723 | 178782 |
| 2020-05-05 | 173288 | 180446 |
| 2020-05-06 | 174709 | 182009 |
| 2020-05-07 | 176086 | 183465 |
| 2020-05-08 | 177481 | 184859 |
| 2020-05-09 | 178766 | 186173 |
| 2020-05-10 | 183662 | 187546 |
| 2020-05-11 | 184319 | 188824 |
| 2020-05-12 | 185206 | 190036 |
| 2020-05-13 | 186293 | 191206 |
| 2020-05-14 | 187848 | 192290 |
| 2020-05-15 | 189031 | 193291 |
| 2020-05-16 | 190408 | 194217 |
| 2020-05-17 | 191073 | 195088 |
| 2020-05-18 | 191650 | 195887 |
| 2020-05-19 | 192374 | 196657 |
| 2020-05-20 | 192840 | 197411 |
| 2020-05-21 | 193951 | 198095 |
| 2020-05-22 | 194667 | 198759 |
| 2020-05-23 | 195452 | 199378 |
| 2020-05-24 | 196098 | 199977 |
| 2020-05-25 | 196623 | 200537 |
| 2020-05-26 | 197351 | 201048 |
| 2020-05-27 | 198255 | 201534 |
| 2020-05-28 | 199038 | 201998 |
| 2020-05-29 | 199893 | 202433 |
| 2020-05-30 | 200547 | 202856 |
| 2020-05-31 | 200830 | 203261 |
| 2020-06-01 | 201123 | 203643 |
| 2020-06-02 | 201806 | 204004 |
| 2020-06-03 | 202319 | 204348 |
| 2020-06-04 | 202829 | 204669 |
| 2020-06-05 | 203430 | 204976 |
| 2020-06-06 | 203819 | 205263 |
| 2020-06-07 | 204253 | 205540 |
| 2020-06-08 | 204576 | 205812 |
| 2020-06-09 | 205011 | 206067 |
| 2020-06-10 | 205405 | 206310 |
| 2020-06-11 | 205846 | 206544 |
| 2020-06-12 | 206322 | 206858 |
| 2020-06-13 | 206606 | 207155 |
| 2020-06-14 | 207049 | 207452 |
| 2020-06-15 | 207373 | 207741 |
| 2020-06-16 | 207821 | 208025 |
| 2020-06-17 | 208097 | 208306 |
| 2020-06-18 | 208455 | 208599 |
| 2020-06-19 | 208763 | 208884 |
| 2020-06-20 | 209313 | 209187 |
| 2020-06-21 | 209562 | 209502 |
| 2020-06-22 | 209893 | 209809 |
| 2020-06-23 | 210185 | 210126 |
| 2020-06-24 | 210523 | 210438 |
| 2020-06-25 | 210908 | 210748 |
| 2020-06-26 | 211229 | 211054 |
| 2020-06-27 | 211569 | 211376 |
| 2020-06-28 | 211893 | 211704 |
| 2020-06-29 | 212072 | 212042 |
| 2020-06-30 | 212412 | 212380 |
| 2020-07-01 | 212774 | 212716 |
| 2020-07-02 | 213212 | 213052 |
| 2020-07-03 | 213561 | 213393 |
| 2020-07-04 | 213852 | 213733 |
| 2020-07-05 | 214061 | 214082 |
| 2020-07-06 | 214371 | 214314 |
| 2020-07-07 | 214570 | 214561 |
| 2020-07-08 | 214952 | 214811 |
| 2020-07-09 | 215265 | 215052 |
| 2020-07-10 | 215598 | 215296 |
| 2020-07-11 | 215924 | 215526 |
| 2020-07-12 | 216199 | 215742 |
| 2020-07-13 | 216468 | 215945 |
| 2020-07-14 | 216834 | 216138 |
| 2020-07-15 | 217230 | 216333 |
| 2020-07-16 | 217562 | 216513 |
| 2020-07-17 | 217895 | 216692 |
| 2020-07-18 | 218159 | 216872 |
| 2020-07-19 | 218478 | 217036 |
| 2020-07-20 | 219030 | 217196 |
| 2020-07-21 | 219128 | 217359 |
| 2020-07-22 | 219489 | 217513 |
| 2020-07-23 | 219641 | 217660 |
| 2020-07-24 | 220172 | 217808 |
| 2020-07-25 | 220266 | 217947 |
| 2020-07-26 | 220483 | 218081 |
| 2020-07-27 | 220907 | 218210 |
| 2020-07-28 | 221220 | 218337 |
| 2020-07-29 | 221534 | 218458 |
| 2020-07-30 | 221822 | 218586 |
| 2020-07-31 | 222131 | 218706 |
| 2020-08-01 | 222330 | 218822 |
| 2020-08-02 | 222522 | 218940 |
| 2020-08-03 | 222840 | 219048 |
| 2020-08-04 | 223186 | 219156 |
| 2020-08-05 | 223473 | 219265 |
| 2020-08-06 | 223876 | 219368 |
| 2020-08-07 | 224117 | 219475 |
| 2020-08-08 | 224438 | 219578 |
| 2020-08-09 | 224684 | 219685 |
| 2020-08-10 | 224920 | 219786 |
| 2020-08-11 | 225284 | 219882 |
| 2020-08-12 | 225713 | 219976 |
| 2020-08-13 | 226043 | 220079 |
| 2020-08-14 | 226367 | 220175 |
| 2020-08-15 | 226756 | 220274 |
| 2020-08-16 | 226977 | 220371 |
| 2020-08-17 | 227158 | 220472 |
| 2020-08-18 | 227419 | 220571 |
| 2020-08-19 | 227724 | 220662 |
| 2020-08-20 | 227927 | 220827 |
| 2020-08-21 | 228144 | 220984 |
| 2020-08-22 | 228428 | 221135 |
| 2020-08-23 | 228617 | 221304 |
| 2020-08-24 | 228788 | 221486 |
| 2020-08-25 | 228971 | 221678 |
| 2020-08-26 | 229207 | 221879 |
| 2020-08-27 | 229518 | 222095 |
| 2020-08-28 | 229751 | 222327 |
| 2020-08-29 | 229980 | 222576 |
| 2020-08-30 | 230205 | 222839 |
| 2020-08-31 | 230496 | 223110 |
| 2020-09-01 | 230744 | 223385 |
| 2020-09-02 | 231068 | 223697 |
| 2020-09-03 | 231290 | 224019 |
| 2020-09-04 | 231594 | 224346 |
| 2020-09-05 |  | 224689 |
| 2020-09-06 | 231824 | 225068 |
| 2020-09-07 | 231594 | 225459 |
| 2020-09-08 | 232232 | 225875 |
| 2020-09-09 | 232710 | 226316 |
| 2020-09-10 | 232926 | 226777 |
| 2020-09-11 | 233216 | 227245 |
| 2020-09-12 |  | 227758 |
| 2020-09-13 | 233636 | 228292 |
| 2020-09-14 | 233972 | 228831 |
| 2020-09-15 | 234225 | 229421 |
| 2020-09-16 | 234500 | 230029 |
| 2020-09-17 | 234827 | 230662 |
| 2020-09-18 | 235461 | 231338 |
| 2020-09-19 | 235649 | 231876 |
| 2020-09-20 | 235969 | 232440 |
| 2020-09-21 | 236253 | 233016 |
| 2020-09-22 | 236462 | 233610 |
| 2020-09-23 | 236841 | 234211 |
| 2020-09-24 | 237227 | 234809 |
| 2020-09-25 | 237628 | 235405 |
| 2020-09-26 | 237971 | 236017 |
| 2020-09-27 | 238338 | 236615 |
| 2020-09-28 | 238733 | 237225 |

Supplement Table 4: Controllable spread date and number of cases on December 31, 2021 for different daily vaccination capacity scenarios when vaccination effectiveness is modeled as a reduction in probability of becoming infected (vaccine effectiveness 90%, vaccine coverage 50%). Numbers in parentheses represent percent reduction relative to no vaccine.

| NYC | | | | | | | | | | | |
| --- | --- | --- | --- | --- | --- | --- | --- | --- | --- | --- | --- |
|  | 90% Adherence | | | 85% Adherence | | 80% Adherence | | 75% Adherence | | Dynamic Adherence | |
| Vaccination capacity | Date | Number of cases | Date | | Number of cases | Date | Number of cases | Date | Number of cases | Date | Number of cases |
| No Vaccine | 22-Mar-2021 | 699,352 | 27-Aug-2021 | | 1,110,070 | 4-Sep-2021 | 3,307,090 | 21-Jun-2021 | 4,999,100 | After June 2022 | 1,308,990 |
| 0.05% | 20-Mar-2021 | 686,086  (2%) | 25-Jun-2021 | | 986,528  (11%) | 20-Aug-2021 | 2,764,620  (16%) | 21-Jun-2021 | 4,570,160  (9%) | After June 2022 | 1,273,190  (3%) |
| 0.1% | 17-Mar-2021 | 674,462  (4%) | 30-May-2021 | | 909,986  (18%) | 3-Aug-2021 | 2,287,610  (31%) | 19-Jun-2021 | 4,132,220  (17%) | 9-Feb-2022 | 1,193,500  (9%) |
| 0.25% | 11-Mar-2021 | 647,315  (7%) | 23-Apr-2021 | | 784,676  (29%) | 15-Jun-2021 | 1,385,800  (58%) | 11-Jun-2021 | 2,839,570  (43%) | 28-Jul-2021 | 873,254  (33%) |
| 0.5% | 3-Mar-2021 | 615,607  (12%) | 26-Mar-2021 | | 685,706  (38%) | 28-Apr-2021 | 882,795  (73%) | 5-Jun-2021 | 1,439,030  (71%) | 13-May-2021 | 689,698  (47%) |

Supplement Table 5: Controllable spread date and number of cases on December 31, 2021 for different vaccination effectiveness scenarios when vaccination effectiveness is modeled as a reduction in probability of becoming infected (vaccine coverage 50%, vaccination capacity 0.01%). Numbers in parentheses represent percent reduction relative to no vaccine.

| NYC | | | | | | | | | | |
| --- | --- | --- | --- | --- | --- | --- | --- | --- | --- | --- |
|  | 90% Adherence | | 85% Adherence | | 80% Adherence | | 75% Adherence | | Dynamic Adherence | |
| Vaccine effectiveness | Date | Number of cases | Date | Number of cases | Date | Number of cases | Date | Number of cases | Date | Number of cases |
| No Vaccine | 22-Mar-2021 | 699,352 | 27-Aug-2021 | 1,110,070 | 4-Sep-2021 | 3,307,090 | 21-Jun-2021 | 4,999,100 | After June 2022 | 1,308,990 |
| 50% | 18-Mar-2021 | 683,968  (2%) | 18-Jun-2021 | 966,379  (13%) | 20-Aug-2021 | 2,670,380  (19%) | 21-Jun-2021 | 4,519,590  (10%) | After June 2022 | 1,276,990  (2%) |
| 75% | 17-Mar-2021 | 678,460  (3%) | 6-Jun-2021 | 930,525  (16%) | 10-Aug-2021 | 2,430,490  (27%) | 20-Jun-2021 | 4,288,620  (14%) | After June 2022 | 1,257,190  (4%) |
| 90% | 17-Mar-2021 | 674,462  (4%) | 30-May-2021 | 909,986  (18%) | 3-Aug-2021 | 2,287,610  (31%) | 19-Jun-2021 | 4,132,220  (17%) | 9-Feb-2022 | 1,193,500  (9%) |

Supplement Figure 5. Impact of vaccine coverage on the number of confirmed cases in different regions for two different scenarios of adherence to nonpharmaceutical interventions when vaccination effectiveness is modeled as a reduction in probability of becoming infected. In this figure, we assumed that vaccine effectiveness is equal to 90%, vaccine capacity is 0.1%/day.

a. NYC, adherence to NPIs is fixed at a level of 85%

Total number of confirmed cases

|  | No Vaccine | 25% | 50% | 60% | 75% | 100% Vaccinated |
| --- | --- | --- | --- | --- | --- | --- |
| 2020-03-15 | 0 | 0 | 0 | 0 | 0 | 0 |
| 2020-03-16 | 0 | 0 | 0 | 0 | 0 | 0 |
| 2020-03-17 | 0 | 0 | 0 | 0 | 0 | 0 |
| 2020-03-18 | 0 | 0 | 0 | 0 | 0 | 0 |
| 2020-03-19 | 0 | 0 | 0 | 0 | 0 | 0 |
| 2020-03-20 | 0 | 0 | 0 | 0 | 0 | 0 |
| 2020-03-21 | 0 | 0 | 0 | 0 | 0 | 0 |
| 2020-03-22 | 0 | 0 | 0 | 0 | 0 | 0 |
| 2020-03-23 | 0 | 0 | 0 | 0 | 0 | 0 |
| 2020-03-24 | 0 | 0 | 0 | 0 | 0 | 0 |
| 2020-03-25 | 0 | 0 | 0 | 0 | 0 | 0 |
| 2020-03-26 | 31 | 31 | 31 | 31 | 31 | 31 |
| 2020-03-27 | 61 | 61 | 61 | 61 | 61 | 61 |
| 2020-03-28 | 96 | 96 | 96 | 96 | 96 | 96 |
| 2020-03-29 | 156 | 156 | 156 | 156 | 156 | 156 |
| 2020-03-30 | 249 | 249 | 249 | 249 | 249 | 249 |
| 2020-03-31 | 360 | 360 | 360 | 360 | 360 | 360 |
| 2020-04-01 | 535 | 535 | 535 | 535 | 535 | 535 |
| 2020-04-02 | 773 | 773 | 773 | 773 | 773 | 773 |
| 2020-04-03 | 1083 | 1083 | 1083 | 1083 | 1083 | 1083 |
| 2020-04-04 | 1476 | 1476 | 1476 | 1476 | 1476 | 1476 |
| 2020-04-05 | 1955 | 1955 | 1955 | 1955 | 1955 | 1955 |
| 2020-04-06 | 2510 | 2510 | 2510 | 2510 | 2510 | 2510 |
| 2020-04-07 | 3152 | 3152 | 3152 | 3152 | 3152 | 3152 |
| 2020-04-08 | 3808 | 3808 | 3808 | 3808 | 3808 | 3808 |
| 2020-04-09 | 4434 | 4434 | 4434 | 4434 | 4434 | 4434 |
| 2020-04-10 | 4939 | 4939 | 4939 | 4939 | 4939 | 4939 |
| 2020-04-11 | 5540 | 5540 | 5540 | 5540 | 5540 | 5540 |
| 2020-04-12 | 6223 | 6223 | 6223 | 6223 | 6223 | 6223 |
| 2020-04-13 | 6936 | 6936 | 6936 | 6936 | 6936 | 6936 |
| 2020-04-14 | 7687 | 7687 | 7687 | 7687 | 7687 | 7687 |
| 2020-04-15 | 8488 | 8488 | 8488 | 8488 | 8488 | 8488 |
| 2020-04-16 | 9316 | 9316 | 9316 | 9316 | 9316 | 9316 |
| 2020-04-17 | 10202 | 10202 | 10202 | 10202 | 10202 | 10202 |
| 2020-04-18 | 11085 | 11085 | 11085 | 11085 | 11085 | 11085 |
| 2020-04-19 | 11997 | 11997 | 11997 | 11997 | 11997 | 11997 |
| 2020-04-20 | 12953 | 12953 | 12953 | 12953 | 12953 | 12953 |
| 2020-04-21 | 13976 | 13976 | 13976 | 13976 | 13976 | 13976 |
| 2020-04-22 | 15054 | 15054 | 15054 | 15054 | 15054 | 15054 |
| 2020-04-23 | 16175 | 16175 | 16175 | 16175 | 16175 | 16175 |
| 2020-04-24 | 17348 | 17348 | 17348 | 17348 | 17348 | 17348 |
| 2020-04-25 | 18536 | 18536 | 18536 | 18536 | 18536 | 18536 |
| 2020-04-26 | 19829 | 19829 | 19829 | 19829 | 19829 | 19829 |
| 2020-04-27 | 21159 | 21159 | 21159 | 21159 | 21159 | 21159 |
| 2020-04-28 | 22505 | 22505 | 22505 | 22505 | 22505 | 22505 |
| 2020-04-29 | 23953 | 23953 | 23953 | 23953 | 23953 | 23953 |
| 2020-04-30 | 25472 | 25472 | 25472 | 25472 | 25472 | 25472 |
| 2020-05-01 | 27024 | 27024 | 27024 | 27024 | 27024 | 27024 |
| 2020-05-02 | 28690 | 28690 | 28690 | 28690 | 28690 | 28690 |
| 2020-05-03 | 30401 | 30401 | 30401 | 30401 | 30401 | 30401 |
| 2020-05-04 | 31300 | 31300 | 31300 | 31300 | 31300 | 31300 |
| 2020-05-05 | 32244 | 32244 | 32244 | 32244 | 32244 | 32244 |
| 2020-05-06 | 33237 | 33237 | 33237 | 33237 | 33237 | 33237 |
| 2020-05-07 | 34171 | 34171 | 34171 | 34171 | 34171 | 34171 |
| 2020-05-08 | 35083 | 35083 | 35083 | 35083 | 35083 | 35083 |
| 2020-05-09 | 35956 | 35956 | 35956 | 35956 | 35956 | 35956 |
| 2020-05-10 | 36801 | 36801 | 36801 | 36801 | 36801 | 36801 |
| 2020-05-11 | 37591 | 37591 | 37591 | 37591 | 37591 | 37591 |
| 2020-05-12 | 38320 | 38320 | 38320 | 38320 | 38320 | 38320 |
| 2020-05-13 | 38972 | 38972 | 38972 | 38972 | 38972 | 38972 |
| 2020-05-14 | 39553 | 39553 | 39553 | 39553 | 39553 | 39553 |
| 2020-05-15 | 40127 | 40127 | 40127 | 40127 | 40127 | 40127 |
| 2020-05-16 | 40683 | 40683 | 40683 | 40683 | 40683 | 40683 |
| 2020-05-17 | 41187 | 41187 | 41187 | 41187 | 41187 | 41187 |
| 2020-05-18 | 41690 | 41690 | 41690 | 41690 | 41690 | 41690 |
| 2020-05-19 | 42151 | 42151 | 42151 | 42151 | 42151 | 42151 |
| 2020-05-20 | 42593 | 42593 | 42593 | 42593 | 42593 | 42593 |
| 2020-05-21 | 43008 | 43008 | 43008 | 43008 | 43008 | 43008 |
| 2020-05-22 | 43380 | 43380 | 43380 | 43380 | 43380 | 43380 |
| 2020-05-23 | 43747 | 43747 | 43747 | 43747 | 43747 | 43747 |
| 2020-05-24 | 44080 | 44080 | 44080 | 44080 | 44080 | 44080 |
| 2020-05-25 | 44394 | 44394 | 44394 | 44394 | 44394 | 44394 |
| 2020-05-26 | 44698 | 44698 | 44698 | 44698 | 44698 | 44698 |
| 2020-05-27 | 44983 | 44983 | 44983 | 44983 | 44983 | 44983 |
| 2020-05-28 | 45243 | 45243 | 45243 | 45243 | 45243 | 45243 |
| 2020-05-29 | 45499 | 45499 | 45499 | 45499 | 45499 | 45499 |
| 2020-05-30 | 45740 | 45740 | 45740 | 45740 | 45740 | 45740 |
| 2020-05-31 | 45964 | 45964 | 45964 | 45964 | 45964 | 45964 |
| 2020-06-01 | 46156 | 46156 | 46156 | 46156 | 46156 | 46156 |
| 2020-06-02 | 46344 | 46344 | 46344 | 46344 | 46344 | 46344 |
| 2020-06-03 | 46522 | 46522 | 46522 | 46522 | 46522 | 46522 |
| 2020-06-04 | 46685 | 46685 | 46685 | 46685 | 46685 | 46685 |
| 2020-06-05 | 46840 | 46840 | 46840 | 46840 | 46840 | 46840 |
| 2020-06-06 | 46997 | 46997 | 46997 | 46997 | 46997 | 46997 |
| 2020-06-07 | 47136 | 47136 | 47136 | 47136 | 47136 | 47136 |
| 2020-06-08 | 47275 | 47275 | 47275 | 47275 | 47275 | 47275 |
| 2020-06-09 | 47404 | 47404 | 47404 | 47404 | 47404 | 47404 |
| 2020-06-10 | 47533 | 47533 | 47533 | 47533 | 47533 | 47533 |
| 2020-06-11 | 47648 | 47648 | 47648 | 47648 | 47648 | 47648 |
| 2020-06-12 | 47758 | 47758 | 47758 | 47758 | 47758 | 47758 |
| 2020-06-13 | 47862 | 47862 | 47862 | 47862 | 47862 | 47862 |
| 2020-06-14 | 47963 | 47963 | 47963 | 47963 | 47963 | 47963 |
| 2020-06-15 | 48053 | 48053 | 48053 | 48053 | 48053 | 48053 |
| 2020-06-16 | 48145 | 48145 | 48145 | 48145 | 48145 | 48145 |
| 2020-06-17 | 48232 | 48232 | 48232 | 48232 | 48232 | 48232 |
| 2020-06-18 | 48310 | 48310 | 48310 | 48310 | 48310 | 48310 |
| 2020-06-19 | 48387 | 48387 | 48387 | 48387 | 48387 | 48387 |
| 2020-06-20 | 48456 | 48456 | 48456 | 48456 | 48456 | 48456 |
| 2020-06-21 | 48522 | 48522 | 48522 | 48522 | 48522 | 48522 |
| 2020-06-22 | 48592 | 48592 | 48592 | 48592 | 48592 | 48592 |
| 2020-06-23 | 48656 | 48656 | 48656 | 48656 | 48656 | 48656 |
| 2020-06-24 | 48710 | 48710 | 48710 | 48710 | 48710 | 48710 |
| 2020-06-25 | 48765 | 48765 | 48765 | 48765 | 48765 | 48765 |
| 2020-06-26 | 48822 | 48822 | 48822 | 48822 | 48822 | 48822 |
| 2020-06-27 | 48873 | 48873 | 48873 | 48873 | 48873 | 48873 |
| 2020-06-28 | 48925 | 48925 | 48925 | 48925 | 48925 | 48925 |
| 2020-06-29 | 48986 | 48986 | 48986 | 48986 | 48986 | 48986 |
| 2020-06-30 | 49048 | 49048 | 49048 | 49048 | 49048 | 49048 |
| 2020-07-01 | 49115 | 49115 | 49115 | 49115 | 49115 | 49115 |
| 2020-07-02 | 49175 | 49175 | 49175 | 49175 | 49175 | 49175 |
| 2020-07-03 | 49231 | 49231 | 49231 | 49231 | 49231 | 49231 |
| 2020-07-04 | 49288 | 49288 | 49288 | 49288 | 49288 | 49288 |
| 2020-07-05 | 49347 | 49347 | 49347 | 49347 | 49347 | 49347 |
| 2020-07-06 | 49407 | 49407 | 49407 | 49407 | 49407 | 49407 |
| 2020-07-07 | 49470 | 49470 | 49470 | 49470 | 49470 | 49470 |
| 2020-07-08 | 49536 | 49536 | 49536 | 49536 | 49536 | 49536 |
| 2020-07-09 | 49603 | 49603 | 49603 | 49603 | 49603 | 49603 |
| 2020-07-10 | 49670 | 49670 | 49670 | 49670 | 49670 | 49670 |
| 2020-07-11 | 49732 | 49732 | 49732 | 49732 | 49732 | 49732 |
| 2020-07-12 | 49792 | 49792 | 49792 | 49792 | 49792 | 49792 |
| 2020-07-13 | 49855 | 49855 | 49855 | 49855 | 49855 | 49855 |
| 2020-07-14 | 49921 | 49921 | 49921 | 49921 | 49921 | 49921 |
| 2020-07-15 | 49989 | 49989 | 49989 | 49989 | 49989 | 49989 |
| 2020-07-16 | 50056 | 50056 | 50056 | 50056 | 50056 | 50056 |
| 2020-07-17 | 50127 | 50127 | 50127 | 50127 | 50127 | 50127 |
| 2020-07-18 | 50197 | 50197 | 50197 | 50197 | 50197 | 50197 |
| 2020-07-19 | 50268 | 50268 | 50268 | 50268 | 50268 | 50268 |
| 2020-07-20 | 50340 | 50340 | 50340 | 50340 | 50340 | 50340 |
| 2020-07-21 | 50409 | 50409 | 50409 | 50409 | 50409 | 50409 |
| 2020-07-22 | 50482 | 50482 | 50482 | 50482 | 50482 | 50482 |
| 2020-07-23 | 50531 | 50531 | 50531 | 50531 | 50531 | 50531 |
| 2020-07-24 | 50582 | 50582 | 50582 | 50582 | 50582 | 50582 |
| 2020-07-25 | 50631 | 50631 | 50631 | 50631 | 50631 | 50631 |
| 2020-07-26 | 50683 | 50683 | 50683 | 50683 | 50683 | 50683 |
| 2020-07-27 | 50737 | 50737 | 50737 | 50737 | 50737 | 50737 |
| 2020-07-28 | 50789 | 50789 | 50789 | 50789 | 50789 | 50789 |
| 2020-07-29 | 50836 | 50836 | 50836 | 50836 | 50836 | 50836 |
| 2020-07-30 | 50876 | 50876 | 50876 | 50876 | 50876 | 50876 |
| 2020-07-31 | 50916 | 50916 | 50916 | 50916 | 50916 | 50916 |
| 2020-08-01 | 50956 | 50956 | 50956 | 50956 | 50956 | 50956 |
| 2020-08-02 | 50992 | 50992 | 50992 | 50992 | 50992 | 50992 |
| 2020-08-03 | 51028 | 51028 | 51028 | 51028 | 51028 | 51028 |
| 2020-08-04 | 51066 | 51066 | 51066 | 51066 | 51066 | 51066 |
| 2020-08-05 | 51105 | 51105 | 51105 | 51105 | 51105 | 51105 |
| 2020-08-06 | 51139 | 51139 | 51139 | 51139 | 51139 | 51139 |
| 2020-08-07 | 51173 | 51173 | 51173 | 51173 | 51173 | 51173 |
| 2020-08-08 | 51204 | 51204 | 51204 | 51204 | 51204 | 51204 |
| 2020-08-09 | 51236 | 51236 | 51236 | 51236 | 51236 | 51236 |
| 2020-08-10 | 51268 | 51268 | 51268 | 51268 | 51268 | 51268 |
| 2020-08-11 | 51297 | 51297 | 51297 | 51297 | 51297 | 51297 |
| 2020-08-12 | 51325 | 51325 | 51325 | 51325 | 51325 | 51325 |
| 2020-08-13 | 51352 | 51352 | 51352 | 51352 | 51352 | 51352 |
| 2020-08-14 | 51378 | 51378 | 51378 | 51378 | 51378 | 51378 |
| 2020-08-15 | 51403 | 51403 | 51403 | 51403 | 51403 | 51403 |
| 2020-08-16 | 51428 | 51428 | 51428 | 51428 | 51428 | 51428 |
| 2020-08-17 | 51454 | 51454 | 51454 | 51454 | 51454 | 51454 |
| 2020-08-18 | 51479 | 51479 | 51479 | 51479 | 51479 | 51479 |
| 2020-08-19 | 51503 | 51503 | 51503 | 51503 | 51503 | 51503 |
| 2020-08-20 | 51527 | 51527 | 51527 | 51527 | 51527 | 51527 |
| 2020-08-21 | 51550 | 51550 | 51550 | 51550 | 51550 | 51550 |
| 2020-08-22 | 51572 | 51572 | 51572 | 51572 | 51572 | 51572 |
| 2020-08-23 | 51594 | 51594 | 51594 | 51594 | 51594 | 51594 |
| 2020-08-24 | 51617 | 51617 | 51617 | 51617 | 51617 | 51617 |
| 2020-08-25 | 51637 | 51637 | 51637 | 51637 | 51637 | 51637 |
| 2020-08-26 | 51657 | 51657 | 51657 | 51657 | 51657 | 51657 |
| 2020-08-27 | 51679 | 51679 | 51679 | 51679 | 51679 | 51679 |
| 2020-08-28 | 51700 | 51700 | 51700 | 51700 | 51700 | 51700 |
| 2020-08-29 | 51717 | 51717 | 51717 | 51717 | 51717 | 51717 |
| 2020-08-30 | 51741 | 51741 | 51741 | 51741 | 51741 | 51741 |
| 2020-08-31 | 51762 | 51762 | 51762 | 51762 | 51762 | 51762 |
| 2020-09-01 | 51783 | 51783 | 51783 | 51783 | 51783 | 51783 |
| 2020-09-02 | 51800 | 51800 | 51800 | 51800 | 51800 | 51800 |
| 2020-09-03 | 51821 | 51821 | 51821 | 51821 | 51821 | 51821 |
| 2020-09-04 | 51839 | 51839 | 51839 | 51839 | 51839 | 51839 |
| 2020-09-05 | 51861 | 51861 | 51861 | 51861 | 51861 | 51861 |
| 2020-09-06 | 51895 | 51895 | 51895 | 51895 | 51895 | 51895 |
| 2020-09-07 | 51927 | 51927 | 51927 | 51927 | 51927 | 51927 |
| 2020-09-08 | 51959 | 51959 | 51959 | 51959 | 51959 | 51959 |
| 2020-09-09 | 51993 | 51993 | 51993 | 51993 | 51993 | 51993 |
| 2020-09-10 | 52033 | 52033 | 52033 | 52033 | 52033 | 52033 |
| 2020-09-11 | 52072 | 52072 | 52072 | 52072 | 52072 | 52072 |
| 2020-09-12 | 52115 | 52115 | 52115 | 52115 | 52115 | 52115 |
| 2020-09-13 | 52159 | 52159 | 52159 | 52159 | 52159 | 52159 |
| 2020-09-14 | 52209 | 52209 | 52209 | 52209 | 52209 | 52209 |
| 2020-09-15 | 52264 | 52264 | 52264 | 52264 | 52264 | 52264 |
| 2020-09-16 | 52313 | 52313 | 52313 | 52313 | 52313 | 52313 |
| 2020-09-17 | 52367 | 52367 | 52367 | 52367 | 52367 | 52367 |
| 2020-09-18 | 52422 | 52422 | 52422 | 52422 | 52422 | 52422 |
| 2020-09-19 | 52495 | 52495 | 52495 | 52495 | 52495 | 52495 |
| 2020-09-20 | 52564 | 52564 | 52564 | 52564 | 52564 | 52564 |
| 2020-09-21 | 52639 | 52639 | 52639 | 52639 | 52639 | 52639 |
| 2020-09-22 | 52715 | 52715 | 52715 | 52715 | 52715 | 52715 |
| 2020-09-23 | 52799 | 52799 | 52799 | 52799 | 52799 | 52799 |
| 2020-09-24 | 52884 | 52884 | 52884 | 52884 | 52884 | 52884 |
| 2020-09-25 | 52975 | 52975 | 52975 | 52975 | 52975 | 52975 |
| 2020-09-26 | 53066 | 53066 | 53066 | 53066 | 53066 | 53066 |
| 2020-09-27 | 53160 | 53160 | 53160 | 53160 | 53160 | 53160 |
| 2020-09-28 | 53257 | 53257 | 53257 | 53257 | 53257 | 53257 |
| 2020-09-29 | 53365 | 53365 | 53365 | 53365 | 53365 | 53365 |
| 2020-09-30 | 53476 | 53476 | 53476 | 53476 | 53476 | 53476 |

b. NYC, adherence to NPIs is dynamic

Total number of confirmed cases

|  | No Vaccine | 25% | 50% | 60% | 75% | 100% Vaccinated |
| --- | --- | --- | --- | --- | --- | --- |
| 2020-03-15 | 0 | 0 | 0 | 0 | 0 | 0 |
| 2020-03-16 | 0 | 0 | 0 | 0 | 0 | 0 |
| 2020-03-17 | 0 | 0 | 0 | 0 | 0 | 0 |
| 2020-03-18 | 0 | 0 | 0 | 0 | 0 | 0 |
| 2020-03-19 | 0 | 0 | 0 | 0 | 0 | 0 |
| 2020-03-20 | 0 | 0 | 0 | 0 | 0 | 0 |
| 2020-03-21 | 0 | 0 | 0 | 0 | 0 | 0 |
| 2020-03-22 | 0 | 0 | 0 | 0 | 0 | 0 |
| 2020-03-23 | 0 | 0 | 0 | 0 | 0 | 0 |
| 2020-03-24 | 0 | 0 | 0 | 0 | 0 | 0 |
| 2020-03-25 | 0 | 0 | 0 | 0 | 0 | 0 |
| 2020-03-26 | 31 | 31 | 31 | 31 | 31 | 31 |
| 2020-03-27 | 61 | 61 | 61 | 61 | 61 | 61 |
| 2020-03-28 | 96 | 96 | 96 | 96 | 96 | 96 |
| 2020-03-29 | 156 | 156 | 156 | 156 | 156 | 156 |
| 2020-03-30 | 249 | 249 | 249 | 249 | 249 | 249 |
| 2020-03-31 | 360 | 360 | 360 | 360 | 360 | 360 |
| 2020-04-01 | 535 | 535 | 535 | 535 | 535 | 535 |
| 2020-04-02 | 773 | 773 | 773 | 773 | 773 | 773 |
| 2020-04-03 | 1083 | 1083 | 1083 | 1083 | 1083 | 1083 |
| 2020-04-04 | 1476 | 1476 | 1476 | 1476 | 1476 | 1476 |
| 2020-04-05 | 1955 | 1955 | 1955 | 1955 | 1955 | 1955 |
| 2020-04-06 | 2510 | 2510 | 2510 | 2510 | 2510 | 2510 |
| 2020-04-07 | 3152 | 3152 | 3152 | 3152 | 3152 | 3152 |
| 2020-04-08 | 3808 | 3808 | 3808 | 3808 | 3808 | 3808 |
| 2020-04-09 | 4434 | 4434 | 4434 | 4434 | 4434 | 4434 |
| 2020-04-10 | 4939 | 4939 | 4939 | 4939 | 4939 | 4939 |
| 2020-04-11 | 5540 | 5540 | 5540 | 5540 | 5540 | 5540 |
| 2020-04-12 | 6223 | 6223 | 6223 | 6223 | 6223 | 6223 |
| 2020-04-13 | 6936 | 6936 | 6936 | 6936 | 6936 | 6936 |
| 2020-04-14 | 7687 | 7687 | 7687 | 7687 | 7687 | 7687 |
| 2020-04-15 | 8488 | 8488 | 8488 | 8488 | 8488 | 8488 |
| 2020-04-16 | 9316 | 9316 | 9316 | 9316 | 9316 | 9316 |
| 2020-04-17 | 10202 | 10202 | 10202 | 10202 | 10202 | 10202 |
| 2020-04-18 | 11085 | 11085 | 11085 | 11085 | 11085 | 11085 |
| 2020-04-19 | 11997 | 11997 | 11997 | 11997 | 11997 | 11997 |
| 2020-04-20 | 12953 | 12953 | 12953 | 12953 | 12953 | 12953 |
| 2020-04-21 | 13976 | 13976 | 13976 | 13976 | 13976 | 13976 |
| 2020-04-22 | 15054 | 15054 | 15054 | 15054 | 15054 | 15054 |
| 2020-04-23 | 16175 | 16175 | 16175 | 16175 | 16175 | 16175 |
| 2020-04-24 | 17348 | 17348 | 17348 | 17348 | 17348 | 17348 |
| 2020-04-25 | 18536 | 18536 | 18536 | 18536 | 18536 | 18536 |
| 2020-04-26 | 19829 | 19829 | 19829 | 19829 | 19829 | 19829 |
| 2020-04-27 | 21159 | 21159 | 21159 | 21159 | 21159 | 21159 |
| 2020-04-28 | 22505 | 22505 | 22505 | 22505 | 22505 | 22505 |
| 2020-04-29 | 23953 | 23953 | 23953 | 23953 | 23953 | 23953 |
| 2020-04-30 | 25472 | 25472 | 25472 | 25472 | 25472 | 25472 |
| 2020-05-01 | 27024 | 27024 | 27024 | 27024 | 27024 | 27024 |
| 2020-05-02 | 28690 | 28690 | 28690 | 28690 | 28690 | 28690 |
| 2020-05-03 | 30401 | 30401 | 30401 | 30401 | 30401 | 30401 |
| 2020-05-04 | 31300 | 31300 | 31300 | 31300 | 31300 | 31300 |
| 2020-05-05 | 32244 | 32244 | 32244 | 32244 | 32244 | 32244 |
| 2020-05-06 | 33237 | 33237 | 33237 | 33237 | 33237 | 33237 |
| 2020-05-07 | 34171 | 34171 | 34171 | 34171 | 34171 | 34171 |
| 2020-05-08 | 35083 | 35083 | 35083 | 35083 | 35083 | 35083 |
| 2020-05-09 | 35956 | 35956 | 35956 | 35956 | 35956 | 35956 |
| 2020-05-10 | 36801 | 36801 | 36801 | 36801 | 36801 | 36801 |
| 2020-05-11 | 37591 | 37591 | 37591 | 37591 | 37591 | 37591 |
| 2020-05-12 | 38320 | 38320 | 38320 | 38320 | 38320 | 38320 |
| 2020-05-13 | 38972 | 38972 | 38972 | 38972 | 38972 | 38972 |
| 2020-05-14 | 39553 | 39553 | 39553 | 39553 | 39553 | 39553 |
| 2020-05-15 | 40127 | 40127 | 40127 | 40127 | 40127 | 40127 |
| 2020-05-16 | 40683 | 40683 | 40683 | 40683 | 40683 | 40683 |
| 2020-05-17 | 41187 | 41187 | 41187 | 41187 | 41187 | 41187 |
| 2020-05-18 | 41690 | 41690 | 41690 | 41690 | 41690 | 41690 |
| 2020-05-19 | 42151 | 42151 | 42151 | 42151 | 42151 | 42151 |
| 2020-05-20 | 42593 | 42593 | 42593 | 42593 | 42593 | 42593 |
| 2020-05-21 | 43008 | 43008 | 43008 | 43008 | 43008 | 43008 |
| 2020-05-22 | 43380 | 43380 | 43380 | 43380 | 43380 | 43380 |
| 2020-05-23 | 43747 | 43747 | 43747 | 43747 | 43747 | 43747 |
| 2020-05-24 | 44080 | 44080 | 44080 | 44080 | 44080 | 44080 |
| 2020-05-25 | 44394 | 44394 | 44394 | 44394 | 44394 | 44394 |
| 2020-05-26 | 44698 | 44698 | 44698 | 44698 | 44698 | 44698 |
| 2020-05-27 | 44983 | 44983 | 44983 | 44983 | 44983 | 44983 |
| 2020-05-28 | 45243 | 45243 | 45243 | 45243 | 45243 | 45243 |
| 2020-05-29 | 45499 | 45499 | 45499 | 45499 | 45499 | 45499 |
| 2020-05-30 | 45740 | 45740 | 45740 | 45740 | 45740 | 45740 |
| 2020-05-31 | 45964 | 45964 | 45964 | 45964 | 45964 | 45964 |
| 2020-06-01 | 46156 | 46156 | 46156 | 46156 | 46156 | 46156 |
| 2020-06-02 | 46344 | 46344 | 46344 | 46344 | 46344 | 46344 |
| 2020-06-03 | 46522 | 46522 | 46522 | 46522 | 46522 | 46522 |
| 2020-06-04 | 46685 | 46685 | 46685 | 46685 | 46685 | 46685 |
| 2020-06-05 | 46840 | 46840 | 46840 | 46840 | 46840 | 46840 |
| 2020-06-06 | 46997 | 46997 | 46997 | 46997 | 46997 | 46997 |
| 2020-06-07 | 47136 | 47136 | 47136 | 47136 | 47136 | 47136 |
| 2020-06-08 | 47275 | 47275 | 47275 | 47275 | 47275 | 47275 |
| 2020-06-09 | 47404 | 47404 | 47404 | 47404 | 47404 | 47404 |
| 2020-06-10 | 47533 | 47533 | 47533 | 47533 | 47533 | 47533 |
| 2020-06-11 | 47648 | 47648 | 47648 | 47648 | 47648 | 47648 |
| 2020-06-12 | 47758 | 47758 | 47758 | 47758 | 47758 | 47758 |
| 2020-06-13 | 47862 | 47862 | 47862 | 47862 | 47862 | 47862 |
| 2020-06-14 | 47963 | 47963 | 47963 | 47963 | 47963 | 47963 |
| 2020-06-15 | 48053 | 48053 | 48053 | 48053 | 48053 | 48053 |
| 2020-06-16 | 48145 | 48145 | 48145 | 48145 | 48145 | 48145 |
| 2020-06-17 | 48232 | 48232 | 48232 | 48232 | 48232 | 48232 |
| 2020-06-18 | 48310 | 48310 | 48310 | 48310 | 48310 | 48310 |
| 2020-06-19 | 48387 | 48387 | 48387 | 48387 | 48387 | 48387 |
| 2020-06-20 | 48456 | 48456 | 48456 | 48456 | 48456 | 48456 |
| 2020-06-21 | 48522 | 48522 | 48522 | 48522 | 48522 | 48522 |
| 2020-06-22 | 48592 | 48592 | 48592 | 48592 | 48592 | 48592 |
| 2020-06-23 | 48656 | 48656 | 48656 | 48656 | 48656 | 48656 |
| 2020-06-24 | 48710 | 48710 | 48710 | 48710 | 48710 | 48710 |
| 2020-06-25 | 48765 | 48765 | 48765 | 48765 | 48765 | 48765 |
| 2020-06-26 | 48822 | 48822 | 48822 | 48822 | 48822 | 48822 |
| 2020-06-27 | 48873 | 48873 | 48873 | 48873 | 48873 | 48873 |
| 2020-06-28 | 48925 | 48925 | 48925 | 48925 | 48925 | 48925 |
| 2020-06-29 | 48986 | 48986 | 48986 | 48986 | 48986 | 48986 |
| 2020-06-30 | 49048 | 49048 | 49048 | 49048 | 49048 | 49048 |
| 2020-07-01 | 49115 | 49115 | 49115 | 49115 | 49115 | 49115 |
| 2020-07-02 | 49175 | 49175 | 49175 | 49175 | 49175 | 49175 |
| 2020-07-03 | 49231 | 49231 | 49231 | 49231 | 49231 | 49231 |
| 2020-07-04 | 49288 | 49288 | 49288 | 49288 | 49288 | 49288 |
| 2020-07-05 | 49347 | 49347 | 49347 | 49347 | 49347 | 49347 |
| 2020-07-06 | 49407 | 49407 | 49407 | 49407 | 49407 | 49407 |
| 2020-07-07 | 49470 | 49470 | 49470 | 49470 | 49470 | 49470 |
| 2020-07-08 | 49536 | 49536 | 49536 | 49536 | 49536 | 49536 |
| 2020-07-09 | 49603 | 49603 | 49603 | 49603 | 49603 | 49603 |
| 2020-07-10 | 49670 | 49670 | 49670 | 49670 | 49670 | 49670 |
| 2020-07-11 | 49732 | 49732 | 49732 | 49732 | 49732 | 49732 |
| 2020-07-12 | 49792 | 49792 | 49792 | 49792 | 49792 | 49792 |
| 2020-07-13 | 49855 | 49855 | 49855 | 49855 | 49855 | 49855 |
| 2020-07-14 | 49921 | 49921 | 49921 | 49921 | 49921 | 49921 |
| 2020-07-15 | 49989 | 49989 | 49989 | 49989 | 49989 | 49989 |
| 2020-07-16 | 50056 | 50056 | 50056 | 50056 | 50056 | 50056 |
| 2020-07-17 | 50127 | 50127 | 50127 | 50127 | 50127 | 50127 |
| 2020-07-18 | 50197 | 50197 | 50197 | 50197 | 50197 | 50197 |
| 2020-07-19 | 50268 | 50268 | 50268 | 50268 | 50268 | 50268 |
| 2020-07-20 | 50340 | 50340 | 50340 | 50340 | 50340 | 50340 |
| 2020-07-21 | 50409 | 50409 | 50409 | 50409 | 50409 | 50409 |
| 2020-07-22 | 50482 | 50482 | 50482 | 50482 | 50482 | 50482 |
| 2020-07-23 | 50531 | 50531 | 50531 | 50531 | 50531 | 50531 |
| 2020-07-24 | 50582 | 50582 | 50582 | 50582 | 50582 | 50582 |
| 2020-07-25 | 50631 | 50631 | 50631 | 50631 | 50631 | 50631 |
| 2020-07-26 | 50683 | 50683 | 50683 | 50683 | 50683 | 50683 |
| 2020-07-27 | 50737 | 50737 | 50737 | 50737 | 50737 | 50737 |
| 2020-07-28 | 50789 | 50789 | 50789 | 50789 | 50789 | 50789 |
| 2020-07-29 | 50836 | 50836 | 50836 | 50836 | 50836 | 50836 |
| 2020-07-30 | 50876 | 50876 | 50876 | 50876 | 50876 | 50876 |
| 2020-07-31 | 50916 | 50916 | 50916 | 50916 | 50916 | 50916 |
| 2020-08-01 | 50956 | 50956 | 50956 | 50956 | 50956 | 50956 |
| 2020-08-02 | 50992 | 50992 | 50992 | 50992 | 50992 | 50992 |
| 2020-08-03 | 51028 | 51028 | 51028 | 51028 | 51028 | 51028 |
| 2020-08-04 | 51066 | 51066 | 51066 | 51066 | 51066 | 51066 |
| 2020-08-05 | 51105 | 51105 | 51105 | 51105 | 51105 | 51105 |
| 2020-08-06 | 51139 | 51139 | 51139 | 51139 | 51139 | 51139 |
| 2020-08-07 | 51173 | 51173 | 51173 | 51173 | 51173 | 51173 |
| 2020-08-08 | 51204 | 51204 | 51204 | 51204 | 51204 | 51204 |
| 2020-08-09 | 51236 | 51236 | 51236 | 51236 | 51236 | 51236 |
| 2020-08-10 | 51268 | 51268 | 51268 | 51268 | 51268 | 51268 |
| 2020-08-11 | 51297 | 51297 | 51297 | 51297 | 51297 | 51297 |
| 2020-08-12 | 51325 | 51325 | 51325 | 51325 | 51325 | 51325 |
| 2020-08-13 | 51352 | 51352 | 51352 | 51352 | 51352 | 51352 |
| 2020-08-14 | 51378 | 51378 | 51378 | 51378 | 51378 | 51378 |
| 2020-08-15 | 51403 | 51403 | 51403 | 51403 | 51403 | 51403 |
| 2020-08-16 | 51428 | 51428 | 51428 | 51428 | 51428 | 51428 |
| 2020-08-17 | 51454 | 51454 | 51454 | 51454 | 51454 | 51454 |
| 2020-08-18 | 51479 | 51479 | 51479 | 51479 | 51479 | 51479 |
| 2020-08-19 | 51503 | 51503 | 51503 | 51503 | 51503 | 51503 |
| 2020-08-20 | 51527 | 51527 | 51527 | 51527 | 51527 | 51527 |
| 2020-08-21 | 51550 | 51550 | 51550 | 51550 | 51550 | 51550 |
| 2020-08-22 | 51572 | 51572 | 51572 | 51572 | 51572 | 51572 |
| 2020-08-23 | 51594 | 51594 | 51594 | 51594 | 51594 | 51594 |
| 2020-08-24 | 51617 | 51617 | 51617 | 51617 | 51617 | 51617 |
| 2020-08-25 | 51637 | 51637 | 51637 | 51637 | 51637 | 51637 |
| 2020-08-26 | 51657 | 51657 | 51657 | 51657 | 51657 | 51657 |
| 2020-08-27 | 51679 | 51679 | 51679 | 51679 | 51679 | 51679 |
| 2020-08-28 | 51700 | 51700 | 51700 | 51700 | 51700 | 51700 |
| 2020-08-29 | 51717 | 51717 | 51717 | 51717 | 51717 | 51717 |
| 2020-08-30 | 51741 | 51741 | 51741 | 51741 | 51741 | 51741 |
| 2020-08-31 | 51762 | 51762 | 51762 | 51762 | 51762 | 51762 |
| 2020-09-01 | 51783 | 51783 | 51783 | 51783 | 51783 | 51783 |
| 2020-09-02 | 51800 | 51800 | 51800 | 51800 | 51800 | 51800 |
| 2020-09-03 | 51821 | 51821 | 51821 | 51821 | 51821 | 51821 |
| 2020-09-04 | 51839 | 51839 | 51839 | 51839 | 51839 | 51839 |
| 2020-09-05 | 51861 | 51861 | 51861 | 51861 | 51861 | 51861 |
| 2020-09-06 | 51895 | 51895 | 51895 | 51895 | 51895 | 51895 |
| 2020-09-07 | 51927 | 51927 | 51927 | 51927 | 51927 | 51927 |
| 2020-09-08 | 51959 | 51959 | 51959 | 51959 | 51959 | 51959 |
| 2020-09-09 | 51993 | 51993 | 51993 | 51993 | 51993 | 51993 |
| 2020-09-10 | 52033 | 52033 | 52033 | 52033 | 52033 | 52033 |
| 2020-09-11 | 52072 | 52072 | 52072 | 52072 | 52072 | 52072 |
| 2020-09-12 | 52115 | 52115 | 52115 | 52115 | 52115 | 52115 |
| 2020-09-13 | 52159 | 52159 | 52159 | 52159 | 52159 | 52159 |
| 2020-09-14 | 52209 | 52209 | 52209 | 52209 | 52209 | 52209 |
| 2020-09-15 | 52264 | 52264 | 52264 | 52264 | 52264 | 52264 |
| 2020-09-16 | 52313 | 52313 | 52313 | 52313 | 52313 | 52313 |
| 2020-09-17 | 52367 | 52367 | 52367 | 52367 | 52367 | 52367 |
| 2020-09-18 | 52422 | 52422 | 52422 | 52422 | 52422 | 52422 |
| 2020-09-19 | 52495 | 52495 | 52495 | 52495 | 52495 | 52495 |
| 2020-09-20 | 52564 | 52564 | 52564 | 52564 | 52564 | 52564 |
| 2020-09-21 | 52639 | 52639 | 52639 | 52639 | 52639 | 52639 |
| 2020-09-22 | 52715 | 52715 | 52715 | 52715 | 52715 | 52715 |
| 2020-09-23 | 52799 | 52799 | 52799 | 52799 | 52799 | 52799 |
| 2020-09-24 | 52884 | 52884 | 52884 | 52884 | 52884 | 52884 |
| 2020-09-25 | 52975 | 52975 | 52975 | 52975 | 52975 | 52975 |
| 2020-09-26 | 53066 | 53066 | 53066 | 53066 | 53066 | 53066 |
| 2020-09-27 | 53160 | 53160 | 53160 | 53160 | 53160 | 53160 |
| 2020-09-28 | 53257 | 53257 | 53257 | 53257 | 53257 | 53257 |
| 2020-09-29 | 53365 | 53365 | 53365 | 53365 | 53365 | 53365 |
| 2020-09-30 | 53476 | 53476 | 53476 | 53476 | 53476 | 53476 |

Supplement Figure 6. Impact of vaccination and adherence to nonpharmaceutical interventions on the number of cases over time when vaccination effectiveness is modeled as a reduction in probability of becoming infected. In these experiments, we assumed that vaccine effectiveness is 90%, vaccine coverage is 50%, and daily vaccination capacity is 0.1% for the vaccination scenario.

a. NYC, vaccination

Total number of confirmed cases

|  | 90% Adherence | 85% | 80% | 75% | Dynamic Adherence |
| --- | --- | --- | --- | --- | --- |
| 2020-03-15 | 0 | 0 | 0 | 0 | 0 |
| 2020-03-16 | 0 | 0 | 0 | 0 | 0 |
| 2020-03-17 | 0 | 0 | 0 | 0 | 0 |
| 2020-03-18 | 0 | 0 | 0 | 0 | 0 |
| 2020-03-19 | 0 | 0 | 0 | 0 | 0 |
| 2020-03-20 | 0 | 0 | 0 | 0 | 0 |
| 2020-03-21 | 0 | 0 | 0 | 0 | 0 |
| 2020-03-22 | 0 | 0 | 0 | 0 | 0 |
| 2020-03-23 | 0 | 0 | 0 | 0 | 0 |
| 2020-03-24 | 0 | 0 | 0 | 0 | 0 |
| 2020-03-25 | 0 | 0 | 0 | 0 | 0 |
| 2020-03-26 | 31 | 31 | 31 | 31 | 31 |
| 2020-03-27 | 61 | 61 | 61 | 61 | 61 |
| 2020-03-28 | 96 | 96 | 96 | 96 | 96 |
| 2020-03-29 | 156 | 156 | 156 | 156 | 156 |
| 2020-03-30 | 249 | 249 | 249 | 249 | 249 |
| 2020-03-31 | 360 | 360 | 360 | 360 | 360 |
| 2020-04-01 | 535 | 535 | 535 | 535 | 535 |
| 2020-04-02 | 773 | 773 | 773 | 773 | 773 |
| 2020-04-03 | 1083 | 1083 | 1083 | 1083 | 1083 |
| 2020-04-04 | 1476 | 1476 | 1476 | 1476 | 1476 |
| 2020-04-05 | 1955 | 1955 | 1955 | 1955 | 1955 |
| 2020-04-06 | 2510 | 2510 | 2510 | 2510 | 2510 |
| 2020-04-07 | 3152 | 3152 | 3152 | 3152 | 3152 |
| 2020-04-08 | 3808 | 3808 | 3808 | 3808 | 3808 |
| 2020-04-09 | 4434 | 4434 | 4434 | 4434 | 4434 |
| 2020-04-10 | 4939 | 4939 | 4939 | 4939 | 4939 |
| 2020-04-11 | 5540 | 5540 | 5540 | 5540 | 5540 |
| 2020-04-12 | 6223 | 6223 | 6223 | 6223 | 6223 |
| 2020-04-13 | 6936 | 6936 | 6936 | 6936 | 6936 |
| 2020-04-14 | 7687 | 7687 | 7687 | 7687 | 7687 |
| 2020-04-15 | 8488 | 8488 | 8488 | 8488 | 8488 |
| 2020-04-16 | 9316 | 9316 | 9316 | 9316 | 9316 |
| 2020-04-17 | 10202 | 10202 | 10202 | 10202 | 10202 |
| 2020-04-18 | 11085 | 11085 | 11085 | 11085 | 11085 |
| 2020-04-19 | 11997 | 11997 | 11997 | 11997 | 11997 |
| 2020-04-20 | 12953 | 12953 | 12953 | 12953 | 12953 |
| 2020-04-21 | 13976 | 13976 | 13976 | 13976 | 13976 |
| 2020-04-22 | 15054 | 15054 | 15054 | 15054 | 15054 |
| 2020-04-23 | 16175 | 16175 | 16175 | 16175 | 16175 |
| 2020-04-24 | 17348 | 17348 | 17348 | 17348 | 17348 |
| 2020-04-25 | 18536 | 18536 | 18536 | 18536 | 18536 |
| 2020-04-26 | 19829 | 19829 | 19829 | 19829 | 19829 |
| 2020-04-27 | 21159 | 21159 | 21159 | 21159 | 21159 |
| 2020-04-28 | 22505 | 22505 | 22505 | 22505 | 22505 |
| 2020-04-29 | 23953 | 23953 | 23953 | 23953 | 23953 |
| 2020-04-30 | 25472 | 25472 | 25472 | 25472 | 25472 |
| 2020-05-01 | 27024 | 27024 | 27024 | 27024 | 27024 |
| 2020-05-02 | 28690 | 28690 | 28690 | 28690 | 28690 |
| 2020-05-03 | 30401 | 30401 | 30401 | 30401 | 30401 |
| 2020-05-04 | 31300 | 31300 | 31300 | 31300 | 31300 |
| 2020-05-05 | 32244 | 32244 | 32244 | 32244 | 32244 |
| 2020-05-06 | 33237 | 33237 | 33237 | 33237 | 33237 |
| 2020-05-07 | 34171 | 34171 | 34171 | 34171 | 34171 |
| 2020-05-08 | 35083 | 35083 | 35083 | 35083 | 35083 |
| 2020-05-09 | 35956 | 35956 | 35956 | 35956 | 35956 |
| 2020-05-10 | 36801 | 36801 | 36801 | 36801 | 36801 |
| 2020-05-11 | 37591 | 37591 | 37591 | 37591 | 37591 |
| 2020-05-12 | 38320 | 38320 | 38320 | 38320 | 38320 |
| 2020-05-13 | 38972 | 38972 | 38972 | 38972 | 38972 |
| 2020-05-14 | 39553 | 39553 | 39553 | 39553 | 39553 |
| 2020-05-15 | 40127 | 40127 | 40127 | 40127 | 40127 |
| 2020-05-16 | 40683 | 40683 | 40683 | 40683 | 40683 |
| 2020-05-17 | 41187 | 41187 | 41187 | 41187 | 41187 |
| 2020-05-18 | 41690 | 41690 | 41690 | 41690 | 41690 |
| 2020-05-19 | 42151 | 42151 | 42151 | 42151 | 42151 |
| 2020-05-20 | 42593 | 42593 | 42593 | 42593 | 42593 |
| 2020-05-21 | 43008 | 43008 | 43008 | 43008 | 43008 |
| 2020-05-22 | 43380 | 43380 | 43380 | 43380 | 43380 |
| 2020-05-23 | 43747 | 43747 | 43747 | 43747 | 43747 |
| 2020-05-24 | 44080 | 44080 | 44080 | 44080 | 44080 |
| 2020-05-25 | 44394 | 44394 | 44394 | 44394 | 44394 |
| 2020-05-26 | 44698 | 44698 | 44698 | 44698 | 44698 |
| 2020-05-27 | 44983 | 44983 | 44983 | 44983 | 44983 |
| 2020-05-28 | 45243 | 45243 | 45243 | 45243 | 45243 |
| 2020-05-29 | 45499 | 45499 | 45499 | 45499 | 45499 |
| 2020-05-30 | 45740 | 45740 | 45740 | 45740 | 45740 |
| 2020-05-31 | 45964 | 45964 | 45964 | 45964 | 45964 |
| 2020-06-01 | 46156 | 46156 | 46156 | 46156 | 46156 |
| 2020-06-02 | 46344 | 46344 | 46344 | 46344 | 46344 |
| 2020-06-03 | 46522 | 46522 | 46522 | 46522 | 46522 |
| 2020-06-04 | 46685 | 46685 | 46685 | 46685 | 46685 |
| 2020-06-05 | 46840 | 46840 | 46840 | 46840 | 46840 |
| 2020-06-06 | 46997 | 46997 | 46997 | 46997 | 46997 |
| 2020-06-07 | 47136 | 47136 | 47136 | 47136 | 47136 |
| 2020-06-08 | 47275 | 47275 | 47275 | 47275 | 47275 |
| 2020-06-09 | 47404 | 47404 | 47404 | 47404 | 47404 |
| 2020-06-10 | 47533 | 47533 | 47533 | 47533 | 47533 |
| 2020-06-11 | 47648 | 47648 | 47648 | 47648 | 47648 |
| 2020-06-12 | 47758 | 47758 | 47758 | 47758 | 47758 |
| 2020-06-13 | 47862 | 47862 | 47862 | 47862 | 47862 |
| 2020-06-14 | 47963 | 47963 | 47963 | 47963 | 47963 |
| 2020-06-15 | 48053 | 48053 | 48053 | 48053 | 48053 |
| 2020-06-16 | 48145 | 48145 | 48145 | 48145 | 48145 |
| 2020-06-17 | 48232 | 48232 | 48232 | 48232 | 48232 |
| 2020-06-18 | 48310 | 48310 | 48310 | 48310 | 48310 |
| 2020-06-19 | 48387 | 48387 | 48387 | 48387 | 48387 |
| 2020-06-20 | 48456 | 48456 | 48456 | 48456 | 48456 |
| 2020-06-21 | 48522 | 48522 | 48522 | 48522 | 48522 |
| 2020-06-22 | 48592 | 48592 | 48592 | 48592 | 48592 |
| 2020-06-23 | 48656 | 48656 | 48656 | 48656 | 48656 |
| 2020-06-24 | 48710 | 48710 | 48710 | 48710 | 48710 |
| 2020-06-25 | 48765 | 48765 | 48765 | 48765 | 48765 |
| 2020-06-26 | 48822 | 48822 | 48822 | 48822 | 48822 |
| 2020-06-27 | 48873 | 48873 | 48873 | 48873 | 48873 |
| 2020-06-28 | 48925 | 48925 | 48925 | 48925 | 48925 |
| 2020-06-29 | 48986 | 48986 | 48986 | 48986 | 48986 |
| 2020-06-30 | 49048 | 49048 | 49048 | 49048 | 49048 |
| 2020-07-01 | 49115 | 49115 | 49115 | 49115 | 49115 |
| 2020-07-02 | 49175 | 49175 | 49175 | 49175 | 49175 |
| 2020-07-03 | 49231 | 49231 | 49231 | 49231 | 49231 |
| 2020-07-04 | 49288 | 49288 | 49288 | 49288 | 49288 |
| 2020-07-05 | 49347 | 49347 | 49347 | 49347 | 49347 |
| 2020-07-06 | 49407 | 49407 | 49407 | 49407 | 49407 |
| 2020-07-07 | 49470 | 49470 | 49470 | 49470 | 49470 |
| 2020-07-08 | 49536 | 49536 | 49536 | 49536 | 49536 |
| 2020-07-09 | 49603 | 49603 | 49603 | 49603 | 49603 |
| 2020-07-10 | 49670 | 49670 | 49670 | 49670 | 49670 |
| 2020-07-11 | 49732 | 49732 | 49732 | 49732 | 49732 |
| 2020-07-12 | 49792 | 49792 | 49792 | 49792 | 49792 |
| 2020-07-13 | 49855 | 49855 | 49855 | 49855 | 49855 |
| 2020-07-14 | 49921 | 49921 | 49921 | 49921 | 49921 |
| 2020-07-15 | 49989 | 49989 | 49989 | 49989 | 49989 |
| 2020-07-16 | 50056 | 50056 | 50056 | 50056 | 50056 |
| 2020-07-17 | 50127 | 50127 | 50127 | 50127 | 50127 |
| 2020-07-18 | 50197 | 50197 | 50197 | 50197 | 50197 |
| 2020-07-19 | 50268 | 50268 | 50268 | 50268 | 50268 |
| 2020-07-20 | 50340 | 50340 | 50340 | 50340 | 50340 |
| 2020-07-21 | 50409 | 50409 | 50409 | 50409 | 50409 |
| 2020-07-22 | 50482 | 50482 | 50482 | 50482 | 50482 |
| 2020-07-23 | 50531 | 50531 | 50531 | 50531 | 50531 |
| 2020-07-24 | 50582 | 50582 | 50582 | 50582 | 50582 |
| 2020-07-25 | 50631 | 50631 | 50631 | 50631 | 50631 |
| 2020-07-26 | 50683 | 50683 | 50683 | 50683 | 50683 |
| 2020-07-27 | 50737 | 50737 | 50737 | 50737 | 50737 |
| 2020-07-28 | 50789 | 50789 | 50789 | 50789 | 50789 |
| 2020-07-29 | 50836 | 50836 | 50836 | 50836 | 50836 |
| 2020-07-30 | 50876 | 50876 | 50876 | 50876 | 50876 |
| 2020-07-31 | 50916 | 50916 | 50916 | 50916 | 50916 |
| 2020-08-01 | 50956 | 50956 | 50956 | 50956 | 50956 |
| 2020-08-02 | 50992 | 50992 | 50992 | 50992 | 50992 |
| 2020-08-03 | 51028 | 51028 | 51028 | 51028 | 51028 |
| 2020-08-04 | 51066 | 51066 | 51066 | 51066 | 51066 |
| 2020-08-05 | 51105 | 51105 | 51105 | 51105 | 51105 |
| 2020-08-06 | 51139 | 51139 | 51139 | 51139 | 51139 |
| 2020-08-07 | 51173 | 51173 | 51173 | 51173 | 51173 |
| 2020-08-08 | 51204 | 51204 | 51204 | 51204 | 51204 |
| 2020-08-09 | 51236 | 51236 | 51236 | 51236 | 51236 |
| 2020-08-10 | 51268 | 51268 | 51268 | 51268 | 51268 |
| 2020-08-11 | 51297 | 51297 | 51297 | 51297 | 51297 |
| 2020-08-12 | 51325 | 51325 | 51325 | 51325 | 51325 |
| 2020-08-13 | 51352 | 51352 | 51352 | 51352 | 51352 |
| 2020-08-14 | 51378 | 51378 | 51378 | 51378 | 51378 |
| 2020-08-15 | 51403 | 51403 | 51403 | 51403 | 51403 |
| 2020-08-16 | 51428 | 51428 | 51428 | 51428 | 51428 |
| 2020-08-17 | 51454 | 51454 | 51454 | 51454 | 51454 |
| 2020-08-18 | 51479 | 51479 | 51479 | 51479 | 51479 |
| 2020-08-19 | 51503 | 51503 | 51503 | 51503 | 51503 |
| 2020-08-20 | 51527 | 51527 | 51527 | 51527 | 51527 |
| 2020-08-21 | 51550 | 51550 | 51550 | 51550 | 51550 |
| 2020-08-22 | 51572 | 51572 | 51572 | 51572 | 51572 |
| 2020-08-23 | 51594 | 51594 | 51594 | 51594 | 51594 |
| 2020-08-24 | 51617 | 51617 | 51617 | 51617 | 51617 |
| 2020-08-25 | 51637 | 51637 | 51637 | 51637 | 51637 |
| 2020-08-26 | 51657 | 51657 | 51657 | 51657 | 51657 |
| 2020-08-27 | 51679 | 51679 | 51679 | 51679 | 51679 |
| 2020-08-28 | 51700 | 51700 | 51700 | 51700 | 51700 |
| 2020-08-29 | 51717 | 51717 | 51717 | 51717 | 51717 |
| 2020-08-30 | 51741 | 51741 | 51741 | 51741 | 51741 |
| 2020-08-31 | 51762 | 51762 | 51762 | 51762 | 51762 |
| 2020-09-01 | 51783 | 51783 | 51783 | 51783 | 51783 |
| 2020-09-02 | 51800 | 51800 | 51800 | 51800 | 51800 |
| 2020-09-03 | 51821 | 51821 | 51821 | 51821 | 51821 |
| 2020-09-04 | 51839 | 51839 | 51839 | 51839 | 51839 |
| 2020-09-05 | 51861 | 51861 | 51861 | 51861 | 51861 |
| 2020-09-06 | 51895 | 51895 | 51895 | 51895 | 51895 |
| 2020-09-07 | 51927 | 51927 | 51927 | 51927 | 51927 |
| 2020-09-08 | 51959 | 51959 | 51959 | 51959 | 51959 |
| 2020-09-09 | 51993 | 51993 | 51993 | 51993 | 51993 |
| 2020-09-10 | 52033 | 52033 | 52033 | 52033 | 52033 |
| 2020-09-11 | 52072 | 52072 | 52072 | 52072 | 52072 |
| 2020-09-12 | 52115 | 52115 | 52115 | 52115 | 52115 |
| 2020-09-13 | 52159 | 52159 | 52159 | 52159 | 52159 |
| 2020-09-14 | 52209 | 52209 | 52209 | 52209 | 52209 |
| 2020-09-15 | 52264 | 52264 | 52264 | 52264 | 52264 |
| 2020-09-16 | 52313 | 52313 | 52313 | 52313 | 52313 |
| 2020-09-17 | 52367 | 52367 | 52367 | 52367 | 52367 |
| 2020-09-18 | 52422 | 52422 | 52422 | 52422 | 52422 |
| 2020-09-19 | 52495 | 52495 | 52495 | 52495 | 52495 |
| 2020-09-20 | 52564 | 52564 | 52564 | 52564 | 52564 |
| 2020-09-21 | 52639 | 52639 | 52639 | 52639 | 52639 |
| 2020-09-22 | 52715 | 52715 | 52715 | 52715 | 52715 |
| 2020-09-23 | 52799 | 52799 | 52799 | 52799 | 52799 |
| 2020-09-24 | 52884 | 52884 | 52884 | 52884 | 52884 |
| 2020-09-25 | 52975 | 52975 | 52975 | 52975 | 52975 |
| 2020-09-26 | 53066 | 53066 | 53066 | 53066 | 53066 |
| 2020-09-27 | 53160 | 53160 | 53160 | 53160 | 53160 |
| 2020-09-28 | 53257 | 53257 | 53257 | 53257 | 53257 |
| 2020-09-29 | 53365 | 53365 | 53365 | 53365 | 53365 |
| 2020-09-30 | 53476 | 53476 | 53476 | 53476 | 53476 |

b. NYC, no vaccination

Total number of confirmed cases

|  | 90% Adherence | 85% | 80% | 75% | Dynamic Adherence |
| --- | --- | --- | --- | --- | --- |
| 2020-03-15 | 0 | 0 | 0 | 0 | 0 |
| 2020-03-16 | 0 | 0 | 0 | 0 | 0 |
| 2020-03-17 | 0 | 0 | 0 | 0 | 0 |
| 2020-03-18 | 0 | 0 | 0 | 0 | 0 |
| 2020-03-19 | 0 | 0 | 0 | 0 | 0 |
| 2020-03-20 | 0 | 0 | 0 | 0 | 0 |
| 2020-03-21 | 0 | 0 | 0 | 0 | 0 |
| 2020-03-22 | 0 | 0 | 0 | 0 | 0 |
| 2020-03-23 | 0 | 0 | 0 | 0 | 0 |
| 2020-03-24 | 0 | 0 | 0 | 0 | 0 |
| 2020-03-25 | 0 | 0 | 0 | 0 | 0 |
| 2020-03-26 | 31 | 31 | 31 | 31 | 31 |
| 2020-03-27 | 61 | 61 | 61 | 61 | 61 |
| 2020-03-28 | 96 | 96 | 96 | 96 | 96 |
| 2020-03-29 | 156 | 156 | 156 | 156 | 156 |
| 2020-03-30 | 249 | 249 | 249 | 249 | 249 |
| 2020-03-31 | 360 | 360 | 360 | 360 | 360 |
| 2020-04-01 | 535 | 535 | 535 | 535 | 535 |
| 2020-04-02 | 773 | 773 | 773 | 773 | 773 |
| 2020-04-03 | 1083 | 1083 | 1083 | 1083 | 1083 |
| 2020-04-04 | 1476 | 1476 | 1476 | 1476 | 1476 |
| 2020-04-05 | 1955 | 1955 | 1955 | 1955 | 1955 |
| 2020-04-06 | 2510 | 2510 | 2510 | 2510 | 2510 |
| 2020-04-07 | 3152 | 3152 | 3152 | 3152 | 3152 |
| 2020-04-08 | 3808 | 3808 | 3808 | 3808 | 3808 |
| 2020-04-09 | 4434 | 4434 | 4434 | 4434 | 4434 |
| 2020-04-10 | 4939 | 4939 | 4939 | 4939 | 4939 |
| 2020-04-11 | 5540 | 5540 | 5540 | 5540 | 5540 |
| 2020-04-12 | 6223 | 6223 | 6223 | 6223 | 6223 |
| 2020-04-13 | 6936 | 6936 | 6936 | 6936 | 6936 |
| 2020-04-14 | 7687 | 7687 | 7687 | 7687 | 7687 |
| 2020-04-15 | 8488 | 8488 | 8488 | 8488 | 8488 |
| 2020-04-16 | 9316 | 9316 | 9316 | 9316 | 9316 |
| 2020-04-17 | 10202 | 10202 | 10202 | 10202 | 10202 |
| 2020-04-18 | 11085 | 11085 | 11085 | 11085 | 11085 |
| 2020-04-19 | 11997 | 11997 | 11997 | 11997 | 11997 |
| 2020-04-20 | 12953 | 12953 | 12953 | 12953 | 12953 |
| 2020-04-21 | 13976 | 13976 | 13976 | 13976 | 13976 |
| 2020-04-22 | 15054 | 15054 | 15054 | 15054 | 15054 |
| 2020-04-23 | 16175 | 16175 | 16175 | 16175 | 16175 |
| 2020-04-24 | 17348 | 17348 | 17348 | 17348 | 17348 |
| 2020-04-25 | 18536 | 18536 | 18536 | 18536 | 18536 |
| 2020-04-26 | 19829 | 19829 | 19829 | 19829 | 19829 |
| 2020-04-27 | 21159 | 21159 | 21159 | 21159 | 21159 |
| 2020-04-28 | 22505 | 22505 | 22505 | 22505 | 22505 |
| 2020-04-29 | 23953 | 23953 | 23953 | 23953 | 23953 |
| 2020-04-30 | 25472 | 25472 | 25472 | 25472 | 25472 |
| 2020-05-01 | 27024 | 27024 | 27024 | 27024 | 27024 |
| 2020-05-02 | 28690 | 28690 | 28690 | 28690 | 28690 |
| 2020-05-03 | 30401 | 30401 | 30401 | 30401 | 30401 |
| 2020-05-04 | 31300 | 31300 | 31300 | 31300 | 31300 |
| 2020-05-05 | 32244 | 32244 | 32244 | 32244 | 32244 |
| 2020-05-06 | 33237 | 33237 | 33237 | 33237 | 33237 |
| 2020-05-07 | 34171 | 34171 | 34171 | 34171 | 34171 |
| 2020-05-08 | 35083 | 35083 | 35083 | 35083 | 35083 |
| 2020-05-09 | 35956 | 35956 | 35956 | 35956 | 35956 |
| 2020-05-10 | 36801 | 36801 | 36801 | 36801 | 36801 |
| 2020-05-11 | 37591 | 37591 | 37591 | 37591 | 37591 |
| 2020-05-12 | 38320 | 38320 | 38320 | 38320 | 38320 |
| 2020-05-13 | 38972 | 38972 | 38972 | 38972 | 38972 |
| 2020-05-14 | 39553 | 39553 | 39553 | 39553 | 39553 |
| 2020-05-15 | 40127 | 40127 | 40127 | 40127 | 40127 |
| 2020-05-16 | 40683 | 40683 | 40683 | 40683 | 40683 |
| 2020-05-17 | 41187 | 41187 | 41187 | 41187 | 41187 |
| 2020-05-18 | 41690 | 41690 | 41690 | 41690 | 41690 |
| 2020-05-19 | 42151 | 42151 | 42151 | 42151 | 42151 |
| 2020-05-20 | 42593 | 42593 | 42593 | 42593 | 42593 |
| 2020-05-21 | 43008 | 43008 | 43008 | 43008 | 43008 |
| 2020-05-22 | 43380 | 43380 | 43380 | 43380 | 43380 |
| 2020-05-23 | 43747 | 43747 | 43747 | 43747 | 43747 |
| 2020-05-24 | 44080 | 44080 | 44080 | 44080 | 44080 |
| 2020-05-25 | 44394 | 44394 | 44394 | 44394 | 44394 |
| 2020-05-26 | 44698 | 44698 | 44698 | 44698 | 44698 |
| 2020-05-27 | 44983 | 44983 | 44983 | 44983 | 44983 |
| 2020-05-28 | 45243 | 45243 | 45243 | 45243 | 45243 |
| 2020-05-29 | 45499 | 45499 | 45499 | 45499 | 45499 |
| 2020-05-30 | 45740 | 45740 | 45740 | 45740 | 45740 |
| 2020-05-31 | 45964 | 45964 | 45964 | 45964 | 45964 |
| 2020-06-01 | 46156 | 46156 | 46156 | 46156 | 46156 |
| 2020-06-02 | 46344 | 46344 | 46344 | 46344 | 46344 |
| 2020-06-03 | 46522 | 46522 | 46522 | 46522 | 46522 |
| 2020-06-04 | 46685 | 46685 | 46685 | 46685 | 46685 |
| 2020-06-05 | 46840 | 46840 | 46840 | 46840 | 46840 |
| 2020-06-06 | 46997 | 46997 | 46997 | 46997 | 46997 |
| 2020-06-07 | 47136 | 47136 | 47136 | 47136 | 47136 |
| 2020-06-08 | 47275 | 47275 | 47275 | 47275 | 47275 |
| 2020-06-09 | 47404 | 47404 | 47404 | 47404 | 47404 |
| 2020-06-10 | 47533 | 47533 | 47533 | 47533 | 47533 |
| 2020-06-11 | 47648 | 47648 | 47648 | 47648 | 47648 |
| 2020-06-12 | 47758 | 47758 | 47758 | 47758 | 47758 |
| 2020-06-13 | 47862 | 47862 | 47862 | 47862 | 47862 |
| 2020-06-14 | 47963 | 47963 | 47963 | 47963 | 47963 |
| 2020-06-15 | 48053 | 48053 | 48053 | 48053 | 48053 |
| 2020-06-16 | 48145 | 48145 | 48145 | 48145 | 48145 |
| 2020-06-17 | 48232 | 48232 | 48232 | 48232 | 48232 |
| 2020-06-18 | 48310 | 48310 | 48310 | 48310 | 48310 |
| 2020-06-19 | 48387 | 48387 | 48387 | 48387 | 48387 |
| 2020-06-20 | 48456 | 48456 | 48456 | 48456 | 48456 |
| 2020-06-21 | 48522 | 48522 | 48522 | 48522 | 48522 |
| 2020-06-22 | 48592 | 48592 | 48592 | 48592 | 48592 |
| 2020-06-23 | 48656 | 48656 | 48656 | 48656 | 48656 |
| 2020-06-24 | 48710 | 48710 | 48710 | 48710 | 48710 |
| 2020-06-25 | 48765 | 48765 | 48765 | 48765 | 48765 |
| 2020-06-26 | 48822 | 48822 | 48822 | 48822 | 48822 |
| 2020-06-27 | 48873 | 48873 | 48873 | 48873 | 48873 |
| 2020-06-28 | 48925 | 48925 | 48925 | 48925 | 48925 |
| 2020-06-29 | 48986 | 48986 | 48986 | 48986 | 48986 |
| 2020-06-30 | 49048 | 49048 | 49048 | 49048 | 49048 |
| 2020-07-01 | 49115 | 49115 | 49115 | 49115 | 49115 |
| 2020-07-02 | 49175 | 49175 | 49175 | 49175 | 49175 |
| 2020-07-03 | 49231 | 49231 | 49231 | 49231 | 49231 |
| 2020-07-04 | 49288 | 49288 | 49288 | 49288 | 49288 |
| 2020-07-05 | 49347 | 49347 | 49347 | 49347 | 49347 |
| 2020-07-06 | 49407 | 49407 | 49407 | 49407 | 49407 |
| 2020-07-07 | 49470 | 49470 | 49470 | 49470 | 49470 |
| 2020-07-08 | 49536 | 49536 | 49536 | 49536 | 49536 |
| 2020-07-09 | 49603 | 49603 | 49603 | 49603 | 49603 |
| 2020-07-10 | 49670 | 49670 | 49670 | 49670 | 49670 |
| 2020-07-11 | 49732 | 49732 | 49732 | 49732 | 49732 |
| 2020-07-12 | 49792 | 49792 | 49792 | 49792 | 49792 |
| 2020-07-13 | 49855 | 49855 | 49855 | 49855 | 49855 |
| 2020-07-14 | 49921 | 49921 | 49921 | 49921 | 49921 |
| 2020-07-15 | 49989 | 49989 | 49989 | 49989 | 49989 |
| 2020-07-16 | 50056 | 50056 | 50056 | 50056 | 50056 |
| 2020-07-17 | 50127 | 50127 | 50127 | 50127 | 50127 |
| 2020-07-18 | 50197 | 50197 | 50197 | 50197 | 50197 |
| 2020-07-19 | 50268 | 50268 | 50268 | 50268 | 50268 |
| 2020-07-20 | 50340 | 50340 | 50340 | 50340 | 50340 |
| 2020-07-21 | 50409 | 50409 | 50409 | 50409 | 50409 |
| 2020-07-22 | 50482 | 50482 | 50482 | 50482 | 50482 |
| 2020-07-23 | 50531 | 50531 | 50531 | 50531 | 50531 |
| 2020-07-24 | 50582 | 50582 | 50582 | 50582 | 50582 |
| 2020-07-25 | 50631 | 50631 | 50631 | 50631 | 50631 |
| 2020-07-26 | 50683 | 50683 | 50683 | 50683 | 50683 |
| 2020-07-27 | 50737 | 50737 | 50737 | 50737 | 50737 |
| 2020-07-28 | 50789 | 50789 | 50789 | 50789 | 50789 |
| 2020-07-29 | 50836 | 50836 | 50836 | 50836 | 50836 |
| 2020-07-30 | 50876 | 50876 | 50876 | 50876 | 50876 |
| 2020-07-31 | 50916 | 50916 | 50916 | 50916 | 50916 |
| 2020-08-01 | 50956 | 50956 | 50956 | 50956 | 50956 |
| 2020-08-02 | 50992 | 50992 | 50992 | 50992 | 50992 |
| 2020-08-03 | 51028 | 51028 | 51028 | 51028 | 51028 |
| 2020-08-04 | 51066 | 51066 | 51066 | 51066 | 51066 |
| 2020-08-05 | 51105 | 51105 | 51105 | 51105 | 51105 |
| 2020-08-06 | 51139 | 51139 | 51139 | 51139 | 51139 |
| 2020-08-07 | 51173 | 51173 | 51173 | 51173 | 51173 |
| 2020-08-08 | 51204 | 51204 | 51204 | 51204 | 51204 |
| 2020-08-09 | 51236 | 51236 | 51236 | 51236 | 51236 |
| 2020-08-10 | 51268 | 51268 | 51268 | 51268 | 51268 |
| 2020-08-11 | 51297 | 51297 | 51297 | 51297 | 51297 |
| 2020-08-12 | 51325 | 51325 | 51325 | 51325 | 51325 |
| 2020-08-13 | 51352 | 51352 | 51352 | 51352 | 51352 |
| 2020-08-14 | 51378 | 51378 | 51378 | 51378 | 51378 |
| 2020-08-15 | 51403 | 51403 | 51403 | 51403 | 51403 |
| 2020-08-16 | 51428 | 51428 | 51428 | 51428 | 51428 |
| 2020-08-17 | 51454 | 51454 | 51454 | 51454 | 51454 |
| 2020-08-18 | 51479 | 51479 | 51479 | 51479 | 51479 |
| 2020-08-19 | 51503 | 51503 | 51503 | 51503 | 51503 |
| 2020-08-20 | 51527 | 51527 | 51527 | 51527 | 51527 |
| 2020-08-21 | 51550 | 51550 | 51550 | 51550 | 51550 |
| 2020-08-22 | 51572 | 51572 | 51572 | 51572 | 51572 |
| 2020-08-23 | 51594 | 51594 | 51594 | 51594 | 51594 |
| 2020-08-24 | 51617 | 51617 | 51617 | 51617 | 51617 |
| 2020-08-25 | 51637 | 51637 | 51637 | 51637 | 51637 |
| 2020-08-26 | 51657 | 51657 | 51657 | 51657 | 51657 |
| 2020-08-27 | 51679 | 51679 | 51679 | 51679 | 51679 |
| 2020-08-28 | 51700 | 51700 | 51700 | 51700 | 51700 |
| 2020-08-29 | 51717 | 51717 | 51717 | 51717 | 51717 |
| 2020-08-30 | 51741 | 51741 | 51741 | 51741 | 51741 |
| 2020-08-31 | 51762 | 51762 | 51762 | 51762 | 51762 |
| 2020-09-01 | 51783 | 51783 | 51783 | 51783 | 51783 |
| 2020-09-02 | 51800 | 51800 | 51800 | 51800 | 51800 |
| 2020-09-03 | 51821 | 51821 | 51821 | 51821 | 51821 |
| 2020-09-04 | 51839 | 51839 | 51839 | 51839 | 51839 |
| 2020-09-05 | 51861 | 51861 | 51861 | 51861 | 51861 |
| 2020-09-06 | 51895 | 51895 | 51895 | 51895 | 51895 |
| 2020-09-07 | 51927 | 51927 | 51927 | 51927 | 51927 |
| 2020-09-08 | 51959 | 51959 | 51959 | 51959 | 51959 |
| 2020-09-09 | 51993 | 51993 | 51993 | 51993 | 51993 |
| 2020-09-10 | 52033 | 52033 | 52033 | 52033 | 52033 |
| 2020-09-11 | 52072 | 52072 | 52072 | 52072 | 52072 |
| 2020-09-12 | 52115 | 52115 | 52115 | 52115 | 52115 |
| 2020-09-13 | 52159 | 52159 | 52159 | 52159 | 52159 |
| 2020-09-14 | 52209 | 52209 | 52209 | 52209 | 52209 |
| 2020-09-15 | 52264 | 52264 | 52264 | 52264 | 52264 |
| 2020-09-16 | 52313 | 52313 | 52313 | 52313 | 52313 |
| 2020-09-17 | 52367 | 52367 | 52367 | 52367 | 52367 |
| 2020-09-18 | 52422 | 52422 | 52422 | 52422 | 52422 |
| 2020-09-19 | 52495 | 52495 | 52495 | 52495 | 52495 |
| 2020-09-20 | 52564 | 52564 | 52564 | 52564 | 52564 |
| 2020-09-21 | 52639 | 52639 | 52639 | 52639 | 52639 |
| 2020-09-22 | 52715 | 52715 | 52715 | 52715 | 52715 |
| 2020-09-23 | 52799 | 52799 | 52799 | 52799 | 52799 |
| 2020-09-24 | 52884 | 52884 | 52884 | 52884 | 52884 |
| 2020-09-25 | 52975 | 52975 | 52975 | 52975 | 52975 |
| 2020-09-26 | 53066 | 53066 | 53066 | 53066 | 53066 |
| 2020-09-27 | 53160 | 53160 | 53160 | 53160 | 53160 |
| 2020-09-28 | 53257 | 53257 | 53257 | 53257 | 53257 |
| 2020-09-29 | 53365 | 53365 | 53365 | 53365 | 53365 |
| 2020-09-30 | 53476 | 53476 | 53476 | 53476 | 53476 |
